# The genetic basis of hydrocephalus: genes, pathways, mechanisms, and global impact

**DOI:** 10.1101/2023.12.03.23299322

**Authors:** Andrew T. Hale, Hunter Boudreau, Rishi Devulapalli, Phan Q. Duy, Travis J. Atchley, Michael C. Dewan, Mubeen Goolam, Graham Fieggen, Heather L. Spader, Anastasia A. Smith, Jeffrey P. Blount, James M. Johnston, Brandon G. Rocque, Curtis J. Rozzelle, Zechen Chong, Jennifer M. Strahle, Steven J. Schiff, Kristopher T. Kahle

**Affiliations:** Department of Neurosurgery, University of Alabama at Birmingham, Birmingham AL; Heersink School of Medicine, University of Alabama at Birmingham, Birmingham, AL; Department of Neurosurgery, University of Virginia School of Medicine, Charlottesville, VA; Division of Pediatric Neurosurgery, Monroe Carell Jr. Children’s Hospital, Vanderbilt University School of Medicine, Nashville, TN; Neuroscience Institute, University of Cape Town, Cape Town, South Africa; Division of Pediatric Neurosurgery, Red Cross War Memorial Children’s Hospital, University of Cape Town, Cape Town, South Africa; Division of Pediatric Neurosurgery, Children’s of Alabama, University of Alabama at Birmingham, Birmingham, AL; Heflin Center for Genomics, University of Alabama at Birmingham, Birmingham, AL; Division of Pediatric Neurosurgery, St. Louis Children’s Hospital, Washington University in St. Louis, St. Louis, MO; Department of Neurosurgery, Yale University School of Medicine, New Haven, CT; Department of Neurosurgery, Massachusetts General Hospital, Harvard Medical School, Boston, MA

## Abstract

Hydrocephalus (HC) is a heterogenous disease characterized by alterations in cerebrospinal fluid (CSF) dynamics that may cause increased intracranial pressure. HC is a component of a wide array of genetic syndromes as well as a secondary consequence of brain injury (intraventricular hemorrhage (IVH), infection, etc.), highlighting the phenotypic heterogeneity of the disease. Surgical treatments include ventricular shunting and endoscopic third ventriculostomy with or without choroid plexus cauterization, both of which are prone to failure, and no effective pharmacologic treatments for HC have been developed. Thus, there is an urgent need to understand the genetic architecture and molecular pathogenesis of HC. Without this knowledge, the development of preventive, diagnostic, and therapeutic measures is impeded. However, the genetics of HC is extraordinarily complex, based on studies of varying size, scope, and rigor. This review serves to provide a comprehensive overview of genes, pathways, mechanisms, and global impact of genetics contributing to all etiologies of HC in humans.

## Introduction

Hydrocephalus (HC) is characterized by aberrant cerebrospinal fluid (CSF) dynamics (with or without ventricular dilation) that can lead to increased intracranial pressure. When left untreated, HC may be fatal and cause severe impairment in neurodevelopment. While classical theories of CSF posited that CSF is produced predominantly in the lateral ventricles via the choroid plexus and flows through the foramina of Monroe, third ventricle, cerebral aqueduct, and the fourth ventricle where it is disseminated through the central canal of the spinal cord and the subarachnoid space to be reabsorbed by arachnoid granulations [2], this model is no longer considered dogmatic [3]. Anatomical disruption of CSF flow and/or CSF pulsatility may result in a buildup of CSF to be classified as obstructive or non-communicating HC. However, HC can be communicating (i.e., no obvious anatomical blockade of absorption or obstruction), the result from increased production of CSF in response to injury, impaired absorption from the subarachnoid space, or result from defects in cortical development. These insults, in turn, may lead to ventricular dilation, among other potential and highly debated pathophysiologic mechanisms. Importantly, the global burden of HC is high [4], with significant morbidity and mortality regardless of treatment [5]. However, the genetic and mechanistic basis of HC remains poorly understood, largely due to the genetic complexity and phenotypic heterogeneity of the disease as well as cost of large-scale human genetics studies.

HC is a component of a wide-array of genetic syndromes [6], a secondary consequence of brain injury (intraventricular hemorrhage (IVH), infection, etc.) [7; 8], and a component of many central nervous system congenital abnormalities (i.e., neural tube defects, Chiari malformation, etc.) with a number of comorbid phenotypes including epilepsy and autism, among others. HC is a highly polygenic disease [9; 10; 11], with genes of varying functions and mechanisms conferring risk to the disease. The current treatments for HC are surgical interventions such as insertion of a ventricular (-peritoneal, atrial, etc.) shunt or endoscopic third ventriculostomy (ETV), which may be combined with choroid plexus cauterization (CPC) [7; 12]. While many studies have evaluated the efficacy and cost of these procedures [13], long-term morbidity of HC remains high and both treatments are prone to failure [14; 15]. Furthermore, while clinical trials have attempted pharmacological strategies to treat HC [16], no pharmacological treatment has been successful. A more sophisticated and detailed understanding the genetic architecture and molecular pathogenesis of HC may lead to development of targeted pharmacologic treatments.

While numerous studies have aimed to identify causative genetic mechanisms leading to HC, largely based on isolated human case studies and murine models [6], critical limitations include cost, patient/family recruitment, number of patients (small by population-genetics’ standards), individual variant validation (typically *de novo* mutations), and very important species differences between model-organisms and human disease. Proposed pathophysiological mechanisms of HC include impaired development of the neural stem cell niche [17; 18; 19; 20; 21], abnormal ciliated ependymal cells [22; 23; 24], disruption of the ventricular zone [25; 26], and primary alterations in CSF absorption and/or secretion [27; 28]. However, our understanding of these mechanisms is derived from varied model systems, which do not always accurately recapitulate the genetic and pathophysiological basis of human HC. Furthermore, there is increasing evidence that germline genetic variation contributes to risk of HC [6; 9; 11; 29]; however, most cases of HC remain genetically undefined and clinical genetic testing is rarely performed.

Elucidation of the genetic architecture of both shared and etiology-specific forms of HC may uncover pathophysiological mechanisms and correlate genetic risk factors with clinical and surgical outcomes, with the potential to directly influence surgical counseling and clinical management. While many genes have been implicated in the pathogenesis of HC in humans, the study designs, approaches, and levels of evidence identifying and validating these genetic findings vary greatly. Uncovering the genetic basis of HC relies on many factors, but most importantly on the clinical phenotype in question because HC rarely occurs in isolation. Comorbid phenotypes (neural tube defects, primary structural brain disorders, epilepsy, cognitive delay, etc.), and antecedent injuries – IVH and/or infection (meningitis, intracranial abscess, and/or sepsis), alone and in concert, confound most classical approaches to understanding genetic disease. Advancing our understanding of HC genetics, therefore, will necessitate understanding the extent to which co-occurring phenotypes are present and integration of multiple molecular and genetic data. Furthermore, elucidation of human-specific molecular mechanisms necessitates study in human tissue representative of the diverse populations HC affects. Here we summarize genetic studies of HC in humans and offer suggestions for advancing the field forward.

## Methods

### Search Criteria

The US National Library of Medicine PubMed database and the Online Mendelian Inheritance in Man (OMIM) were queried for English-language studies using Title/Abstract, MeSH headings, key words, and genetic descriptors relevant to genetic causes of HC and ventriculomegaly. The OMIM database was used as an additional adjunct database as well. Our search terms are included below. Duplicates identified across multiple databases were identified. We strictly adhered to PRISMA guidelines [30].

Our PubMed search syntax included the following: (HC[Title/Abstract]) OR (Ventriculomegaly[Title/Abstract]); ((HC[MeSH Major Topic]) OR (Ventriculomegaly[MeSH Major Topic])) AND (“mendelian” OR “de novo” OR “functional genomics” OR “whole exome sequencing” OR “whole-genome sequencing” OR “genotyping” OR “genotype” OR “microarray” OR “genome-wide association study” OR “genome wide association study” OR “GWAS” OR “transcriptome wide association study” OR “transcriptome-wide association study” OR “TWAS” OR “gene expression” OR “copy number variation” OR “insertion” OR “deletion” OR “mosaic” OR “mosaicism” OR “genetic variation” OR “consanguineous” OR “consanguinity” OR “autosomal recessive” OR “autosomal dominant” OR “x-linked recessive” OR “x-linked dominant” OR “inherited” OR “inheritance” OR “non-coding” OR “coding” OR “co-expression” OR “germline” OR “linkage” OR “linkage disequilibrium” OR “genetic counseling” OR “syndrome” OR “syndromic” OR “genetic testing” OR “aqueductal stenosis” OR “obstructive HC” OR “acquired HC” OR “congenital HC” OR “proteomics” OR “proteomic” OR “metabolomic” OR “metabolomics” OR “methylation” OR “mutation” OR “genetic deficiency” OR “gain of function” OR “gain-of-function” OR “loss of function” OR “loss-of-function” OR “molecular”[Title/Abstract]).

We next queried the Online Mendelian Inheritance in Man (OMIM) database [31] using the search terms: “HC” or “ventriculomegaly” to identify genetic disease of which HC is a component. The search returned 671 entries which were manually reviewed. Duplicates within the OMIM database were excluded (n=95). The resulting search query resulted in 3,709 studies.

### Inclusion and Exclusion Criteria

Records (n=3,709) from the above search were initially evaluated via abstract and screened for exclusion criteria: 1) Records published before 1970; 2) no genetic data of any kind; 3) no HC diagnosis; or 4) animal subjects. A total of 2,652 studies were excluded. Full text screening of the remaining papers (n=1,057) was then screened for inclusion criteria. The second round of screening was carried out by full text review (n=1,057). The same exclusion criteria were applied, while inclusion criteria were implemented: 1) Records published after 1970; 2) pediatric cohort (0-18 years of age); 3) primary genetic analysis; 4) confirmed diagnosis of human HC; and 5) human subjects. The final records were assessed for eligibility and records unavailable in English were excluded (n=2). The final studies included (n=332) were then evaluated for the methodology and type of genetic analysis performed. The papers included in our study (n=332) were then subject to secondary analyses to assess for 1) change in number of publications over time; 2) geographic and ethnic associations of congenital HC; 3) size of study; 4) central nervous system and non-central nervous system phenotypic associations. Figures were created using BioRender.

### Author Affiliation and Subject Country of Origin

Authors’ institutional affiliation was obtained via PubMed’s “Affiliations” tab within the respective research articles PubMed webpage. Authors were then cross-referenced via Google search to increase validity of institutional affiliation at the time the study was performed. Articles were individually queried for the country of origin of patients with HC. If not explicitly stated, it was assumed that the patients were from the same country as the senior author’s affiliation. The total number of cases were tallied and tabulated on a world map using OpenStreetMap.

## Results

HC in humans can be caused by or is secondary to several factors including structural brain disorders, cilia abnormalities, brain tumors resulting in CSF obstruction requiring CSF diversion, neural tube defects, prematurity and germinal matrix fragility, neonatal systemic and CNS infections, intracranial hemorrhage, evolutionary selection pressures, and ‘genetic’ anomalies, classically thought as Mendelian disorders (**Figure 1**). Thus, we conducted a systematic review of human genetic studies of HC to quantify and summarize the current state of genetic contributions to HC of various etiologies (**Figure 2**). However, genetic susceptibility confers risk to all these preceding factors as well as to HC directly. Thus, understanding the pleiotropic effect of genes on both risk factors and development of HC is needed and requires highly detailed phenomics analysis [33]. Here, we summarize all genetic studies of *human* HC, including discussion of animal models of HC only as corroborating findings of genes and pathways identified in humans where there is a reasonable degree of evolutionary conservation. We believe this is essential as regulation of CSF and brain development is highly divergent across evolution, necessitating clarification and specificity of how genetics plays a role in human disease. Categories are defined *a priori* based on either phenotypic, molecular, or known genetic classifications. While many forms of HC can reasonably be classified into multiple categories, we attempt to simplify the groupings below.

**Figure 1.**
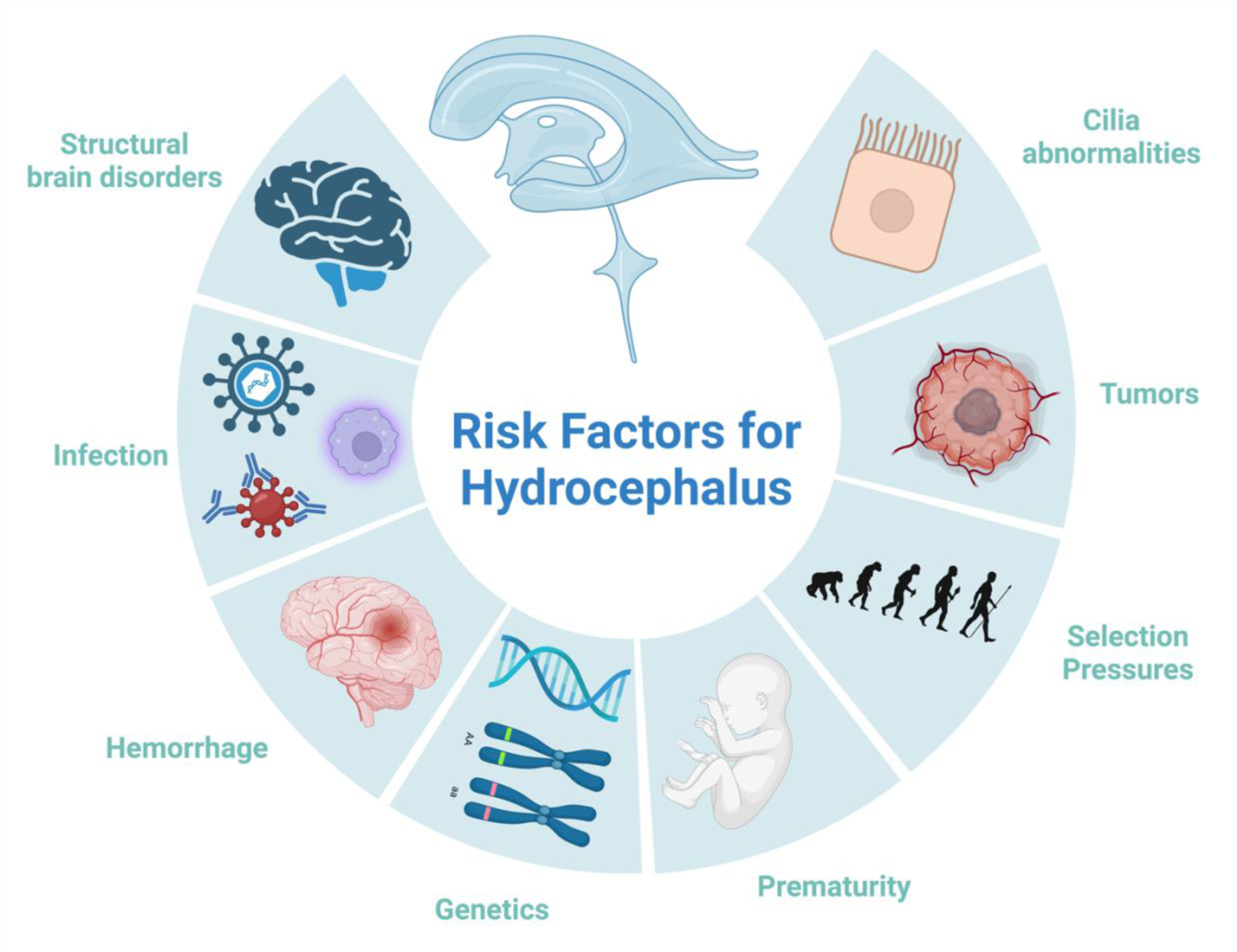
Factors contributing to the development of hydrocephalus in humans.

**Figure 2.**
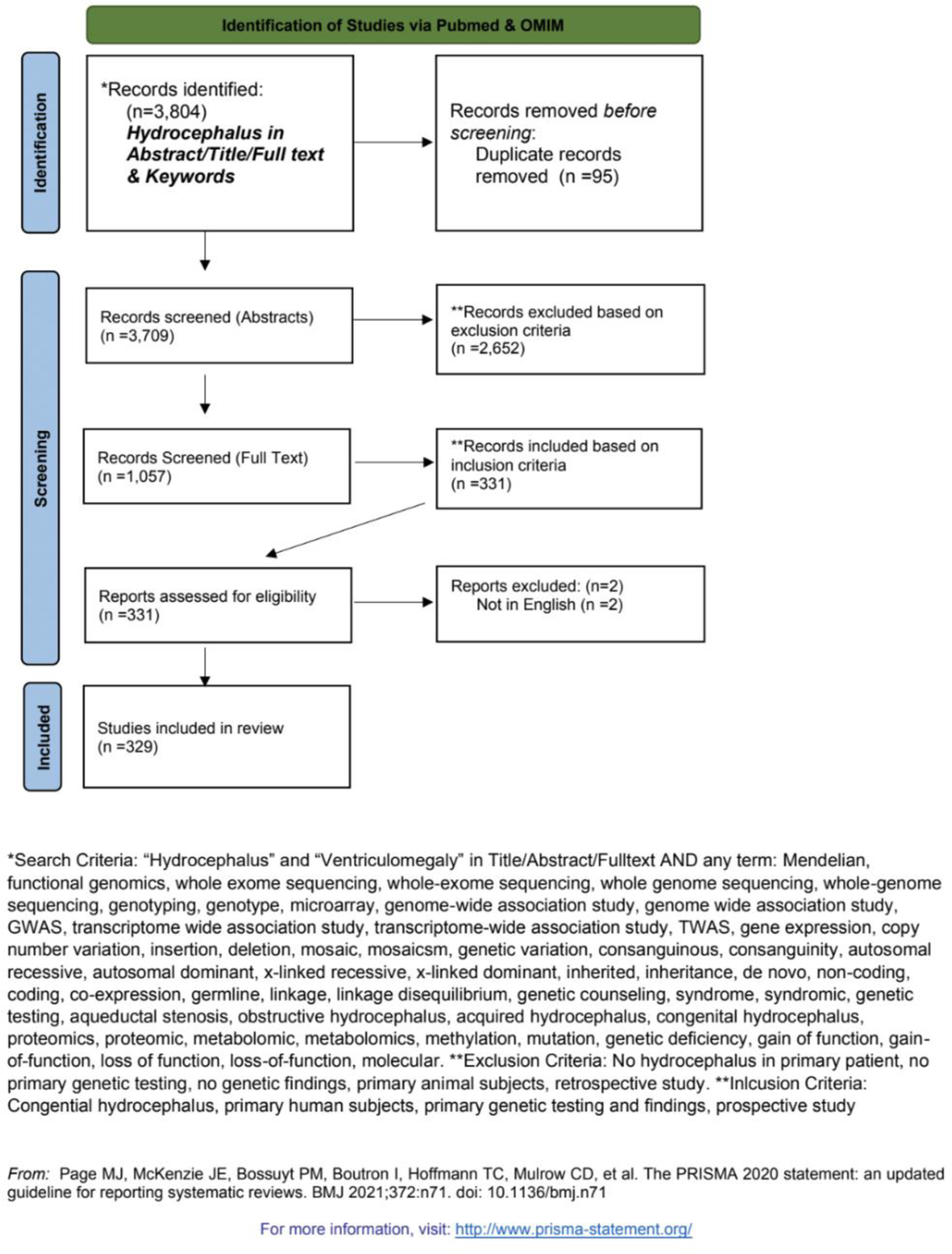
PRISMA flowchart outlining literature search to identify genes, mutations, and genetic mechanisms contributing to hydrocephalus in humans.

### Hydrocephalus secondary to Aqueductal Stenosis (AS)

Human genetics studies of HC secondary to aqueductal stenosis (AS) are summarized in **Table 1**. Fifteen unique gene mutations on 11 chromosomes inherited in both X-linked and autosomal patterns underlying HC secondary to AS have been identified. These genes include protocadherin 9 (*PCDH9),* immunoglobulin superfamily containing leucine rich repeat 2 *(ISLR2),* ATPase Na+/K+ transporting subunit alpha 3 *(ATP1A3),* L1 cell adhesion molecule *(L1CAM),* FA complementation group C *(FAC),* fibroblast growth factor receptor 3 *(FGFR3),* solute carrier family 12 member 6 *(SLC12A6),* crumbs cell polarity complex component 2 *(CRB2),* Bardet-Biedl syndrome 7 *(BBS7),* podocin gene *(NPHS2),* multiple PDZ domain crumbs cell polarity complex component *(MPDZ),* laminin subunit beta 1 *(LAMB1),* alpha glucosidase *(GAA),* A-Disintegrin and Metalloproteinase with Thrombospondin motifs like 2 *(ADAMTSL2),* collagen type IV alpha 2 chain *(COL4A2)*. A duplication in the Xp22.33 region and deletions of the long arm of chromosome 9, 12q22-q23.1, mutation in SRY-box transcription factor 2 (*SOX2*) gene, and mutation in the solute carrier family 12-member 7 (*SLC12A7)* gene were also identified.

**Table 1.** Aqueductal Stenosis.

Abbreviations include the following: Amplification created restriction site (ACRS).
| Citation | Title | Author Affiliation | Case # | Ancestry | Study Design | CNS Phenotype | Non-CNS Phenotype | Type of Hydrocephalus | Genetic Methodology | Genetic Analysis | Inheritance | Genetic Findings |
| --- | --- | --- | --- | --- | --- | --- | --- | --- | --- | --- | --- | --- |
| Alazami et al., 2019 [1] | A novel ISLR2-linked autosomal recessive syndrome of congenital hydrocephalus, arthrogryposis and abdominal distension | King Faisal Specialist Hospital and Research Center, Riyadh, Saudi Arabia | 2 Subjects, 8 Controls | Saudi | Case series | generalized hypotonia, diminished reflexes | arthrogryposis and abdominal distension | Obstructive | WES | Autozygome analysis; Variant analysis | AR | 13q21.32 (c.652T>C, p.Y218H in PCDH9); 15q24.1 (c.1660delT, p.W554Gfs*40 in ISLR2) |
| Allocco et al., 2019 [2] | Recessive Inheritance of Congenital Hydrocephalus with Other Structural Brain Abnormalities Caused by Compound Heterozygous Mutations in ATP1A3 | Yale University, New Haven, CT, United States | 1 Subject, 2 Parents | Caucasian | Case study | open schizencephaly, type 1 Chiari malformation, and dysgenesis of the corpus callosum | - | Obstructive | WES | CNV, Sanger sequencing | AR | 19q13.2 (p.R19C in exon 2 and p.R463C in exon 11 of ATP1A3) |
| Chassaing et al., 2007 [3] | Germinal mosaicism and familial recurrence of a SOX2 mutation with highly variable phenotypic expression extending from AEG syndrome to absence of ocular involvement | Hôpital Purpan, Toulouse, France | 1 Subject, 1 Control | - | Case study | Brain malformations, corpus callosum hypoplasia | ocular dysgenesis, male genital tract malformations, postnatal growth retardation, and facial dysmorphic features | Obstructive | TGS | - | AD | 3q26.33 (deletion within SOX2) |
| Cox et al., 1997 [4] | VACTERL with hydrocephalus in twins due to Fanconi anemia (FA): mutation in the FAC gene | Royal Postgraduate Medical School, Hammersmith Hospital, London, United Kingdom | 2 Subjects, 2 Parents | South African Ashkenazi Jew | Case study | isolated hydrocephalus | absent thumb, pericardial effusion, tracheo-esophageal fistula, pulmonary dysgenesis, intestinal malrotation, ectopic kidney, tetralogy of fallot | Obstructive | TGS | - | AR | 9q22.32 (IVS4 + 4 A to T splice mutation in intron 4 of FAC) |
| De Keersmaecker et al., 2013 [5] | Prenatal diagnosis of MPPH syndrome | University Hospitals, Leuven, Belgium | 1 Subject | - | Case study | polymicrogyria | postaxial polydactyly | Obstructive | Cytogenetics, TGS | aCGH | - | - |
| Escobar et al., 2009 [6] | Significant phenotypic variability of Muenke syndrome in identical twins | St. Vincent Children's Hospital, Indianapolis, Indiana, USA | 2 Subjects | - | Case study | coronal craniosynostosis, porencephalic cyst, and absence of the corpus callosum | Bilateral sensorineural hearing loss, Tracheoesophageal Fistula, ASD, VSD | Obstructive | TGS | - | De novo | 4p16.3 (c.C749G, p.P250R in FGFR3 gene) |
| Gomy et al., 2008 [7] | Two new Brazilian patients with Gómez-López-Hernández syndrome: reviewing the expanded phenotype with molecular insights | School of Medicine of Ribeirão Preto, University of São Paulo, Ribeirão Preto, Brazil | 2 Subjects | Brazilian | Case study | craniosynostosis , craniofacial anomalies, trigeminal anesthesia, cerebellar ataxia, intellectual disability, and rhombencephalo synapsis | Scalp alopecia, developmental delay | Obstructive | TGS | Direct sequencing | - | No pathogenic mutations |
| Isik et al., 2018 (78) [8] | Clinical and genetic features of L1 syndrome patients: Definition of two novel mutations | Faculty of Medicine, Ege University, Izmir, Turkey | 2 Subjects | - | Case series | intellectual disability, microcephaly, spasticity | developmental delay, broad forehead, hypertelorism, low set ears, long philtrum, bilateral adducted thumbs, atrial septal defect | Obstructive | Molecular analysis, unspecified | De-novo mutation analysis, Segregation analysis | X-linked | Xq28 (c.3166 + 1G > A; c.749delG, p.S250Tfs*51) |
| Jin et al., 2019 [9] | SLC12A ion transporter mutations in sporadic and familial human congenital hydrocephalus | Yale University School of Medicine, New Haven, CT, USA | 2 Subjects | - | Case series | agenesis of the corpus callosum, and schizencephaly | - | Obstructive | WES | CNV | *De novo* | 15q14 (c.C1814T, p.Pr05L in SLC12A6); 5p15.33 (SLC12A7 deletion) |
| Jouet et al., 1993 [10] | Refining the genetic location of the gene for X linked hydrocephalus within Xq28 | University of Cambridge | 4 Subjects | - | Case series | seizures, intellectual disability, callosal agenesis, aqueduct stenosis, spastic paraparesis | adducted thumbs | Obstructive | Genotyping | Two point/multi point linkage analysis | X-linked recessive | Xq28 (HSAS gene proximal to DXS605 & coincident with DXS52) |
| Khattab et al., 2011 [11] | A de novo 3.54 Mb deletion of 17q22-q23.1 associated with hydrocephalus: a case report and review of literature | Yale University School of Medicine, New Haven, Connecticut 06520-8064, USA | 1 Subject | - | Case study | generalized hypotonia | sutural separation with full anterior fontanel, small palpebral fissures, hypertelorism, low-set ears, micrognathia, downturned corners of the mouth, arachnodactyly of fingers and toes, contractures of joints | Obstructive | TGS, cytogenetics | aCGH; FISH | *De novo* | 17q22-q23.1 deletion |
| Lamont et al., 2016 [12] | Expansion of phenotype and genotypic data in CRB2-related syndrome | Alberta Children's Hospital Research Institute, University of Calgary, Calgary, Alberta, Canada | 5 Subjects | - | Case series | Hypoplastic cerebellum, subependymal heterotopias, hypertonia | bilateral echogenic kidneys, hypoplastic right lung, hypoplastic right pulmonary artery, dextroposition of the heart, ASD defect, low visual acuity, irregular retinal pigmentation, mild optic atrophy | Obstructive | WES | NGS, Sanger sequencing, SNP genotyping array | AR, *De novo*, maternal | 9q33.3 (CRB2 mutations (p.C620S; p.R628C; p.C629S; p.G1036Afs*42; p.R1248Q; p.W498C; p.R633W; p.E643A; p.W759X; p.N800K; p.D1076A; p.C1098Sfs*53; p.R1115C; p.C1129R)); 4q27 (BBS7 mutation (p.R238Efs*59)); |
|  |  |  |  |  |  |  |  |  |  |  |  | 1q25.2 (NPHS2 mutation (p.R229Q)) |
| Lyonnet et al., 1992 [13] | The gene for X-linked hydrocephalus maps to Xq28, distal to DXS52 | Hôpital des Enfants-Malades, Paris, France | 58 Subjects | French, German | Case series | Intellectual disability and spasticity | abnormal flexion deformity of the thumbs | - | Haplotyping | Pairwise and multipoint linkage analysis | X-linked | HSAS1 localized to Xq28 and distal to DXS52 |
| Maurya et al., 2021 [14] | A case report of Arnold Chiari type 1 malformation in acromesomelic dwarf infant | Seth Gordhandas Sunderdas Medical College and King Edward Memorial Hospital, Mumbai, Maharashtra, India | 1 Subject | - | Case study | Arnold Chiari type 1 malformation, atrophy of both lentiform nuclei, paucity of white matter in bilateral occipital regions, thinning of the corpus callosum | acromesomelic dwarfism | Obstructive | WGS | NGS | AR | 4p16.3 (c.G1138A, p.G380R in exon 9 of FGFR3); 9p23 (c.G394A, p.G132S in exon 5 of MPDZ); 7q31.1 (c.T4133A, p.L1378H in exon 27 of LAMB1); 17q25.3 (c.A1G, p.M1V in exon 2 of GAA) |
| Porayette et al., 2013 [15] | Novel mutations in geleophysic dysplasia type 1 | Boston Children's Hospital, Harvard Medical School, Boston, MA, USA | 1 Subject | - | Case study | Isolated hydrocephalus | large head, prominent flat forehead, hypertelorism, wide mouth with long thin lips, full cheeks, downturned corners of the mouth, short palpebral fissures, long flat philtrum, grooved tongue with tongue-tie, low-set ears, small nose with depressed nasal bridge, short neck, mild abdominal distension, symmetrical shortening of all extremities, aortic stenosis, pulmonary valve stenosis | Obstructive | TGS | - | AD | 9q34.2 (c.1934G>A, p.R645H in exon 13 and mutation in intron 8) of ADAMTSL2) |
| Serville et al., 1993 [16] | Prenatal exclusion of X-linked hydrocephalus-stenosis of the aqueduct of Sylvius sequence using closely linked DNA markers | Unité de Recherches sur les Handicaps Génétiques de l'Enfant INSERM U-12, Hôpital des Enfants-Malades, Paris, France | 2 Subjects | - | Case series | cortex thinning, spasticity, cerebral palsy, intellectual disability, corpus callosum agenesis, aqueductal stenosis | bilateral adducted thumbs | Obstructive | Southern blotting, DNA probes; autoradiography | Linkage analysis | X-linked recessive | Xq28 region linkage with the HSAS locus |
| Strain et al., 1994 [17] | Genetic heterogeneity in X-linked hydrocephalus: linkage to markers within Xq27.3 | Human Genetics Unit, University of Edinburgh, Western General Hospital, UK | 4 Subjects, Controls used | - | Case series | septum pellucidum and corpus callosum agenesis, aqueductal stenosis | adducted thumbs | Obstructive | Chromosomal banding | Two-point/multipoint linkage analysis | - | Linkage to DXS548 and FRAXA loci in Xq27.3 |
| Tzschach et al., 2012 [18] | Interstitial 9q34.11-q34.13 deletion in a | Institute of Human Genetics, | 1 Subject | - | Case study | Intellectual disability | Cleft lip and palate, bilateral talipes equinovarus, | Obstructive | Chromosome analysis | SNP array | - | 9q34.11–q34.13 (3.7 Mb deletion) |
|  | patient with severe intellectual disability, hydrocephalus, and cleft lip/palate | University of Tuebingen, Tuebingen, Germany | , 2 Parents |  |  |  | kyphoscoliosis, psychomotor development delay, short stature, bilateral convergent strabismus, dysmorphic facial features |  |  |  |  |  |
| Verbeek et al., 2012 [19] | COL4A2 mutation associated with familial porencephaly and small-vessel disease | Erasmus University Medical Center, Rotterdam, The Netherlands | 10 Subjects, Controls used | Caucasian , Afghani | Case series | porencephaly, periventricular leukoencephalopathy, cerebellar hypoplasia, cerebral atrophy | developmental delay, feeding difficulties, bilateral ICA, ophthalmological signs | Obstructive | TGS | SNP array | AD | 13q34 (c.4165G4A, p.G1389R in exon 44 and c.3206delC in exon 34 in COL4A2) |
| Vieira et al., 2012 [20] | Primary ciliary dyskinesia and hydrocephalus with aqueductal stenosis | Hospital de Dona Estefânia, Centro Hospitalar de Lisboa Central, Lisbon, Portugal | 1 Subject | Gypsy | Case study | aqueductal stenosis | dextrocardia, a complex heart malformation, situs inversus, intestinal malrotation | Obstructive | TES | - | AR | No mutations in DNAI1 or DNAH5 |

Understanding the function of these genes may confer a mechanistic and phenotypic understanding of HC secondary to AS. For example, some patients with AS will display abnormal brainstem development leading to near complete obliteration of the aqueduct, whereas other children may display relatively normal anatomy associated with a web obscuring CSF flow. Genetics factors contributing to AS include *ATP1A3*, which encodes an ATPase ion channel that has been associated with CNS development and ventricular dilatation when disrupted in zebrafish [34]. In addition, *SLC12A6* codes for the ion transporter KCC3 (K-Cl co transporter) that has been associated with AS among other phenotypes including peripheral neuropathy and agenesis of the corpus callosum in mice [35]. These ion channels are localized to the choroid plexus and are involved in neural stem cell development [36]. *ADAMTSL2*, encoding a glycoprotein, has been shown to interact with fibrillin 1 to enhance transforming growth factor-β (TGFβ) and fibroblast function. Additionally, TGFβ has been implicated in skeletal dysplasia and developmental dysfunction [37]. Thus, it is evident that genes with varying functions may contribute to AS and the diverse co-occurring phenotypes observed in these patients.

### X-linked hydrocephalus

Genes contributing to X-linked HC include apoptosis inducing factor mitochondria associated 1 (*AIFM1),* adaptor related protein complex 1 subunit sigma 2 *(AP1S2),* EBP cholestenol delta-isomerase *(EBP),* FA complementation group B *(FANCB),* histone deacetylase 6 *(HDAC6),* OFD1 centriole and centriolar satellite protein *(OFD1),* OTU deubiquitinase 5 *(OTUD5),* coiled-coil domain containing 22 (*CCDC22*), and porcupine O-acyltransferase *(PORCN).* **Table 2** summarizes the genetic studies of X-linked HC in humans. *AIFM1* is involved in regulation of apoptosis [38]. In addition, *AP1S2* regulates endosomal protein trafficking and structural integrity [39]. HDAC6 has been shown to interact with Runx2, a transcription factor involved in osteoblast differentiation, and other HDACs exhibit high expression patterns in prehypertrophic chondrocytes, indicating their role in endochondral ossification and skeletal dysplasias [40]. OTUD5 mutations also impact transcriptional regulation with its inability to prevent HDAC degradation and maintain neural stem cell development [41]. OFD1 and PORCN mutations affect signaling pathways such as hedgehog signaling or wingless/integrated (Wnt) signaling [42; 43].

**Table 2:**
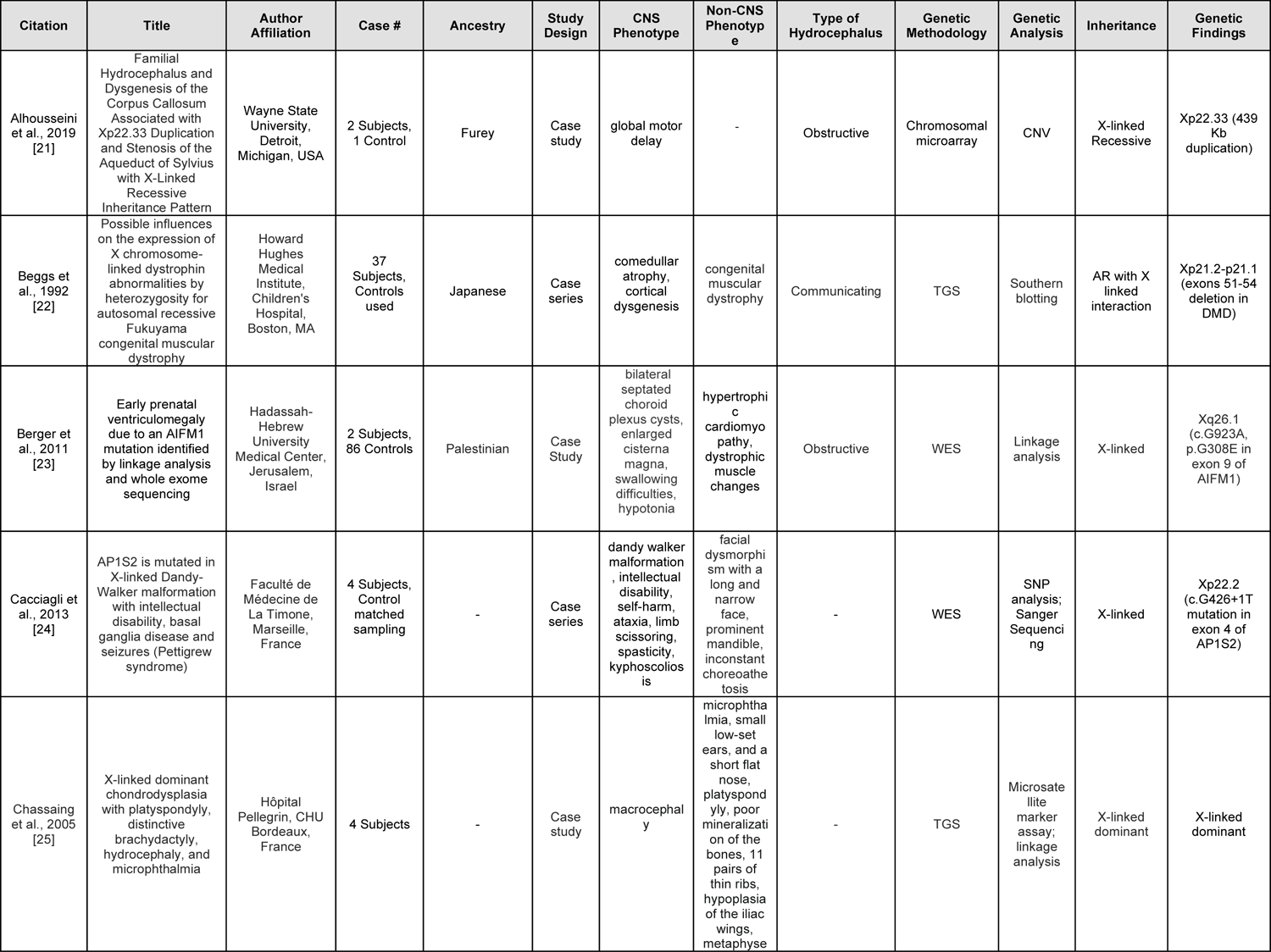
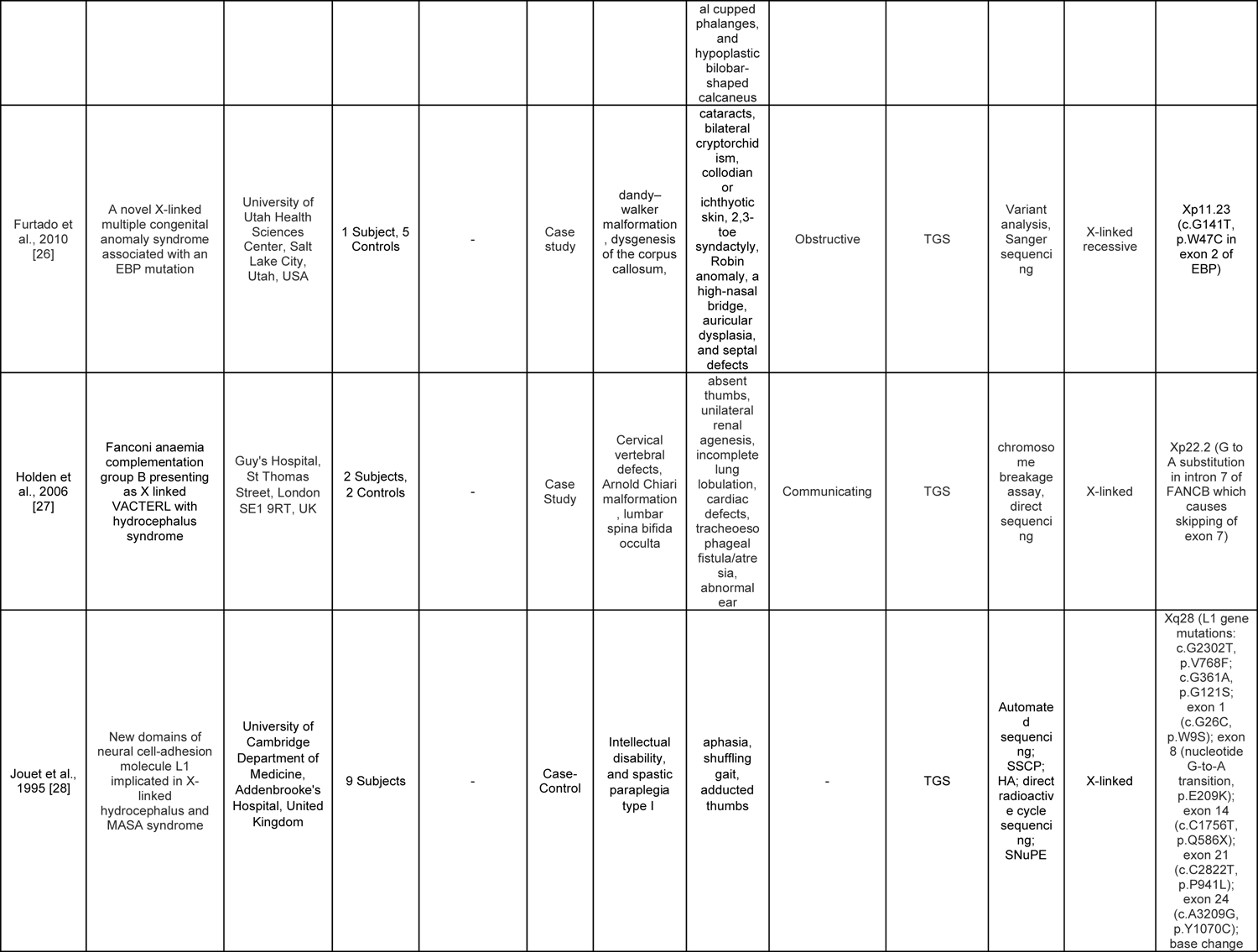
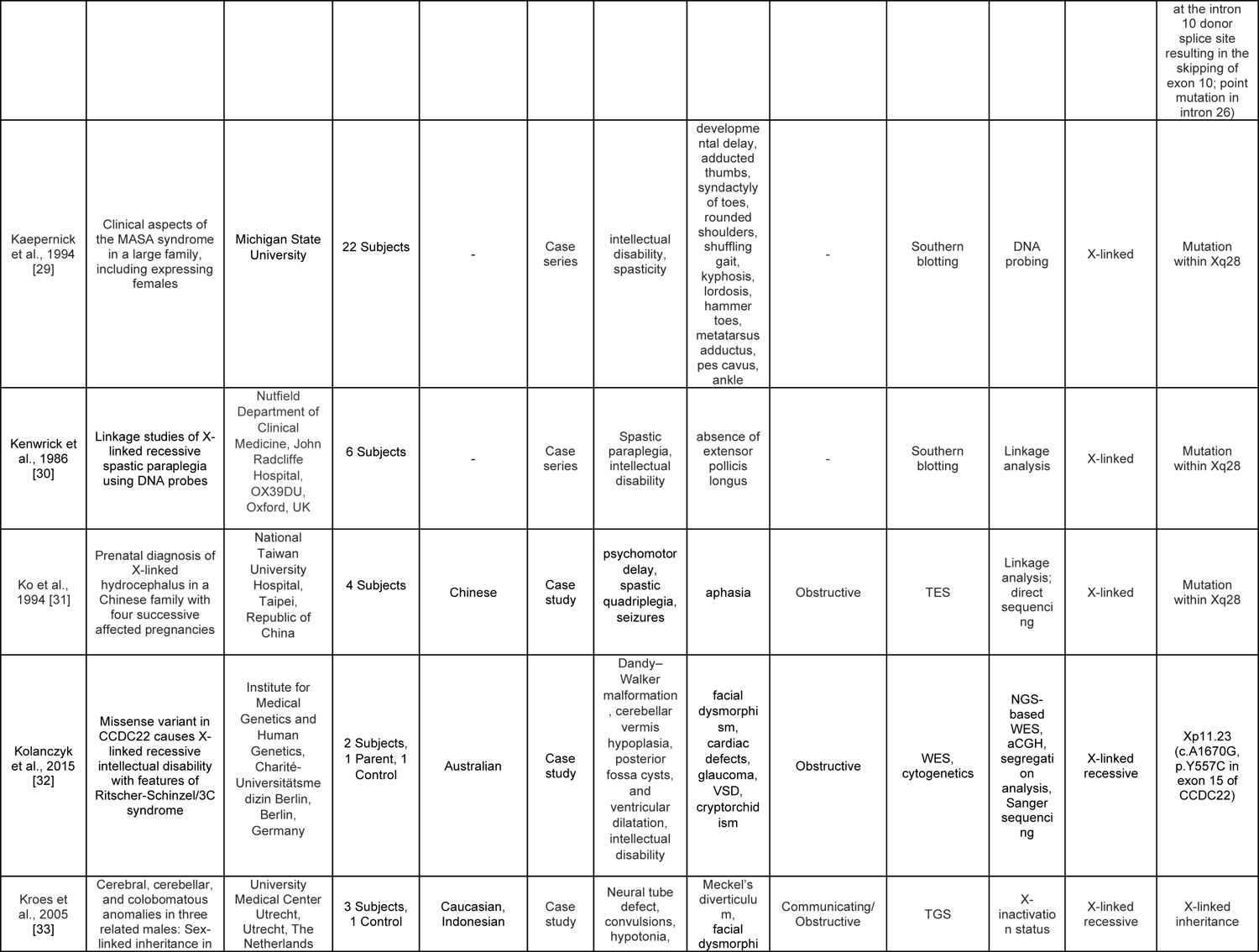
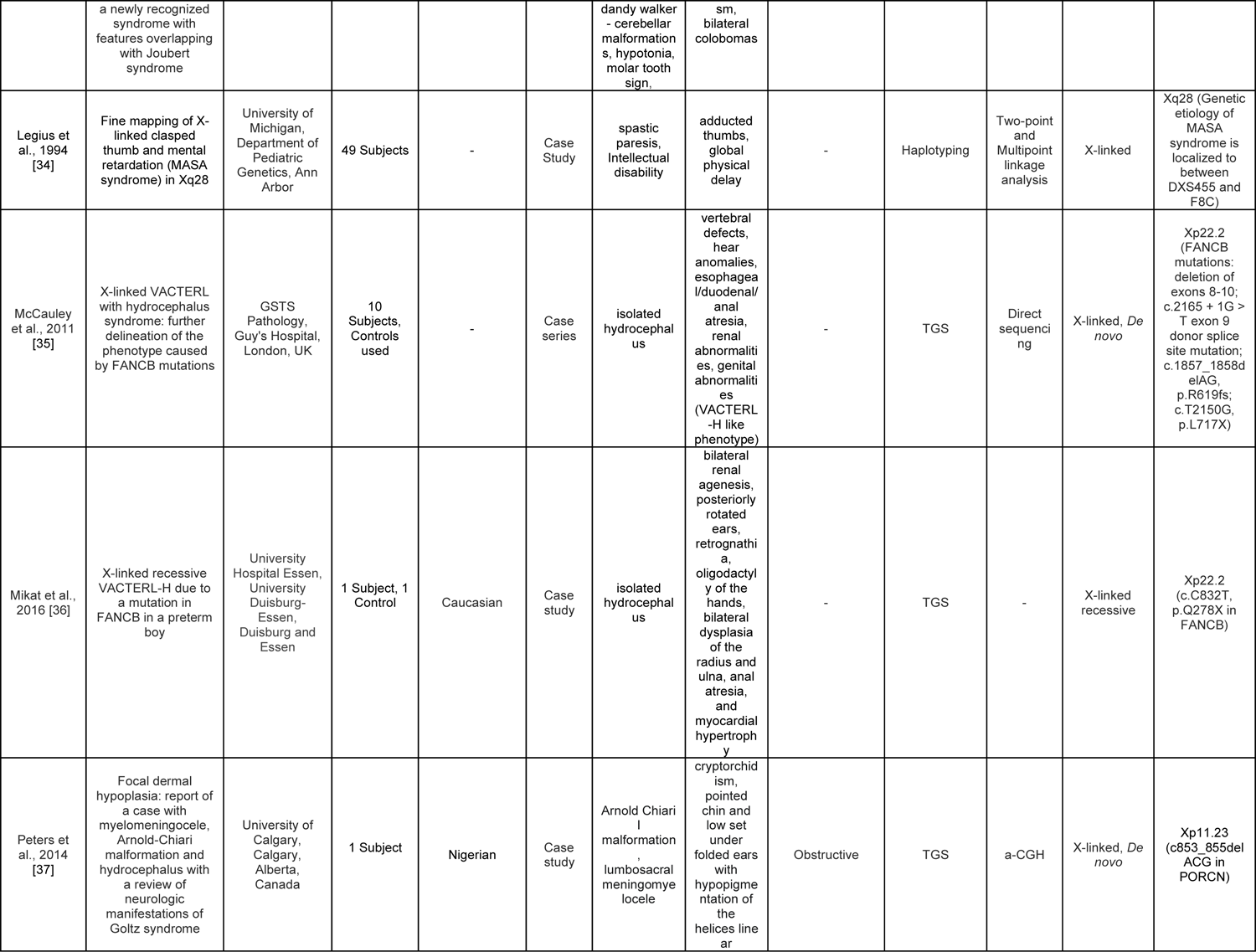
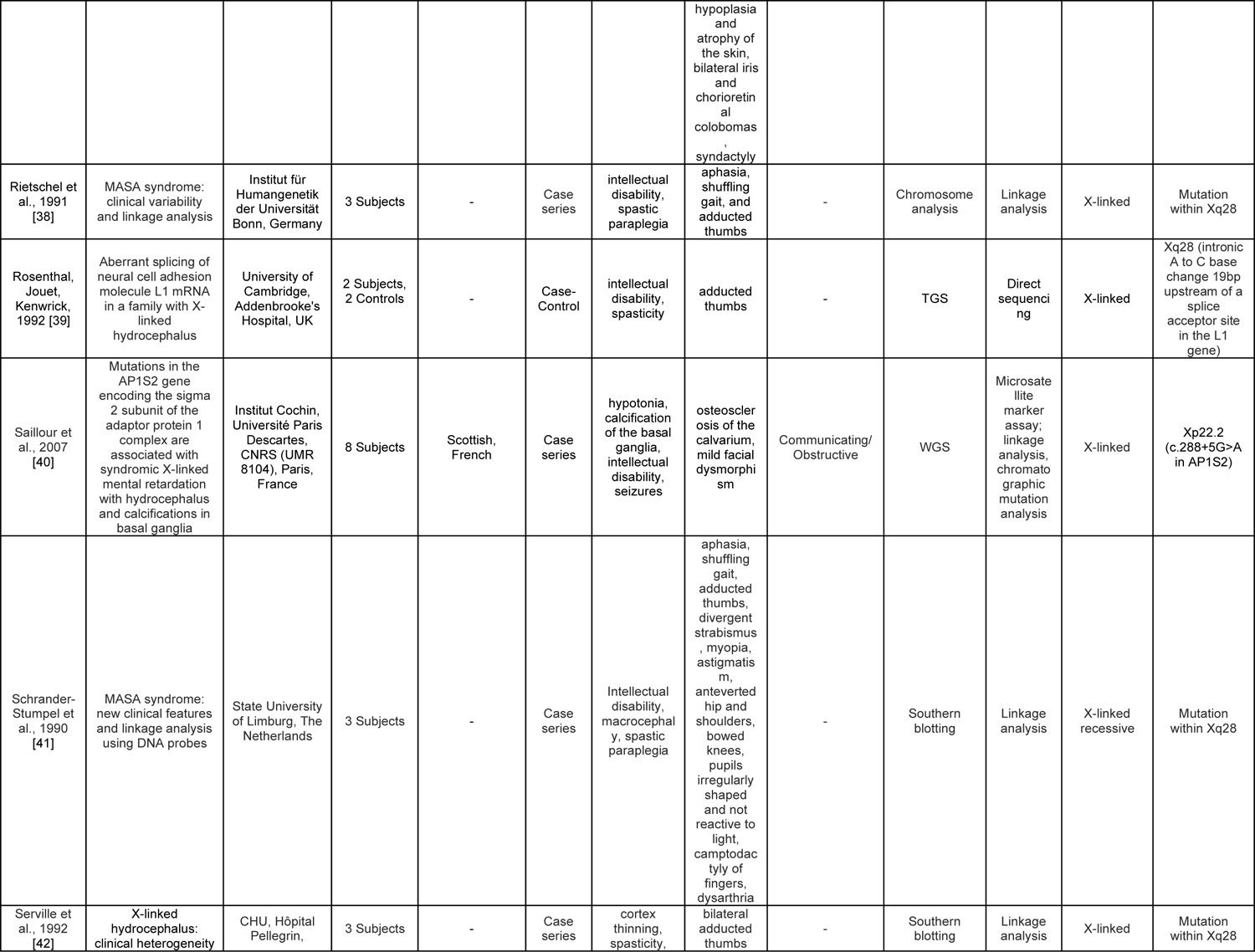
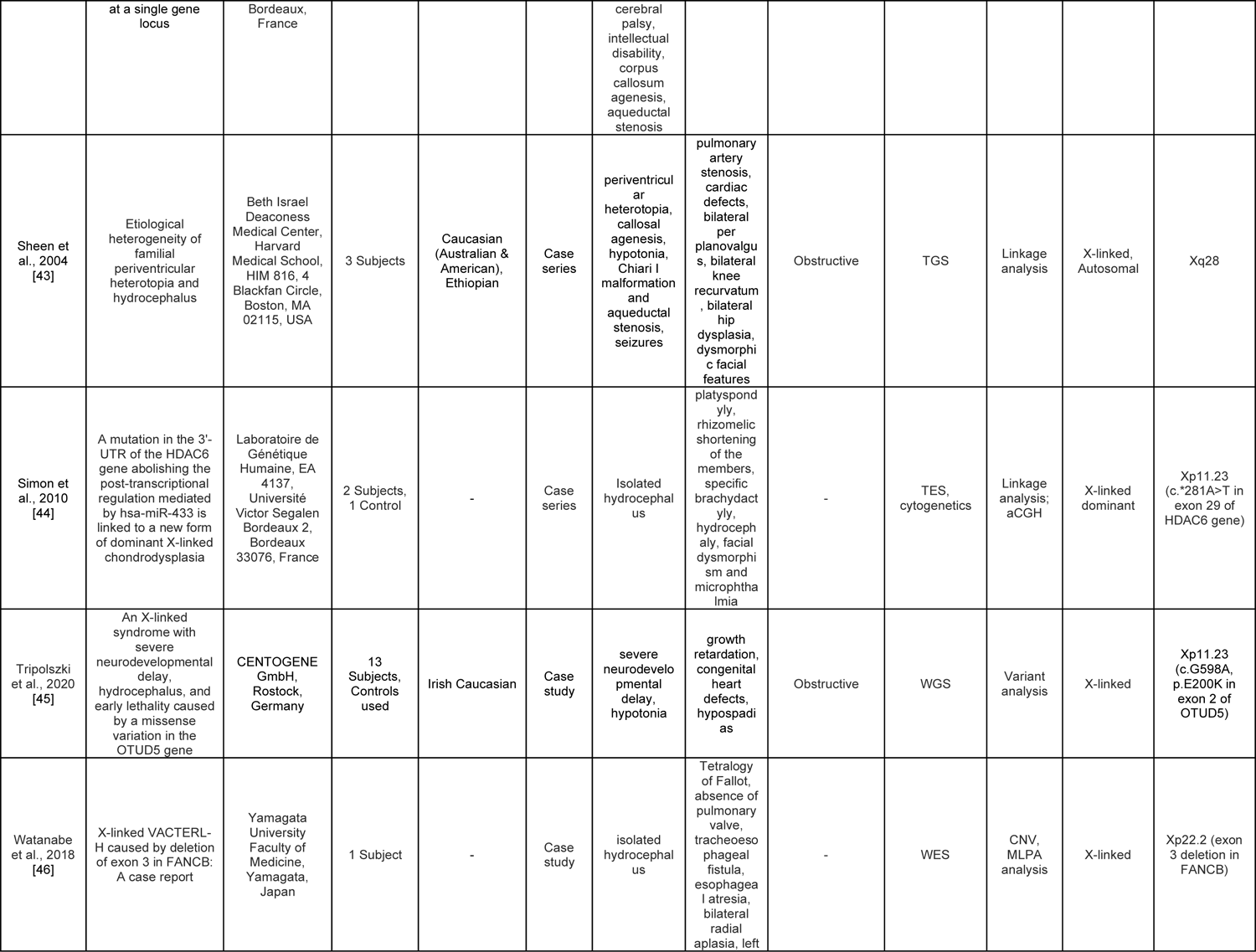
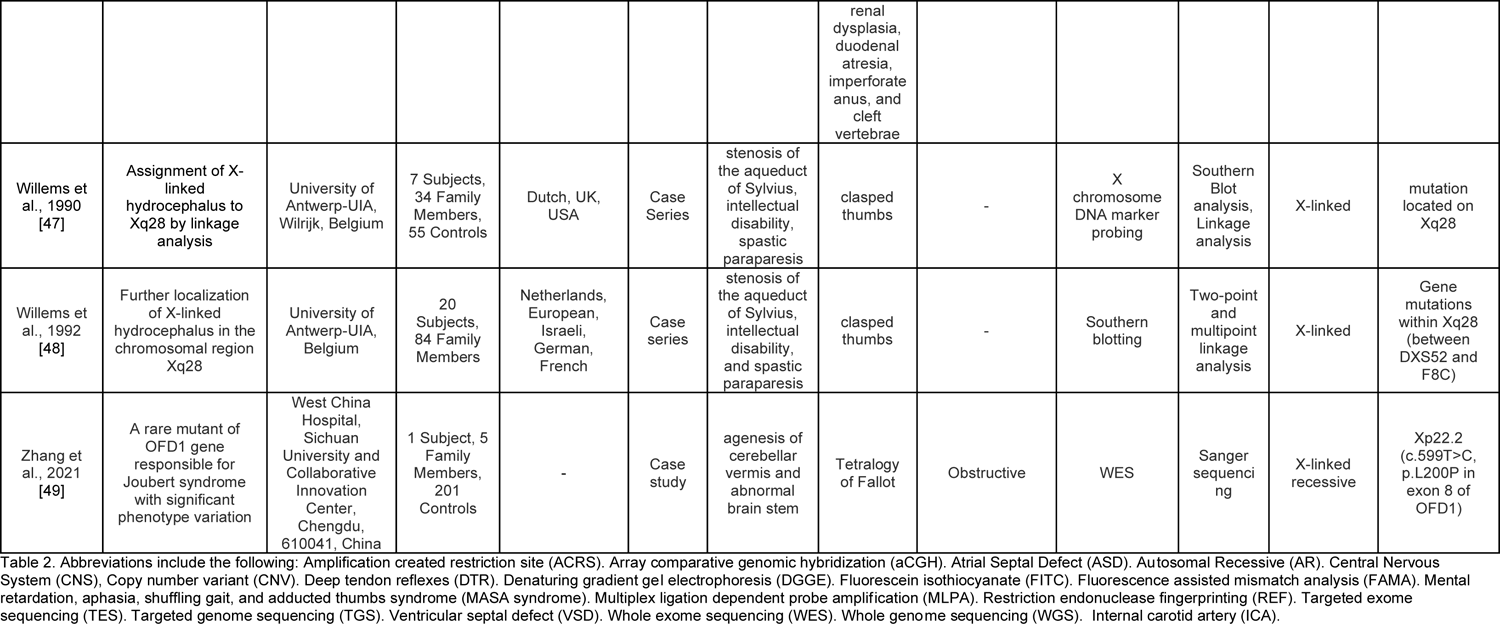
X-linked Hydrocephalus.

### L1CAM associated hydrocephalus

Next, we discuss L1CAM associated HC, as this entity is well described and distinct phenotypically. Early linkage analysis studies of HC identified a mutation within the long arm of chromosome X, specifically Xq28. Further genomic analyses localized to a region between the gene loci of *DXS52* and *F8C*, within which L1 cell adhesion molecule (*L1CAM)* resides. The genetic understanding of X-linked HC has primarily been linked to genetic alterations at the *L1CAM* locus. *L1CAM* duplications include the 3’ end of the open reading frame and exons 2-10. *L1CAM* insertions include exon 18 and the junction sequence between *L1CAM* and *AVPR2*. *L1CAM* deletions/microdeletions include exons 2, 5-8, 10, 11, 18, 19, 21-23, 26, intron 18, and whole gene deletion. *L1CAM* missense mutations include exons 1-16, 18, 20, 21, 24, 27, 28 and introns 2-4, 6-8, 10-15, 18, 21, 22, 24, and 26. *L1CA*M nonsense mutations include exons 1, 3, 8, 10-14, and 20-22. A silent mutation in Exon 8 of *L1CAM* has been associated with HC. A summary of all mutations across L1CAM can be found in **Figure 3** and **Table 3**. Mutations in L1CAM are also associated with MASA syndrome (characterized by mental retardation, aphasia, shuffling gait, and adducted thumb), and spastic paraplegia, highlighting the pleiotropic role of L1CAM in human disease.

**Figure 3.**
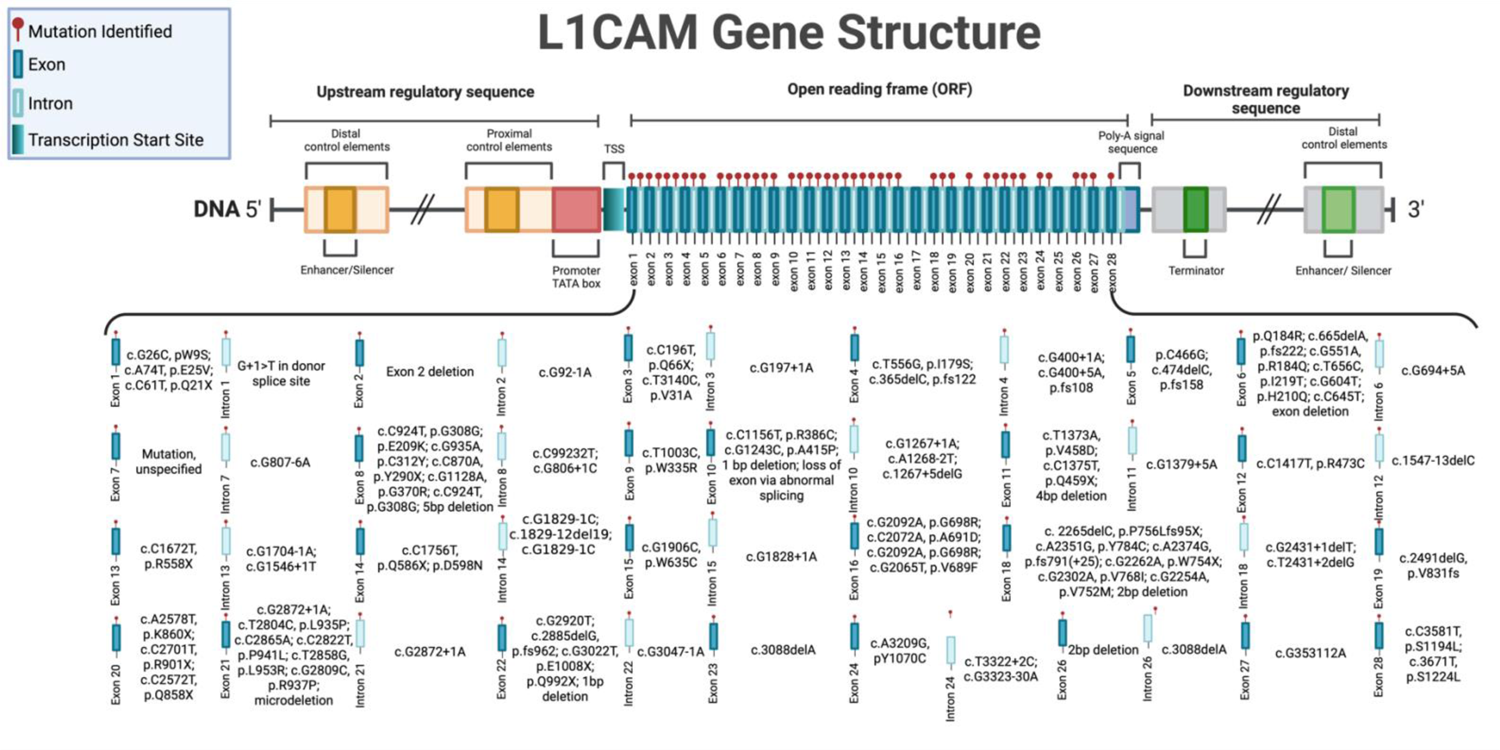
L1CAM mutations implicated in human patients with hydrocephalus.

**Figure 4.**
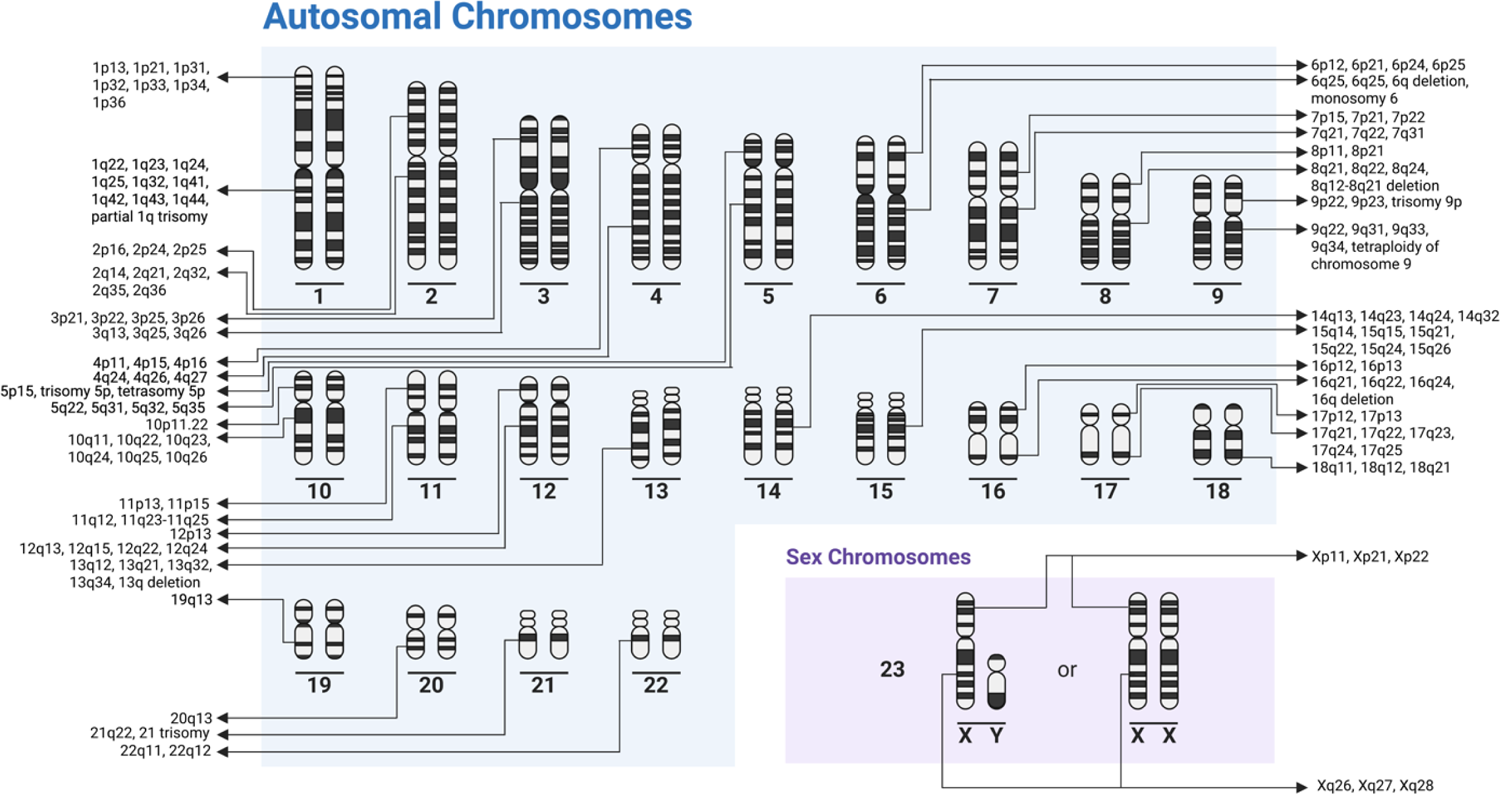
Chromosome map of hydrocephalus-associated loci across autosomal and sex chromosomes in humans.

**Table 3:** L1CAM.

Abbreviations include the following: Amplification created restriction site (ACRS).
| Citation | Title | Author Affiliation | Case # | Ancestry | Study Design | CNS Phenotype | Non-CNS Phenotype | Type of Hydrocephalus | Genetic Methodology | Genetic Analysis | Inheritance | Genetic Findings |
| --- | --- | --- | --- | --- | --- | --- | --- | --- | --- | --- | --- | --- |
| Bott et al., 2004 [50] | Congenital idiopathic intestinal pseudo-obstruction and hydrocephalus with stenosis of the aqueduct of sylvius | Jeanne de Flandre Hospital, Faculty of Medicine Lille, France | 1 Subject | French | Case study | bilateral nystagmus, convergent strabismus, spastic paraplegia, callosal agenesis | bilateral adducted thumbs, abdominal distension | Obstructive | - | - | X-linked recessive | Xq28 (G2920T in exon 22 of L1CAM) |
| Brewer et al., 1996 [51] | X-linked hydrocephalus masquerading as spina bifida and destructive porencephaly in successive generations in one family | Western General Hospital, Edinburgh, UK | 1 Subject | - | Case study | midline cysts, callosal agenesis, cognitive impairment | asymmetric tetraplegia, low vision, eye movement disorder | Obstructive | - | - | X-linked recessive | Xq28 (Frameshift mutation in L1CAM) |
| Chidsey et al., 2014 [52] | L1CAM whole gene deletion in a child with L1 syndrome | ARUP Laboratories, Salt Lake City, Utah | 1 Subject, 1 Parent | North European | Case study | absent septum pellucidum | adducted thumbs with contractures | Obstructive | microarray analysis | SNP analysis | X-linked | Xq28 (62 kb deletion) |
| Claes et al., 1998 [53] | Hydrocephalus and spastic paraplegia result from a donor splice site mutation (2872 + 1G to A) in the L1CAM gene in a Venezuelan pedigree | Center for Human Genetics, University of Leuven, Belgium | 3 Subjects | Venezuela | Case study | aqueductal stenosis, psychomotor delay, hypotonia, spastic paraplegia | Learning difficulties | Obstructive | cDNA analysis; TGS | Solid-phase approach w/ FITC primer | X-linked | Xq28 (exon 21 microdeletion in L1CAM; G-to-A transition at bp 2872 + 1 of exon 21) |
| Coucke et al., 1994 [54] | Identification of a 5' splice site mutation in intron 4 of the L1CAM gene in an X-linked hydrocephalus family | University of Antwerp-UIA, Belgium | 1 Subject, 1 Control | - | Case study | aqueductal stenosis, intellectual disability, spastic paresis | clapsed thumbs | - | RT-PCR | Linkage analysis | X-linked | Xq28 (G to A transition at position -5 of the 5' splice site of intron 4 of L1CAM) |
| Du et al., 1998 [55] | A silent mutation, C924T (G308G), in the Li CAM gene results in X linked hydrocephalus (HSAS) | J C Self Research Institute of Human Genetics, Greenwood Genetic Center, SC 29646, USA | 1 Subject, 1 Family member | - | Case study | callosal agenesis, intellectual disability, spastic paraplegia | clenched fingers, overlapping digits | Obstructive | TES | REF | X-linked | Xq28 (c.C924T, p.G308G silent mutation in exon 8 of L1CAM) |
| Du et al., 1998 [56] | Multiple exon screening using restriction endonuclease fingerprinting (REF): detection of six novel mutations in the L1 cell adhesion molecule (L1CAM) gene | J C Self Research Institute of Human Genetics, Greenwood Genetic Center, SC 29646, USA | 5 Subjects, 200 Controls | - | Case series | Intellectual disability | adducted thumbs with contractures , aphasia | - | TGS | SSCP, REF | X-linked, *De novo* | Xq28 (6 novel mutations in the L1CAM gene: intron 7 (G to A base substitution at position 807-6 at the 3' splice site); intron 11 (G to A transition at position 1379+5 within the 5' splice site); intron 10 (A to T base change at position 1268-2 of the 3' splice site); within exons |
|  |  |  |  |  |  |  |  |  |  |  |  | 16-18 (c.G2092A, p.G698R); exon 16 (c.C2072A, p.A691D); exon 21 (c.T2804C, p.L935P)) |
| Du et al., 1998 [57] | Somatic and germ line mosaicism and mutation origin for a mutation in the L1 gene in a family with X-linked hydrocephalus | J C Self Research Institute of Human Genetics, Greenwood Genetic Center, SC 29646, USA | 5 Subjects, 200 Controls | - | Case study | Intellectual disability | adducted thumbs with contractures , aphasia | Obstructive | TGS | SSCP | X-linked | Xq28 (G to A nucleotide change at the first position of intron 10 of L1CAM) |
| Ferese et al., 2016 [58] | A New Splicing Mutation in the L1CAM Gene Responsible for X-Linked Hydrocephalus (HSAS) | Localita' Camerelle, Pozzilli, Italy | 1 Subject, 1 Control | - | Case study | aqueductal stenosis, thinned cerebral parenchyma, lissencephaly, corpus callosum agenesis, | adducted thumbs, dysmorphic facial features | Obstructive | TGS | Direct sequencing | X-linked recessive | Xq28 (Intron 10 in L1CAM hemizygous for c.1267+5delG; loss of exon 10 via abnormal splicing) |
| Fernandez et al., 2012 ([59] | Association of X-linked hydrocephalus and Hirschsprung disease: report of a new patient with a mutation in the L1CAM gene | Instituto de Biomedicina de Sevilla (IBIS), Hospital Universitario Virgen del Rocío/CSIC/Universidad de Sevilla, Sevilla, Spain. | 1 Subject | - | Case study | bilateral spastic tetraparesis, psychomotor growth delay | cephalo-pelvic dystocia, Hirschsprung's disease | Obstructive | TES | - | X-linked | Xq28 (c.2092G>A, p.G698R in exon 16 of L1CAM) |
| Finckh et al., 2000 [60] | Spectrum and Detection Rate of L1CAM Mutations in Isolated and Familial Cases with Clinically Suspected L1-Disease | University Hospital Eppendorf, University of Hamburg, Hamburg, Germany. | 153 Subjects, 100 controls | - | Case series | spastic paraplegia, intellectual disability, agenesis, hypoplasia of corpus callosum | adducted thumbs with contractures , cleft palate, heart malformation, esophageal atresia, club feet | - | TES | SSCP | X-linked, *De novo* | Xq28 (L1CAM mutations) |
| Fransen et al., 1994 [61] | X-linked hydrocephalus and MASA syndrome present in one family are due to a single missense mutation in exon 28 of the L1CAM gene | University of Antwerp-UIA, Belgium | 1 Subject | - | Case study | extreme macrocephaly, severe spasticity, and intellectual disability | shuffling gait, adducted thumbs | - | Southern Blotting | SSCP analysis | X-linked | Xq28 (c.C3581T, p. S1194L in exon 28 of L1CAM) |
| Gigarel et al., 2004 [62] | Single cell co-amplification of polymorphic markers for the indirect preimplantation genetic diagnosis of hemophilia A, X-linked adrenoleukodystrophy, X-linked hydrocephalus and incontinentia pigmenti loci on Xq28 | Hôpital Necker Enfants Malades, 75015 Paris, France | 10 Subjects, Controls used | - | Case Series | isolated hydrocephalus | Hemophilia A | - | TGS | Microsatellite marker assay | X-linked recessive | Xq28 (2872+1 G to A mutation in intron 21 of L1CAM) |
| Graf et al., 2000 [63] | Diffusion-weighted magnetic resonance imaging in boys with neural cell adhesion molecule L1 mutations and congenital hydrocephalus | University of Washington School of Medicine, Seattle, USA | 5 Subjects, 1 Control | Italian | Case series | agenesis of the corpus callosum, diffuse cerebral dysplasia, decreased white matter, small posterior fossa, Chiari I malformation | developmental delay | Obstructive | TES | REF | X-linked | Xq28 (14bp deletion in exon 11, 1bp deletion in exon 10, p.C466G in exon 5, and p.R184W in L1CAM) |
| Gregory et al., 2019 [64] | Mutations in MAGEL2 and L1CAM Are Associated with Congenital Hypopituitarism and Arthrogryposis | UCL Great Ormond Street Institute of Child Health, London, United Kingdom | 5 subjects, Controls used | European, Chile, Afro-Caribbean | Case series | Hypotonia, bulky tectum, white matter loss, thin corpus callosum | bilateral radial clubbed hands, plagiocephaly, distal arthrogryposis with adducted thumbs, flexion deformities, growth hormone deficiency, ventricular septal defect, severe obstructive sleep apnea, global developmental delay, right hip subluxation, scoliosis, bilateral astigmatism, visual impairment | Communicating | WES; chromosome microarray | Sanger Sequencing; Human embryonic expression analysis; Ingenuity Variant analysis | X-linked | Xq28 (c.G1354A, p.G452R in L1CAM) |
| Griseri et al., 2009 [65] | Complex pathogenesis of Hirschsprung's disease in a patient with hydrocephalus, vesico-ureteral reflux and a balanced translocation t(3;17)(p12;q11) | Laboratory Molecular Genetics and Cytogenetics, Genova, Italy | 1 Subject, 1 Control | - | Case Study | intellectual disability, bilateral spastic paraplegia | adducted thumbs, vesico-ureteral reflux, developmental delay | - | TGS | - | X-linked, *De novo* | Xq28 (c.2265delC, p.P756Lfs95X in exon 18 of L1CAM); haploinsufficiency of MYO18A/TIAF1 genes involved in a balanced translocation (3;17)(p12;q21) |
| Gu et al., 1996 [66] | Five novel mutations in the L1CAM gene in families with X linked hydrocephalus | Institut für Humangenetik, Medizinische Universität zu Lubeck, Germany | 5 Subjects | - | Case series | intellectual disability, spastic paresis, complex brain malformation with agenesis of the corpus callosum and fusion of the thalamus | Deafness, adducted thumbs, global physical delay, cleft lip and palate | - | TES | SSCP, HA | X-linked, *De novo* | Xq28 (mutations in exon 1, 6, 7, and 8 of L1CAM) |
| Guo et al., 2020 [67] | A novel nonsense mutation in the L1CAM gene responsible for X-linked congenital hydrocephalus | Xiangya Hospital, Central South University, Changsha, Hunan, China | 1 Subject | Chinese | Case study | agenesis of the corpus callosum, vermis hypoplasia and enlargement of the quadrigeminal plate, aqueductal stenosis | tower-shaped skull, contractions of both thumbs | Obstructive | WES; chromosomal karyotyping ; chromosomal microarray analysis | Sanger Sequencing; Variant segregation analysis | X-linked recessive | Xq28 (c.C2865A in exon 21 of L1CAM) |
| Hubner et al., 2004 [68] | Intronic mutations in the L1CAM gene may cause X-linked hydrocephalus by aberrant splicing | University Hospital Eppendorf, Hamburg, Germany | 7 Subjects, 50 Controls | - | Case series | intellectual disability, hypoplastic or absent corticospinal tract, callosal agenesis, spastic paraparesis, aqueductal stenosis | aphasia, shuffling gait, adducted thumbs | - | TGS | SSCP analysis | X-linked | Xq28 (Intronic L1CAM sequence variants: c.523+5G>A; c.1123+1G>A; c.1547-13delC in intron 12; c.3323-17dupG; c.3457+3A>T; c.3457+18C>T; and c.523+12C>T) |
| Jouet et al., 1994 (79) | X-linked spastic paraplegia (SPG1), MASA syndrome and X-linked hydrocephalus result from mutations in the L1 gene | University of Cambridge, Addenbrooke's Hospital, UK | 6 Subjects | - | Case series | agenesis of the corpus callosum; agenesis of the septum pellucidum; fusion of thalami and hypoplasia of the corticospinal tract, aqueductal stenosis, spastic diplegia | adducted thumbs | Obstructive | TES | SSCP, HA | X-linked | Xq28 (L1 gene mutations: 2 bp deletion in exon 26; single nucleotide deletion in exon 22; p.H210Q in second Ig domain; G to A nucleotide change, p.Q184R in exon 6; C to T mutation in exon 12 that introduces a stop codon at amino acid position 485; G to A mutation in exon 11 that changes a Gly to Arg residue) |
| Jouet et al., 1995 [69] | Gene analysis of L1 neural cell adhesion molecule in prenatal diagnosis of hydrocephalus | Addenbrooke's Hospital, Cambridge, UK | 2 Subjects | - | Case series | Intellectual disability, spastic paraparesis | developmental delay, adducted thumbs | - | TGS | SSCP, Direct sequencing | X-linked, *De novo* | Xq28 (g+ 1->t in the intron 1 donor splice site and 1bp deletion in exon 22 of L1CAM) |
| Jouet et al., 1996 [70] | Discordant segregation of Xq28 markers and a mutation in the L1 gene in a family with X linked hydrocephalus | University of Cambridge, UK | 19 Subjects | - | Case study | Intellectual disability, and spastic paraplegia type I | aphasia, shuffling gait, adducted thumbs | - | TES | SSCP; HA | X-linked | Xq28 (deletion of a single adenosine at position 3088 in exon 23 of L1CAM) |
| Kanemura et al., 2006 [71] | Molecular mechanisms and neuroimaging criteria for severe L1 syndrome with X-linked hydrocephalus | Osaka National Hospital, Osaka, Japan | 96 Subjects, 7 Controls | Japanese | Case series | corpus callosum agenesis, vermis hypoplasia, epilepsy, spastic paraplegia | bilateral adducted thumbs, developmental delay, elevated diaphragm | - | TGS | Direct sequencing | X-linked, *De novo* | Xq28 (L1CAM mutations: exon 1 (c.A74T, p.E25V); exon 5 (c.474delC, p.fs158); exon 6 (c.665delA, p.fs222); exon 8 (c.G935A, p.C312Y; c.C870A, p.Y290X); exon 11 (c.T1373A, p.V458D); exon 16 (c.G2065T, p.V689F); exon 18 (c.G2254A, p.V752M); exon 20 (c.A2578T, p.K860X; c.C2701T, p.R901X); exon 21 (c.T2858G, p.L953R); exon 22 (c.2885delG, p.fs962; c.G3022T, p.E1008X); intron 2 (c.92-1gA); intron 3 (c.197+1gA); intron 4 (400+1gA); intron 6 (c.694+5gA); intron 13 (c.1704-1gA); intron 14 (c.1829-1gC; c.1829-12del19bp); intron 15 (c.1940-21~1940-6); intron 18 (c.2431+1delGT); intron 21 (c.2872+1gA); intron 22 (c.3047-1gA)) |
| Kong et al., 2019 [72] | A new frameshift mutation in L1CAM producing X-linked hydrocephalus | Sichuan Provincial Hospital for Women and Children, Chengdu, China | 1 Subject, 2 Parents | - | Case study | callosal agenesis and lissencephaly | - | - | WES | Sanger Sequencing | X-linked recessive | Xq28 (c.2491delG (p.V831fs) in exon 19 of L1CAM) |
| Liebau et al., 2007 [73] | L1CAM mutation in a boy with hydrocephalus and duplex kidneys | University Hospital of Freiburg, Mathildenstrasse 1, Freiburg, Germany | 1 Subject | - | Case study | tower-shaped skull, corpus callosum agenesis, Intellectual disability, microcephaly, strabismus, neurogenic bladder dysfunction, spasticity | bilateral duplex kidneys and ureters, unilateral mega-ureter, adducted thumbs | - | TGS | SSCP | X-linked | Xq28 (c.2431+2delTG at the beginning of intron 18 of L1CAM) |
| Limbrick et al., 2017 [74] | Cerebrospinal fluid biomarkers of infantile congenital hydrocephalus | Washington University in St. Louis, School of Medicine, Saint Louis, MO, United States of America | 20 Subjects, 51 Controls | Caucasian , Black, Asian | Case series | Isolated hydrocephalus | - | Obstructive | Chromosomal microarrays , TGS | - | - | Xq28 (G847X mutation in L1CAM); 1q25.2 anomaly; 11q24.2 anomaly |
| MacFarlane et al., 1997 [75] | Nine novel L1 CAM mutations in families with X linked hydrocephalus | University of Cambridge Department of Medicine, Addenbrooke's Hospital, Cambridge, UK | 20 Subjects, 56 Controls | - | Case series | intellectual disability, spastic paraplegia, corpus callosum agenesis, absence of the cortical spinal tract | adducted thumbs | Obstructive | TES | SSCP; SNuPE | X-linked | Xq28 (L1CAM: exon 6 (c.G551A, p.R184Q); exon 11 (microdeletion); exon 13 (c.C1672T, p.R558X); exon 18 (c.A2351G, p.Y784C; c.A2374GG, p.fs791(+25); c.G2262A, p.W754X); exon 20 (c.C2701T, p.R901X); exon 21 (microdeletion); intron 7 (c.G(807-6)A); intron 24 (c.T(3322+2)C)) |
| Marin et al., 2015 [76] | Three cases with L1 syndrome and two novel mutations in the L1CAM gene | Hospital Universitario Puerta del Mar, Cádiz, Spain | 3 subjects | - | Case series | corpus callosum agenesis, microcephaly, spastic paraplegia, | Developmental delay, bilaterally flexed adducted thumbs, bilateral clinodactyly of the fifth finger | - | TES | Sanger Sequencing | X-Linked recessive, *De novo* | Xq28 (L1CAM mutations: c.A1754C, p.D585A; c.C3478T, p.Q1160X; c.G353112A in exon 27) |
| Marx et al., 2012 [77] | Pathomechanistic characterization of two exonic L1CAM variants located in trans in an obligate carrier of X-linked hydrocephalus | Institute of Anatomy and Cell Biology, Center for Neurosciences, University of Freiburg, Freiburg, Germany | 3 Subjects, Control cells used | - | Case study | aqueductal stenosis | Adducted thumbs | Obstructive | TGS | Direct sequencing | X-linked | Xq28 (L1CAM mutations: c.C99232T in intron 8; c.G1906C, p.W635C in exon 15; c.G2302A, p.V768I in exon 18) |
| Michaelis et al., 1998 [78] | The site of a missense mutation in the extracellular Ig or FN domains of L1CAM influences infant mortality and the severity of X linked hydrocephalus | Center for Molecular Studies, J C Self Research Institute, Greenwood Genetic Center, SC 29646, USA | 7 Subjects, Controls used | - | Case series | intellectual disability, spasticity, aqueductal stenosis | adducted thumbs | Obstructive | TES | SSCP, REF | X-linked | Xq28 (missense mutations in the extracellular Ig or FN domains of L1CAM) |
| Nakakimura et al., 2008 [79] | Hirschsprung's disease, acrocallosal syndrome, and congenital hydrocephalus: report of 2 patients and literature review | Hokkaido University Graduate School of Medicine, Sapporo, Japan | 2 Subjects | - | Case series | callosal body agenesis, spastic paralysis, and porencephaly | bilateral inferior limbs, and bilateral thumb adduction, polydactyly | Obstructive | TGS | Direct sequencing | X-linked | Xq28 (c.T3140C, p.V31A in exon 3 of L1CAM) |
| Okamoto et al., 1997 [80] | Hydrocephalus and Hirschsprung's disease in a patient with a mutation of L1CAM | Osaka Medical Centre, Japan | 1 Subject | Japanese | Case study | intellectual disability, spastic quadriplegia, agenesis of the corpus callosum and septum pellucidum, irregular ventricular wall, hypoplastic white matter, cerebellar hypoplasia, and fusion of the thalami | cleft palate, micrognathia, abdominal distension, bilateral adducted thumbs, and flexion contractures of the fingers | - | TES | Fluorescent dideoxy terminator method | X-linked, *De novo* | Xq28 (2bp deletion in exon 18 of L1CAM) |
| Okamoto et al., 2004 [81] | Hydrocephalus and Hirschsprung's disease with a mutation of L1CAM | Osaka Medical Center and Research Institute for Maternal and Child Health, 840 Murodo-cho, Izumi, Osaka, Japan | 3 Subjects | Canadian, Spanish | Case series | cerebellar hypoplasia, corpus callosal dysgenesis, thalami fusion, decreased white matter, aqueductal stenosis, intellectual disability, spastic paraparesis | Hirschsprung's disease, bilateral adducted thumbs, flexion contracture of fingers, aphasia | Obstructive | TGS | Direct sequencing | X-linked recessive | Xq28 (Intron 15 mutation and p.Q992X in exon 22 of L1CAM) |
| Panayi et al., 2005 [82] | Prenatal diagnosis in a family with X-linked hydrocephalus | National Taiwan University Hospital, Taipei, Republic of China | 1 Subject | - | Case study | Aqueductal stenosis, underdevelopment of brain tissue, spastic quadriplegia, seizures, and psychomotor retardation | aphasia | Obstructive | TES | Cycle sequencing, SSCP, HA | X-linked, *De novo* | Xq28 (deletion of exon 2 and 6 in L1CAM) |
| Parisi et al., 2002 [83] | Hydrocephalus and intestinal aganglionosis: is L1CAM a modifier gene in Hirschsprung disease? | University of Washington and Children's Hospital and Regional Medical Center, Seattle, Washington 98105, USA | 1 Subject, 1 Control | - | Case study | macrocephaly, aqueductal stenosis, corpus callosum agenesis | bilateral adducted thumbs and index fingers, bilateral inguinal hernias, Hirschsprung's disease, developmental delay, micropenis, small descended right testis, cryptorchid left testis, upgoing toes, limb spasticity, strabismus, amblyopia | Obstructive | TGS | REF, SSCP | X-linked | Xq28 (c.G2254A, p. V752M in exon 18 of L1CAM) |
| Pomili et al., 2000 [84] | MASA syndrome: ultrasonographic evidence in a male fetus | University Hospital, Perugia, Italy | 1 Subject | Italian | Case study | intellectual disability, spasticity of the lower limbs, callosal hypoplasia | colorblindness, bilaterally adducted thumbs | - | TGS | DGGE; direct sequencing | X-linked | Xq28 (G>A base substitution 12 bp upstream from the intron/exon boundary of exon 27 in L1CAM gene) |
| Rehnberg et al., 2010 [85] | Novel L1CAMSplice Site Mutation in a Young Male with L1 Syndrome | Linköping University, University Hospital, Linköping, Sweden | 1 Subject, 3 Family Members | Swedish | Case study | global hypotonia, intellectual disability, spastic paraplegia | bilateral adducted thumbs | - | TGS | Dideoxynucleotide sequencing | X-linked, *De novo* | Xq28 (c.G3458-1C in L1CAM) |
| Rodríguez Criado et al., 2003 [86] | X-linked hydrocephalus: another two families with an L1 mutation | Unidad de Dismorfología, H.I.U.V. Rocío, Sevilla, Spain | 3 Subjects | - | Case series | intellectual disability | aphasia, shuffling gait, and adducted thumbs | Obstructive | TGS | DGGE; REF; direct sequencing | X-linked, *De novo* | Xq28 (c.C196T, p.Q66X in exon 3 of L1CAM; 1267+1G>A in intron 10 of L1CAM) |
| Ruiz et al., 1995 [87] | Mutations in L1-CAM in two families with X linked complicated spastic paraplegia, MASA syndrome, and HSAS | University of Leuven, Belgium | 3 Subjects | - | Case-Contr ol | spastic paresis, intellectual disability, aqueductal stenosis | adducted thumbs | Obstructive | TGS | Solid-phase approach w/ FITC primer; Dot Blot Assay | X-linked | Xq28 (15 bp deletion was found at coding position 97 of the cDNA; 12bp deletion at bp3551; c.T875C; insertion of a cytosine at nucleotide position 3806 within the 3' untranslated region; exons 4,5,6 (c.T556G, p. I179S); exons 8,9,10 (c.G1128A, p.G370R)) |
| Saugier-Weber et al., 1998 [88] | Identification of novel L1CAM mutations using fluorescence-assisted mismatch analysis | Laboratoire de Génétique Moléculaire, CHU de Rouen, France | 13 Subjects, 100 Controls | French | Case series | intellectual disability, spastic paraplegia | aphasia, shuffling gait, adducted thumbs, Hirschsprung's disease | Obstructive | TGS; genotyping | FAMA; ACRS | X-linked, *De novo* | Xq28 (L1CAM nucleotide changes: c.365delC, p.FS122 in exon 4; c.400+5G>A, p.FS108 in intron 4; c.T656C, p. I219T in exon 6; c.T1003C, p. W335R in exon 9; c.C1156T, p. R386C in exon 10; c.C1417T, p. R473C in exon 12; c.C2572T, p. Q858SX in exon 20; c.2872+1G>A in intron 21; c.C3671T, p. S1224L in exon 28; c.3323-30G>A in intron 24) |
| Senat et al., 2001 [89] | Prenatal diagnosis of hydrocephalus-stenosis of the aqueduct of Sylvius by ultrasound in the first trimester of pregnancy. Report of two cases | CHI Poissy, France | 2 Subjects | Caucasian | Case study | corpus callosum agenesis, hypoplasia of pyramidal tract, spasticity, intellectual disability | adducted thumbs | Obstructive | TGS | FAMA | *De novo* | Xq28 (p.Y589H in exon 14 of L1CAM) |
| Serikawa et al., 2014 [90] | Prenatal molecular diagnosis of X-linked hydrocephalus via a silent C924T mutation in the L1CAM gene | Niigata University Medical and Dental Hospital, Niigata, Japan | 4 Subjects, 2 Parents | Japanese | Case study | cortex thinning, cerebral palsy, intellectual disability, corpus callosum agenesis, aqueductal stenosis | bilateral adducted thumbs | Obstructive | TGS | Sanger Sequencing | X-linked | Xq28 (c.C924T, p.G308G silent mutation in exon 8 of L1CAM) |
| Silan et al., 2005 [91] | A novel L1CAM mutation with L1 spectrum disorders | Abant Izzet Baysal University, Duzce School of Medicine, Duzce, Turkey | 14 Subjects | Turkish | Case series | Corpus callosum agenesis, intellectual disability, spastic quadriplegia | bilateral adducted thumbs | - | - | - | X-linked | Xq28 (c.C1375T, Q459X in exon 11 of L1CAM) |
| Stowe et al., 2018 [92] | Clinical Reasoning: Ventriculomegaly detected on 20-week anatomic fetal ultrasound | Baylor College of Medicine, Texas Children's Hospital, Houston | 1 Subject | - | Case study | aqueductal stenosis, diencephalic fusion, and brainstem dysplasia | fisted thumbs | Obstructive | WES | Trio-based WES | X-linked, *De novo* | Xq28 (c.1703+5G>A in L1CAM) |
| Sullivan et al., 2020 [93] | Exome Sequencing as a Potential Diagnostic Adjunct in Sporadic Congenital Hydrocephalus | Yale School of Medicine, New Haven, Connecticut | 475 Subjects | European, African American, south Asian | Case series | hypotonia, cerebral palsy, epilepsy, white-matter hypoplasia, agenesis of the corpus callosum, macrocephaly | bilateral adducted thumbs, skeletal abnormalities | Obstructive | WES | Sanger sequencing | X-linked, *De novo* | Xq28 (L1CAM mutations: p.W460C; p.W635R; c.1828 + 1G>A (localizing to intron 15); c.1546 + 1G>T (located in intron 13); p.E304X; p.V788F; c.806 + 1G>C (positioned in intron 8)) |
| Sztriha et al., 2000 [94] | Novel missense mutation in the L1 gene in a child with corpus callosum agenesis, retardation, adducted thumbs, spastic paraparesis, and hydrocephalus | Faculty of Medicine and Health Sciences, United Arab Emirates University | 1 Subject, 1 Parent, 1 Control | Arabic | Case study | corpus callosum agenesis, intellectual disability, spastic paraparesis | adducted thumbs | Communicating | TES | DGGE analysis | X-linked | Xq28 (c.G604T in exon 6 of L1CAM) |
| Sztriha et al., 2002 [95] | X-linked hydrocephalus: a novel missense mutation in the L1CAM gene | Faculty of Medicine and Health Sciences, United Arab Emirates University, Al Ain | 1 Subject | Pakistani | Case study | spastic diplegia, intellectual disability, multiple small gyri, markedly reduced white matter volume, agenesis of the corpus callosum, and lack of cleavage of the thalami | adducted thumbs | Obstructive | - | - | X-linked | Xq28 (c.G1243C, p.A415P in exon 10 of L1CAM) |
| Takahashi et al., 1997 [96] | L1CAM mutation in a Japanese family with X-linked hydrocephalus: a study for genetic counseling | Asahikawa Medical College, Nishikagura, Japan | 1 Subject, 2 Parents, 2 Sisters | Japanese | Case study | intellectual disability, spastic quadriplegia | bilateral adducted thumbs | Obstructive | TES | - | X-linked | Xq28 (1 bp deletion in exon 22 of L1CAM resulting in a premature stop codon) |
| Takechi et al., 1996 [97] | A deletion of five nucleotides in the L1CAM gene in a Japanese family with X-linked hydrocephalus | National Institute of Neuroscience, Tokyo, Japan | 2 Subjects, 1 Sister | Japanese | Case study | aqueduct of Sylvius, mental retardation, and spastic paraparesis | bilateral clasped thumbs | - | TES | Dideoxy plasmid-based sequencing | X-linked | Xq28 (5bp deletion in exon 8 of L1CAM) |
| Takenouchi et al., 2011 [98] | Hydrocephalus with Hirschsprung disease: severe end of X-linked hydrocephalus spectrum | Keio University School of Medicine, Tokyo, Japan | 1 Subject | Japanese | Case study | aqueductal stenosis, hypoplasia of the corpus callosum | Hirschsprung disease, frontal bossing, adducted thumbs | Obstructive | TGS | Mutation analysis, unspecified | X-linked, *De novo* | Xq28 (c.C61T, p.Q21X in exon 1 of L1CAM) |
| Tegay et al., 2007 [99] | Contiguous gene deletion involving L1CAM and AVPR2 causes X-linked hydrocephalus with nephrogenic diabetes insipidus | Stony Brook University Hospital, Stony Brook, New York, USA | 1 Subject, Mother, Grandmother, 1 Control | Northern European | Case study | hypotonia | bilateral adducted thumbs, Hirschsprung disease | Obstructive | WGS | GeneDX, microdeletion | X-linked | Xq28 (32.7 kb deletion and 90 bp insertion at the L1CAM-AVPR2 junction sequence (from L1CAM intron1 to AVPR2 exon2)) |
| Van Camp et al., 1993 [100] | A duplication in the L1CAM gene associated with X-linked hydrocephalus | University of Antwerp-UIA, Belgium | 25 Subjects, Controls used | The Netherlands, United Kingdom, USA, Israel, Germany, Hungary, Belgium | Case series | stenosis of the aqueduct of Sylvius, intellectual disability, spastic paraparesis of the lower extremities, aplasia or hypoplasia of the corpus callosum | bilateral adducted thumbs | - | Southern Blotting | - | X-linked recessive | Xq28 (1.3 kb duplication in L1CAM) |
| Verhagen et al., 1998 [101] | Familial congenital hydrocephalus and aqueduct stenosis with probably autosomal dominant inheritance and variable expression | Canisius Wilhelmina Hospital, Nijmegen, Netherlands | 12 Subjects | - | Case series | septum pellucidum cavitation, aqueductal stenosis | - | Obstructive | TES | - | AD | No mutations in L1CAM |
| Vits et al., 1994 [102] | MASA syndrome is due to mutations in the neural cell adhesion gene L1CAM | University of Antwerp, Belgium | 8 Subjects, 50 Controls | United States, the Netherlands, Mexico, UK, Germany | Case series | intellectual disability | adducted thumbs, shuffling gait, aphasia | - | TGS | SSCP | X-linked | Xq28 (p.D598N in exon 14 and p.H210Q in exon 6 of L1CAM) |
| Vos et al., 2010 [103] | Genotype-phenotype correlations in L1 syndrome: a guide for genetic counselling and mutation analysis | University Medical Centre Groningen, Hanzeplein 1, 9713 GZ Groningen, The Netherlands | 367 Subjects, 3 Controls | Various | Case-Control | aqueductal stenosis, intellectual disability, callosal agenesis | adducted thumbs, shuffling gait, aphasia | Obstructive | TES | DGGE; direct sequencing; MLPA | X-linked recessive | Xq28 (L1CAM mutations: 23 missense mutations; 3 in-frame deletions/duplications; 18 splice site mutations; 14 nonsense mutations; 8 frame-shift mutations; 1 duplication of exons 2-10; 1 deletion of the entire gene; c.C645T within exon 6 of L1CAM) |
| Wilson et al., 2009 [104] | Prenatal identification of a novel R937P L1CAM missense mutation | University of Oklahoma Health Sciences Center, Oklahoma City, Oklahoma, USA | 2 Subjects | Caucasian | Case study | aqueductal stenosis, agenesis or hypoplasia of the corpus callosum and corticospinal tracts, intellectual disability, spastic paraplegia | adducted thumbs, short femurs, right clubbed foot | Obstructive | TGS | bidirectional DNA sequencing | X-linked | Xq28 (c.G2809C, p.R937P in exon 21 of L1CAM) |
| Xie et al., 2018 [105] | Two novel pathogenic variants of L1CAM gene in two fetuses with isolated X-linked hydrocephaly: A case report | Guangxi Maternal and Child Health Hospital, Nanning, Guangxi, P.R. China | 2 Subjects, 4 Parents, 100 Controls | Chinese | Case-Contr ol | isolated hydrocephalus | - | - | TES | Sanger sequencing | X-linked | Xq28 (c.C998T, p.P333L and c.G2362T, p.V788F in L1CAM) |
| Yamasaki et al., 2011 [106] | Prenatal molecular diagnosis of a severe type of L1 syndrome (X-linked hydrocephalus) | Osaka National Hospital, National Hospital Organization, Osaka City, Japan | 14 Subjects | Japanese | Case series | intellectual disability, spastic paraplegia | adducted thumbs, shuffling gait, aphasia | - | TGS | Direct sequencing | X-linked | Xq28 (L1CAM mutations: c.G1829-1C 1 bp downstream from the 5' of intron 14; ACC (817–819) nucleotide deletion in exon 8, deletion of T at amino acid position 273; c.C1146A, p.Y382X in exon 10) |

### Dandy Walker Malformation

Dandy Walker malformation is a cerebellar structural anomaly that can impede CSF flow but can also be related to primary brain developmental alterations and contribute to HC development. Missense mutations are found in forkhead box C1 (*FOXC1*), fukutin (*FKTN*), laminin subunit gamma 1 (*LAMC1*), sphingosine-1-phosphate phosphatase 2 (*SGPP2*), and exocyst complex component 3 like 2 (*EXOC3L2*). Nonsense mutations are found in FKTN, nidogen 1 (*NID1*), and potassium channel tetramerization domain containing 3 (*KCTD3*). SIL1 nucleotide exchange factor (*SIL1*) displayed a nonstop mutation and carnitine palmitoyltransferase 2 (*CPT2*) displayed a deletion-insertion variant. Additional mutations included Zic family member 2 (*ZIC2*) and Zic family member 5 (*ZIC5).* Deletions were found in lysine methyltransferase 2D (*KMT2D*), chromosome 2 (2q36.1), chromosome 3 (3q25.1), chromosome 6 (6p24.1, 6p25.3), chromosome 7 (7p21.3), chromosome 8 (8q21), chromosome 12 (12q24), chromosome 13 (13q32), and chromosome 16 (16q21). The deletion of 8p21 resulted in the downregulation of fibroblast growth factor 17 (*FGF17*). Duplications were found in chromosome 6 (6p25.3), chromosome 7 (7p21.3), and chromosome 12 (12q24). In addition, EXOC3L2 regulates vesicular trafficking at synapses and cell polarity; a mutation within this gene locus can impact normal brain development [44]. *KCTD3* is also highly expressed in the brain and kidneys and regulates ion channels such as hyperpolarization activated cyclic nucleotide-gated channel 3 (HCN3) [45]. SIL1 is a glycoprotein that regulates protein trafficking into the ER and ATPase activity, suggesting a mutated implication in protein folding through development [46; 47]. A patient with a mutation in *CPT2*, an enzyme responsible for breaking down long chain fatty acids, suggests a role of metabolic enzymes in the genetic susceptibility of HC secondary to Dandy Walker malformation [48]. Thus, Dandy Walker malformation related HC may be caused by a wide variety of genes involved in many biological processes. These data are summarized in **Table 4**.

**Table 4:**
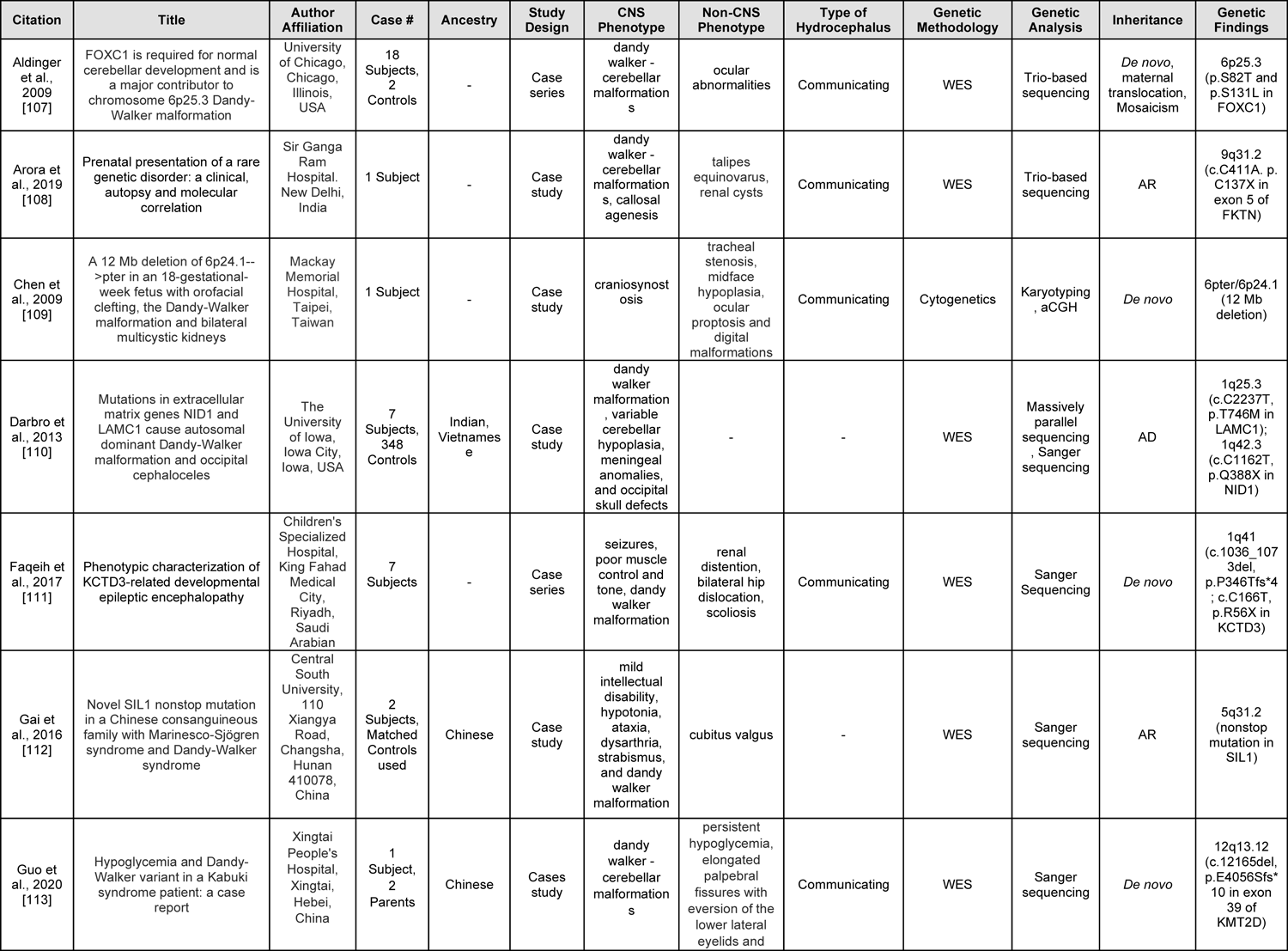
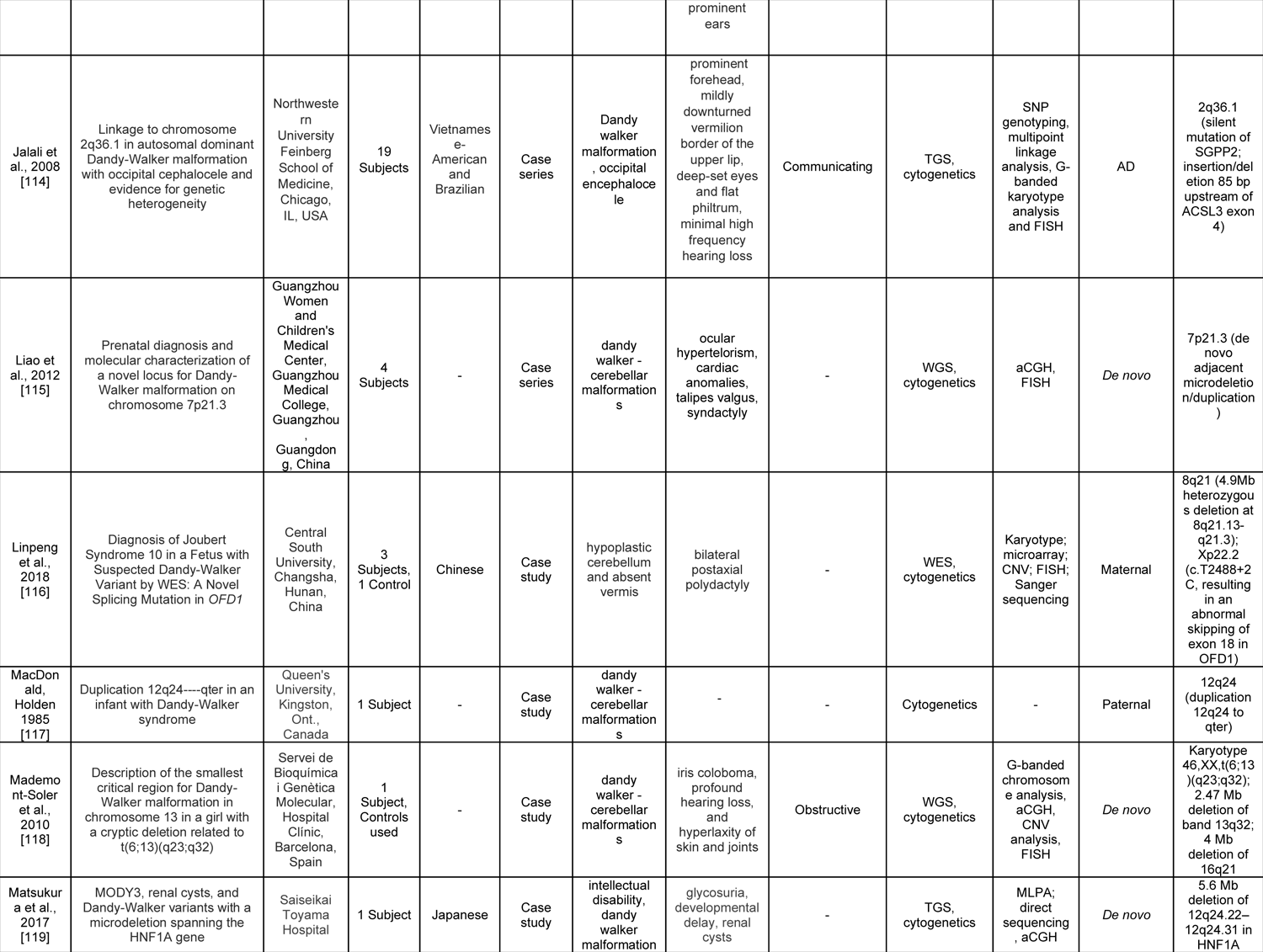
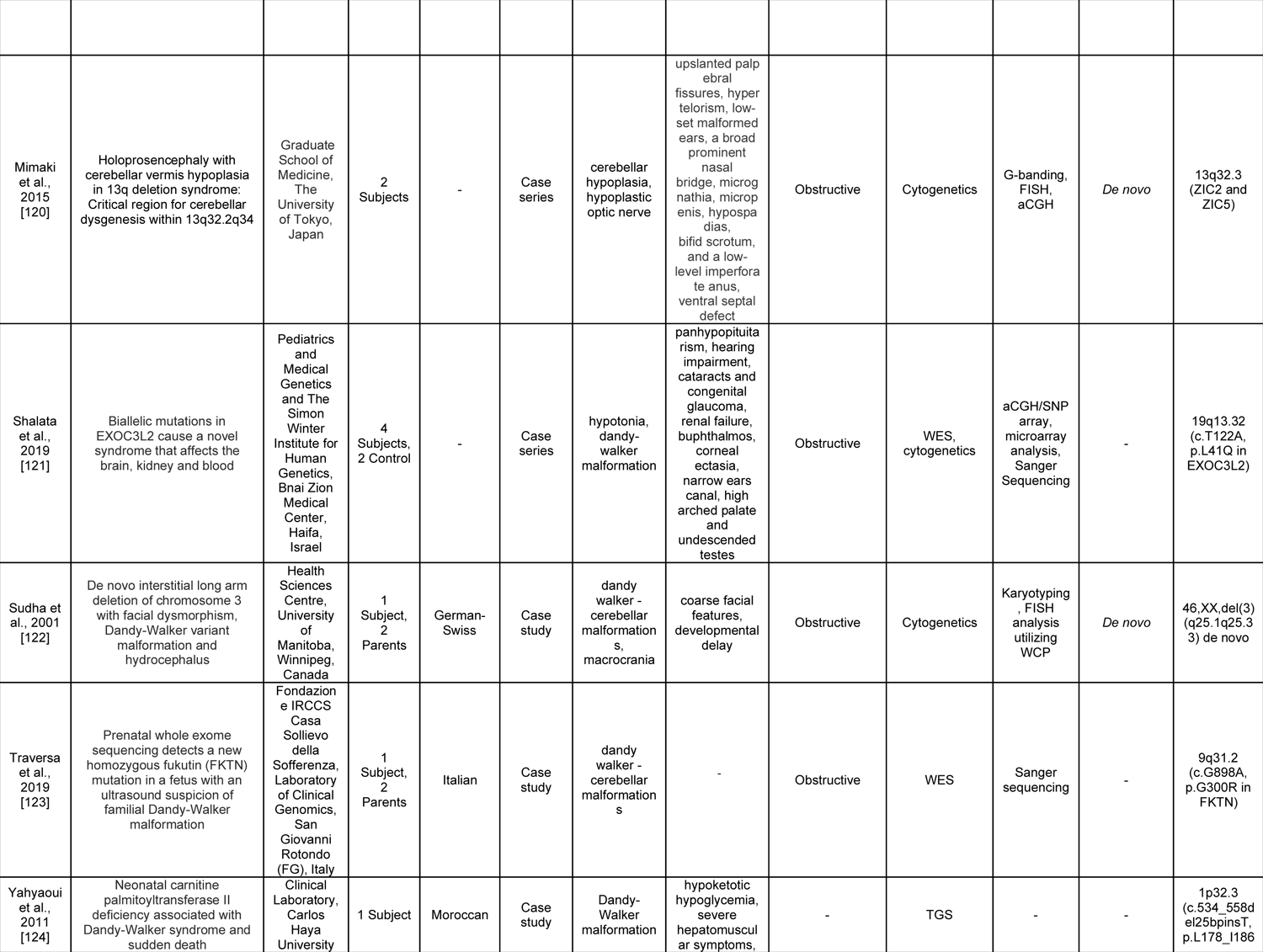
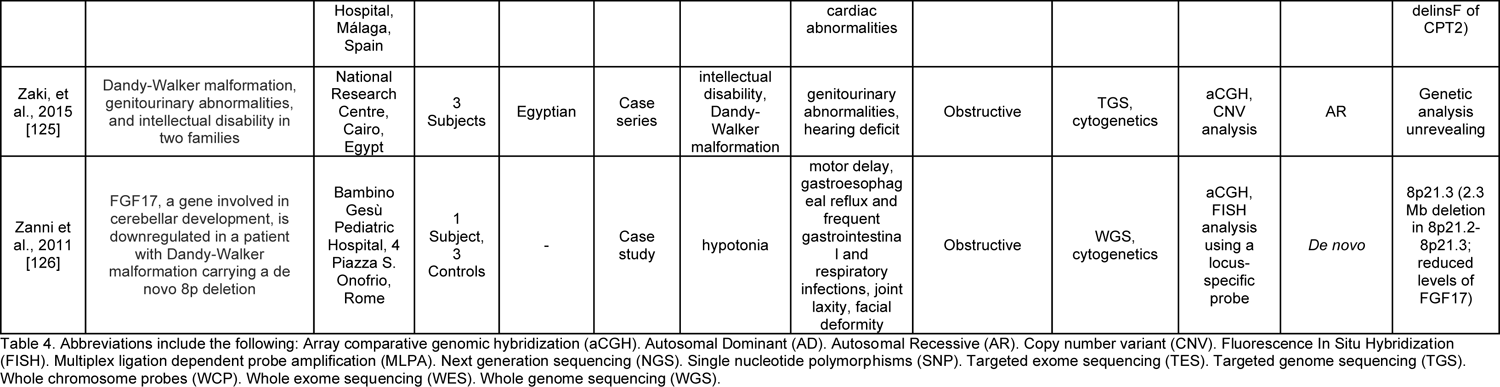
Dandy Walker Malformation.

Abbreviations include the following: Array comparative genomic hybridization (aCGH).
| Citation | Title | Author Affiliation | Case # | Ancestry | Study Design | CNS Phenotype | Non-CNS Phenotype | Type of Hydrocephalus | Genetic Methodology | Genetic Analysis | Inheritance | Genetic Findings |
| --- | --- | --- | --- | --- | --- | --- | --- | --- | --- | --- | --- | --- |
| Aldinger et al., 2009 [107] | FOXC1 is required for normal cerebellar development and is a major contributor to chromosome 6p25.3 Dandy-Walker malformation | University of Chicago, Chicago, Illinois, USA | 18 Subjects, 2 Controls | - | Case series | dandy walker - cerebellar malformation s | ocular abnormalities | Communicating | WES | Trio-based sequencing | *De novo*, maternal translocation, Mosaicism | 6p25.3 (p.S82T and p.S131L in FOXC1) |
| Arora et al., 2019 [108] | Prenatal presentation of a rare genetic disorder: a clinical, autopsy and molecular correlation | Sir Ganga Ram Hospital, New Delhi, India | 1 Subject | - | Case study | dandy walker - cerebellar malformation s, callosal agenesis | talipes equinovarus, renal cysts | Communicating | WES | Trio-based sequencing | AR | 9q31.2 (c.C411A, p. C137X in exon 5 of FKTN) |
| Chen et al., 2009 [109] | A 12 Mb deletion of 6p24.1-->pter in an 18-gestational-week fetus with orofacial clefting, the Dandy-Walker malformation and bilateral multicystic kidneys | Mackay Memorial Hospital, Taipei, Taiwan | 1 Subject | - | Case study | craniosynostosis | tracheal stenosis, midface hypoplasia, ocular proptosis and digital malformations | Communicating | Cytogenetics | Karyotyping , aCGH | *De novo* | 6pter/6p24.1 (12 Mb deletion) |
| Darbro et al., 2013 [110] | Mutations in extracellular matrix genes NID1 and LAMC1 cause autosomal dominant Dandy-Walker malformation and occipital cephaloceles | The University of Iowa, Iowa City, Iowa, USA | 7 Subjects, 348 Controls | Indian, Vietnamese | Case study | dandy walker malformation , variable cerebellar hypoplasia, meningeal anomalies, and occipital skull defects | - | - | WES | Massively parallel sequencing , Sanger sequencing | AD | 1q25.3 (c.C2237T, p.T746M in LAMC1); 1q42.3 (c.C1162T, p.Q388X in NID1) |
| Faqeih et al., 2017 [111] | Phenotypic characterization of KCTD3-related developmental epileptic encephalopathy | Children's Specialized Hospital, King Fahad Medical City, Riyadh, Saudi Arabian | 7 Subjects | - | Case series | seizures, poor muscle control and tone, dandy walker malformation | renal distention, bilateral hip dislocation, scoliosis | Communicating | WES | Sanger Sequencing | *De novo* | 1q41 (c.1036_1073del, p.P346Tfs*4 ; c.C166T, p.R56X in KCTD3) |
| Gai et al., 2016 [112] | Novel SIL1 nonstop mutation in a Chinese consanguineous family with Marinesco-Sjögren syndrome and Dandy-Walker syndrome | Central South University, 110 Xiangya Road, Changsha, Hunan 410078, China | 2 Subjects, Matched Controls used | Chinese | Case study | mild intellectual disability, hypotonia, ataxia, dysarthria, strabismus, and dandy walker malformation | cubitus valgus | - | WES | Sanger sequencing | AR | 5q31.2 (nonstop mutation in SIL1) |
| Guo et al., 2020 [113] | Hypoglycemia and Dandy-Walker variant in a Kabuki syndrome patient: a case report | Xingtai People's Hospital, Xingtai, Hebei, China | 1 Subject, 2 Parents | Chinese | Cases study | dandy walker - cerebellar malformation s | persistent hypoglycemia, elongated palpebral fissures with eversion of the lower lateral eyelids and prominent ears | Communicating | WES | Sanger sequencing | *De novo* | 12q13.12 (c.12165del, p.E4056Sfs*10 in exon 39 of KMT2D) |
| Jalali et al., 2008 [114] | Linkage to chromosome 2q36.1 in autosomal dominant Dandy-Walker malformation with occipital cephalocele and evidence for genetic heterogeneity | Northwestern University Feinberg School of Medicine, Chicago, IL, USA | 19 Subjects | Vietnamese-American and Brazilian | Case series | Dandy walker malformation, occipital encephalocele | prominent forehead, mildly downturned vermilion border of the upper lip, deep-set eyes and flat philtrum, minimal high frequency hearing loss | Communicating | TGS, cytogenetics | SNP genotyping, multipoint linkage analysis, G-banded karyotype analysis and FISH | AD | 2q36.1 (silent mutation of SGPP2; insertion/deletion 85 bp upstream of ACSL3 exon 4) |
| Liao et al., 2012 [115] | Prenatal diagnosis and molecular characterization of a novel locus for Dandy-Walker malformation on chromosome 7p21.3 | Guangzhou Women and Children's Medical Center, Guangzhou Medical College, Guangzhou, Guangdong, China | 4 Subjects | - | Case series | dandy walker - cerebellar malformations | ocular hypertelorism, cardiac anomalies, talipes valgus, syndactyly | - | WGS, cytogenetics | aCGH, FISH | *De novo* | 7p21.3 (de novo adjacent microdeletion/duplication) |
| Linpeng et al., 2018 [116] | Diagnosis of Joubert Syndrome 10 in a Fetus with Suspected Dandy-Walker Variant by WES: A Novel Splicing Mutation in *OFD1* | Central South University, Changsha, Hunan, China | 3 Subjects, 1 Control | Chinese | Case study | hypoplastic cerebellum and absent vermis | bilateral postaxial polydactyly | - | WES, cytogenetics | Karyotype; microarray; CNV; FISH; Sanger sequencing | Maternal | 8q21 (4.9Mb heterozygous deletion at 8q21.13-q21.3); Xp22.2 (c.T2488+2 C, resulting in an abnormal skipping of exon 18 in OFD1) |
| MacDonald, Holden 1985 [117] | Duplication 12q24----qter in an infant with Dandy-Walker syndrome | Queen's University, Kingston, Ont., Canada | 1 Subject | - | Case study | dandy walker - cerebellar malformations | - | - | Cytogenetics | - | Paternal | 12q24 (duplication 12q24 to qter) |
| Mademont-Soler et al., 2010 [118] | Description of the smallest critical region for Dandy-Walker malformation in chromosome 13 in a girl with a cryptic deletion related to t(6;13)(q23;q32) | Servei de Bioquímica i Genètica Molecular, Hospital Clínic, Barcelona, Spain | 1 Subject, Controls used | - | Case study | dandy walker - cerebellar malformations | iris coloboma, profound hearing loss, and hyperlaxity of skin and joints | Obstructive | WGS, cytogenetics | G-banded chromosome analysis, aCGH, CNV analysis, FISH | *De novo* | Karyotype 46,XX,t(6;13)(q23;q32); 2.47 Mb deletion of band 13q32; 4 Mb deletion of 16q21 |
| Matsukura et al., 2017 [119] | MODY3, renal cysts, and Dandy-Walker variants with a microdeletion spanning the HNF1A gene | Saiseikai Toyama Hospital | 1 Subject | Japanese | Case study | intellectual disability, dandy walker malformation | glycosuria, developmental delay, renal cysts | - | TGS, cytogenetics | MLPA; direct sequencing, aCGH | *De novo* | 5.6 Mb deletion of 12q24.22–12q24.31 in HNF1A |
| Mimaki et al., 2015 [120] | Holoprosencephaly with cerebellar vermis hypoplasia in 13q deletion syndrome: Critical region for cerebellar dysgenesis within 13q32.2q34 | Graduate School of Medicine, The University of Tokyo, Japan | 2 Subjects | - | Case series | cerebellar hypoplasia, hypoplastic optic nerve | upslanted palpebral fissures, hypertelorism, low-set malformed ears, a broad prominent nasal bridge, micrognathia, micropenis, hypospadias, bifid scrotum, and a low-level imperforate anus, ventral septal defect | Obstructive | Cytogenetics | G-banding, FISH, aCGH | *De novo* | 13q32.3 (ZIC2 and ZIC5) |
| Shalata et al., 2019 [121] | Biallelic mutations in EXOC3L2 cause a novel syndrome that affects the brain, kidney and blood | Pediatrics and Medical Genetics and The Simon Winter Institute for Human Genetics, Bnai Zion Medical Center, Haifa, Israel | 4 Subjects, 2 Control | - | Case series | hypotonia, dandy-walker malformation | panhypopituitarism, hearing impairment, cataracts and congenital glaucoma, renal failure, buphthalmos, corneal ectasia, narrow ears canal, high arched palate and undescended testes | Obstructive | WES, cytogenetics | aCGH/SNP array, microarray analysis, Sanger Sequencing | - | 19q13.32 (c.T122A, p.L41Q in EXOC3L2) |
| Sudha et al., 2001 [122] | De novo interstitial long arm deletion of chromosome 3 with facial dysmorphism, Dandy-Walker variant malformation and hydrocephalus | Health Sciences Centre, University of Manitoba, Winnipeg, Canada | 1 Subject, 2 Parents | German-Swiss | Case study | dandy walker - cerebellar malformations, macrocrania | coarse facial features, developmental delay | Obstructive | Cytogenetics | Karyotyping, FISH analysis utilizing WCP | *De novo* | 46,XX,del(3)(q25.1q25.33) de novo |
| Traversa et al., 2019 [123] | Prenatal whole exome sequencing detects a new homozygous fukutin (FKTN) mutation in a fetus with an ultrasound suspicion of familial Dandy-Walker malformation | Fondazione IRCCS Casa Sollievo della Sofferenza, Laboratory of Clinical Genomics, San Giovanni Rotondo (FG), Italy | 1 Subject, 2 Parents | Italian | Case study | dandy walker - cerebellar malformations | - | Obstructive | WES | Sanger sequencing | - | 9q31.2 (c.G898A, p.G300R in FKTN) |
| Yahyaoui et al., 2011 [124] | Neonatal carnitine palmitoyltransferase II deficiency associated with Dandy-Walker syndrome and sudden death | Clinical Laboratory, Carlos Haya University | 1 Subject | Moroccan | Case study | Dandy-Walker malformation | hypoketotic hypoglycemia, severe hepatomuscular symptoms, | - | TGS | - | - | 1p32.3 (c.534_558del25bpinsT, p.L178_I186 |
|  |  | Hospital, Málaga, Spain |  |  |  |  | cardiac abnormalities |  |  |  |  | delinsF of CPT2) |
| Zaki, et al., 2015 [125] | Dandy-Walker malformation, genitourinary abnormalities, and intellectual disability in two families | National Research Centre, Cairo, Egypt | 3 Subjects | Egyptian | Case series | intellectual disability, Dandy-Walker malformation | genitourinary abnormalities, hearing deficit | Obstructive | TGS, cytogenetics | aCGH, CNV analysis | AR | Genetic analysis unrevealing |
| Zanni et al., 2011 [126] | FGF17, a gene involved in cerebellar development, is downregulated in a patient with Dandy-Walker malformation carrying a de novo 8p deletion | Bambino Gesù Pediatric Hospital, 4 Piazza S. Onofrio, Rome | 1 Subject, 3 Controls | - | Case study | hypotonia | motor delay, gastroesophageal reflux and frequent gastrointestinal and respiratory infections, joint laxity, facial deformity | Obstructive | WGS, cytogenetics | aCGH, FISH analysis using a locus-specific probe | *De novo* | 8p21.3 (2.3 Mb deletion in 8p21.2-8p21.3; reduced levels of FGF17) |

#### Ciliopathy

Genes involved in cilia function that are associated with HC are summarized in **Table 5**. Primary cilia dysfunction has been demonstrated to play a role in HC with numerous Mendelian ‘ciliopathies’ resulting in HC. Missense mutations were observed in Meckel-Gruber syndrome gene (*MKS3*), MKS transition zone complex subunit 1 (*MKS1*), intraflagellar transport 43 (*IFT43*), WD repeat domain 35 (*IFT121*), coiled-coil and C2 domain containing 2A (*CC2D2A*), transmembrane protein 216 (*TMEM216*), PKHD1 ciliary IPT domain containing fibrocystin/polyductin (*PKHD1*), intestinal cell kinase (*ICK*), exon 14 of KIAA0586, exons 4 and 13 of centrosomal protein 83 (*CEP83*), exons 6, 11, 12, 20, 23, 24, 28, 29, 32, and 36 of SET binding factor 2 (*SBF2*), exon 9 of zinc finger E-box binding homeobox 1 (*ZEB1*), and exon 5 of G protein subunit alpha i2 (*GNAI2*). Nonsense mutations were identified in *CC2D2A*, *IFT121*, forkhead box J1 (*FOXJ1*), exon 2 of *KIAA0586*, exon 3 of centrosomal protein 55 (*CEP55*), exons 3, 4, 7, and 13 of *CEP83*, and exon 11 of *SBF2*. Deletions and duplications resulting in frameshift mutations were found in *CC2D2A*, *MKS3*, *MKS1*, dynein axonemal intermediate chain 2 (*DNAI2*), *IFT121*, *FOXJ1*, exon 5 and 17 of *CEP83*, and exon 4 of *ZEB1*. Exon 2 was deleted in WD repeat-containing protein 16 (*WDR16*). Additional mutations were found in WD repeat domain 93 (*WDR93*). Loss of *MKS3* and *MKS1* are associated with ciliary shortening and dysfunction, suggesting a role in primary ciliary development. *TMEM216* also contributes to ciliary development through apical polarization and formation and may result in Joubert, Meckel and related syndromes [49]. IFT43 and IFT121 maintain cilium organization and regulate intraflagellar transport in interaction with the IFT-A complex [50]. In addition, CEP83 also interacts with IFT proteins and guides vesicular docking ciliogenesis [51]. One patient was identified with a mutation in *DNAI2*, a component of the outer dynein arm complex (ODA), which is involved in cilia motility [52]. ZEB1, SBF2, and GNAI2 are involved in other signaling pathways previously identified in association with HC [53].

**Table 5:**
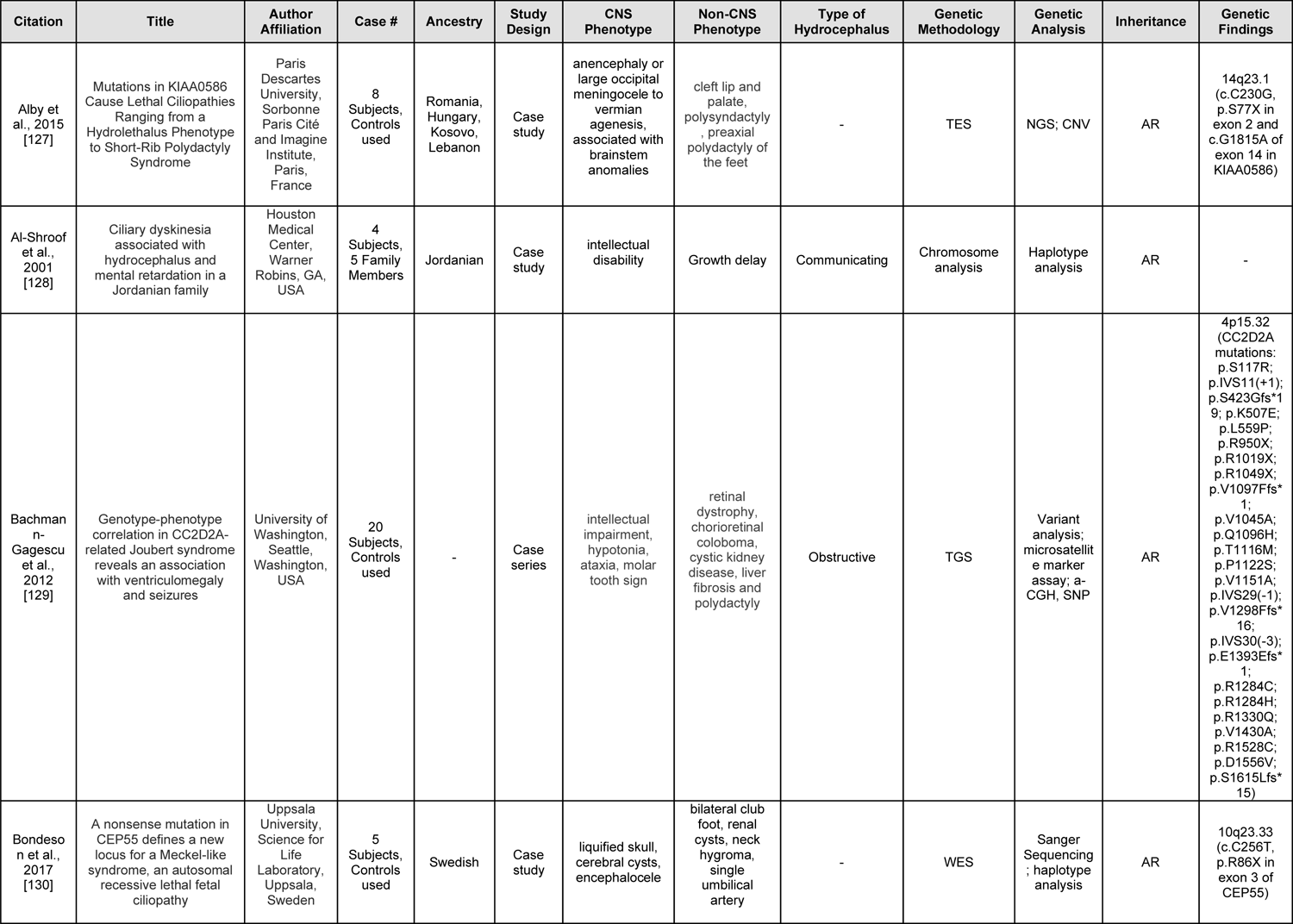
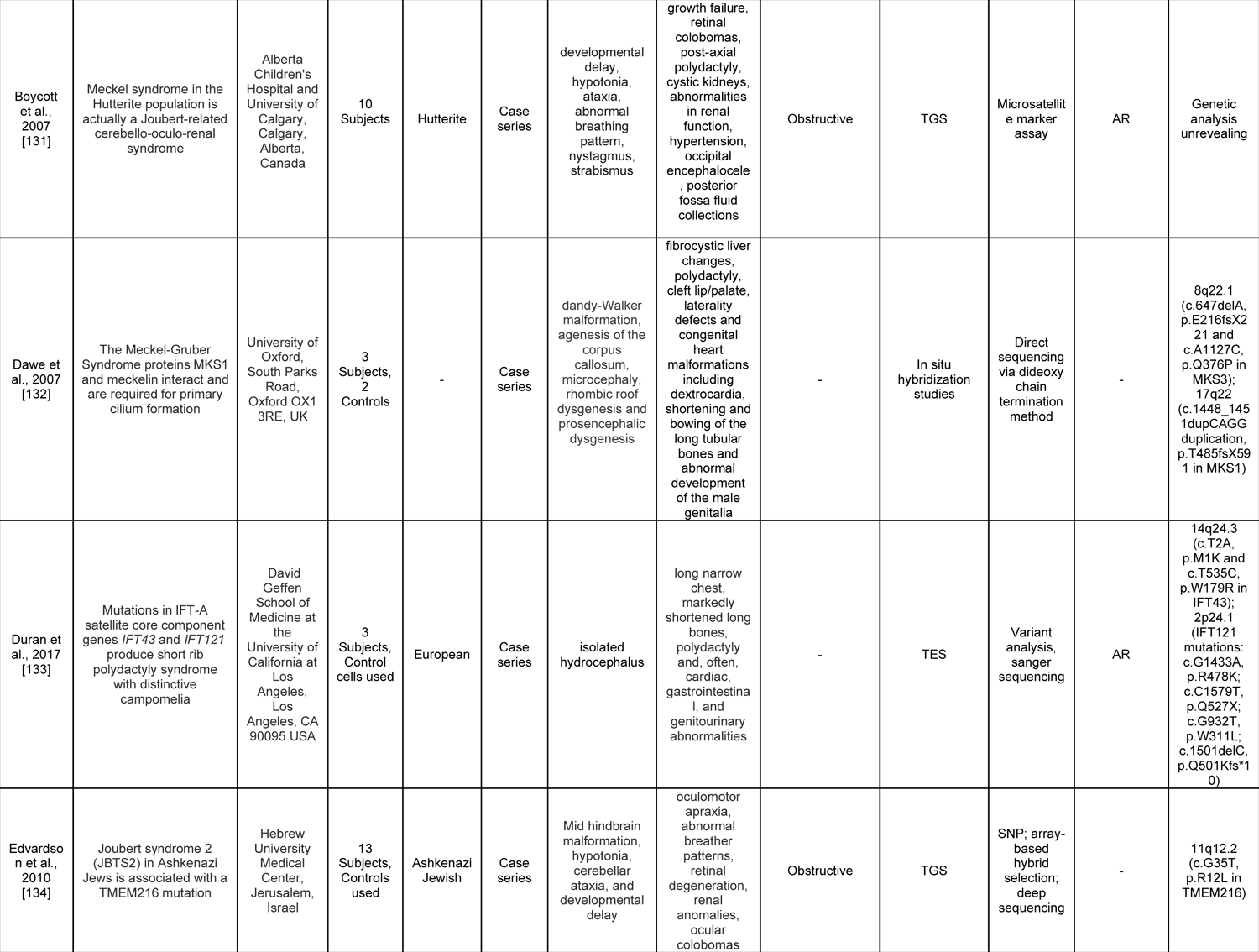
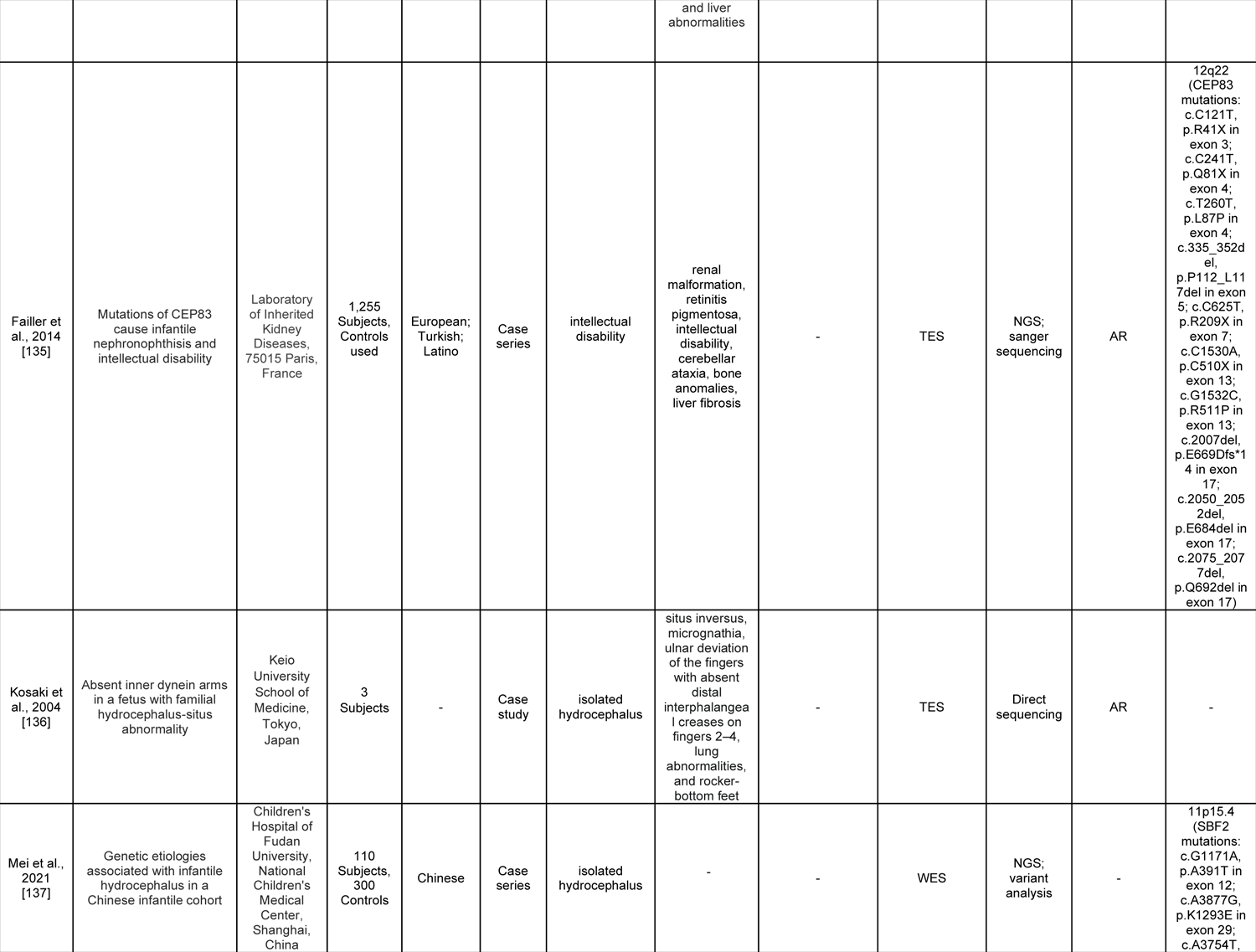
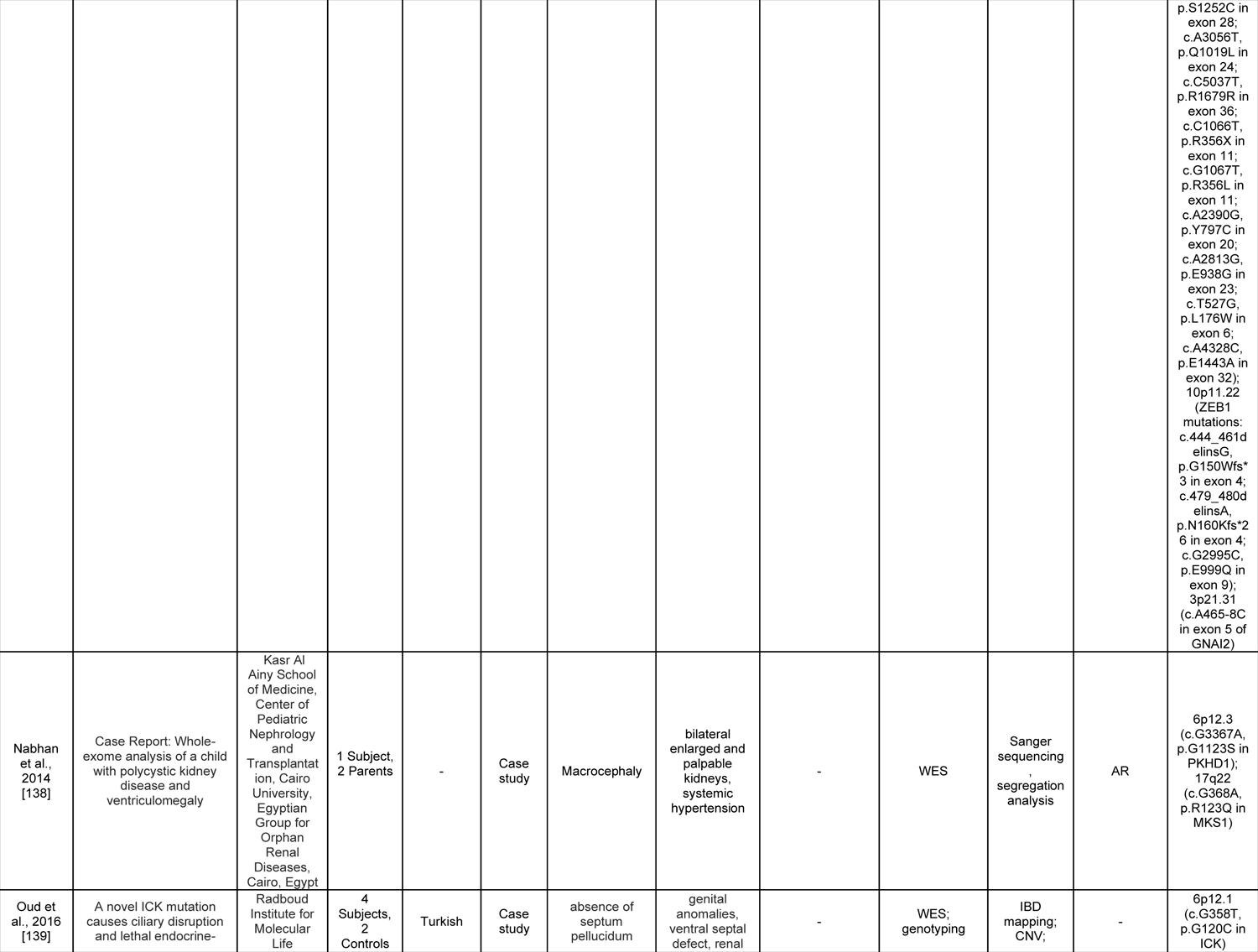
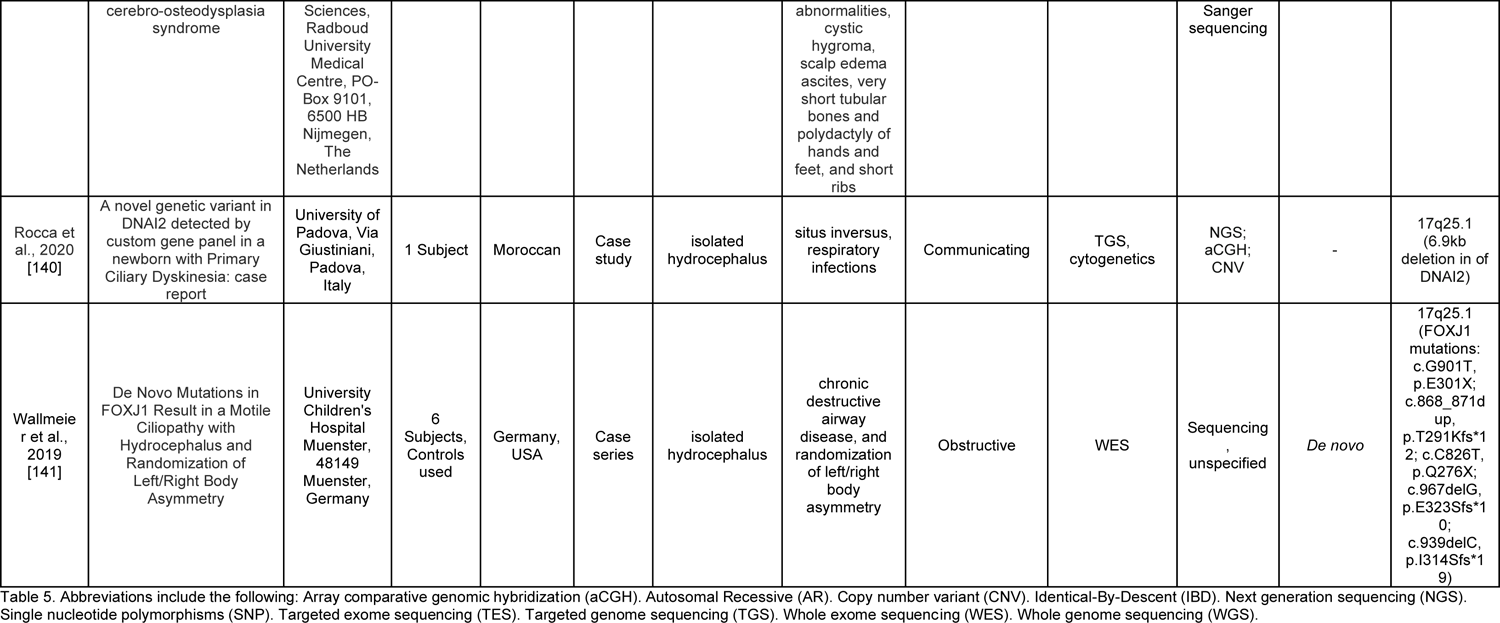
Ciliopathy.

Abbreviations include the following: Array comparative genomic hybridization (aCGH).
| Citation | Title | Author Affiliation | Case # | Ancestry | Study Design | CNS Phenotype | Non-CNS Phenotype | Type of Hydrocephalus | Genetic Methodology | Genetic Analysis | Inheritance | Genetic Findings |
| --- | --- | --- | --- | --- | --- | --- | --- | --- | --- | --- | --- | --- |
| Alby et al., 2015 [127] | Mutations in KIAA0586 Cause Lethal Ciliopathies Ranging from a Hydroletharus Phenotype to Short-Rib Polydactyly Syndrome | Paris Descartes University, Sorbonne Paris Cité and Imagine Institute, Paris, France | 8 Subjects, Controls used | Romania, Hungary, Kosovo, Lebanon | Case study | anencephaly or large occipital meningocele to vermian agenesis, associated with brainstem anomalies | cleft lip and palate, polysyndactyly , preaxial polydactyly of the feet | - | TES | NGS; CNV | AR | 14q23.1 (c.C230G, p.S77X in exon 2 and c.G1815A of exon 14 in KIAA0586) |
| Al-Shroof et al., 2001 [128] | Ciliary dyskinesia associated with hydrocephalus and mental retardation in a Jordanian family | Houston Medical Center, Warner Robins, GA, USA | 4 Subjects, 5 Family Members | Jordanian | Case study | intellectual disability | Growth delay | Communicating | Chromosome analysis | Haplotype analysis | AR | - |
| Bachmann-Gagescu et al., 2012 [129] | Genotype-phenotype correlation in CC2D2A-related Joubert syndrome reveals an association with ventriculomegaly and seizures | University of Washington, Seattle, Washington, USA | 20 Subjects, Controls used | - | Case series | intellectual impairment, hypotonia, ataxia, molar tooth sign | retinal dystrophy, chorioretinal coloboma, cystic kidney disease, liver fibrosis and polydactyly | Obstructive | TGS | Variant analysis; microsatellite marker assay; a-CGH, SNP | AR | 4p15.32 (CC2D2A mutations: p.S117R; p.IVS11(+1); p.S423Gfs*19; p.K507E; p.L559P; p.R950X; p.R1019X; p.R1049X; p.V1097Ffs*1; p.V1045A; p.Q1096H; p.T1116M; p.P1122S; p.V1151A; p.IVS29(-1); p.V1298Ffs*16; p.IVS30(-3); p.E1393Efs*1; p.R1284C; p.R1284H; p.R1330Q; p.V1430A; p.R1528C; p.D1556V; p.S1615Lfs*15) |
| Bondeson et al., 2017 [130] | A nonsense mutation in CEP55 defines a new locus for a Meckel-like syndrome, an autosomal recessive lethal fetal ciliopathy | Uppsala University, Science for Life Laboratory, Uppsala, Sweden | 5 Subjects, Controls used | Swedish | Case study | liquified skull, cerebral cysts, encephalocele | bilateral club foot, renal cysts, neck hygroma, single umbilical artery | - | WES | Sanger Sequencing ; haplotype analysis | AR | 10q23.33 (c.C256T, p.R86X in exon 3 of CEP55) |
| Boycott et al., 2007 [131] | Meckel syndrome in the Hutterite population is actually a Joubert-related cerebello-oculo-renal syndrome | Alberta Children's Hospital and University of Calgary, Calgary, Alberta, Canada | 10 Subjects | Hutterite | Case series | developmental delay, hypotonia, ataxia, abnormal breathing pattern, nystagmus, strabismus | growth failure, retinal colobomas, post-axial polydactyly, cystic kidneys, abnormalities in renal function, hypertension, occipital encephalocele, posterior fossa fluid collections | Obstructive | TGS | Microsatellite marker assay | AR | Genetic analysis unrevealing |
| Dawe et al., 2007 [132] | The Meckel-Gruber Syndrome proteins MKS1 and meckelin interact and are required for primary cilium formation | University of Oxford, South Parks Road, Oxford OX1 3RE, UK | 3 Subjects, 2 Controls | - | Case series | dandy-Walker malformation, agenesis of the corpus callosum, microcephaly, rhombic roof dysgenesis and prosencephalic dysgenesis | fibrocystic liver changes, polydactyly, cleft lip/palate, laterality defects and congenital heart malformations including dextrocardia, shortening and bowing of the long tubular bones and abnormal development of the male genitalia | - | In situ hybridization studies | Direct sequencing via dideoxy chain termination method | - | 8q22.1 (c.647delA, p.E216fsX221 and c.A1127C, p.Q376P in MKS3); 17q22 (c.1448_1451dupCAGG duplication, p.T485fsX591 in MKS1) |
| Duran et al., 2017 [133] | Mutations in IFT-A satellite core component genes *IFT43* and *IFT121* produce short rib polydactyly syndrome with distinctive campomelia | David Geffen School of Medicine at the University of California at Los Angeles, Los Angeles, CA 90095 USA | 3 Subjects, Control cells used | European | Case series | isolated hydrocephalus | long narrow chest, markedly shortened long bones, polydactyly and, often, cardiac, gastrointestinal, and genitourinary abnormalities | - | TES | Variant analysis, sanger sequencing | AR | 14q24.3 (c.T2A, p.M1K and c.T535C, p.W179R in IFT43); 2p24.1 (IFT121 mutations: c.G1433A, p.R478K; c.C1579T, p.Q527X; c.G932T, p.W311L; c.1501delC, p.Q501Kfs*10) |
| Edvardson et al., 2010 [134] | Joubert syndrome 2 (JBTS2) in Ashkenazi Jews is associated with a TMEM216 mutation | Hebrew University Medical Center, Jerusalem, Israel | 13 Subjects, Controls used | Ashkenazi Jewish | Case series | Mid hindbrain malformation, hypotonia, cerebellar ataxia, and developmental delay | oculomotor apraxia, abnormal breather patterns, retinal degeneration, renal anomalies, ocular colobomas and liver abnormalities | Obstructive | TGS | SNP; array-based hybrid selection; deep sequencing | - | 11q12.2 (c.G35T, p.R12L in TMEM216) |
| Failler et al., 2014 [135] | Mutations of CEP83 cause infantile nephronophthisis and intellectual disability | Laboratory of Inherited Kidney Diseases, 75015 Paris, France | 1,255 Subjects, Controls used | European; Turkish; Latino | Case series | intellectual disability | renal malformation, retinitis pigmentosa, intellectual disability, cerebellar ataxia, bone anomalies, liver fibrosis | - | TES | NGS; sanger sequencing | AR | 12q22 (CEP83 mutations: c.C121T, p.R41X in exon 3; c.C241T, p.Q81X in exon 4; c.T260T, p.L87P in exon 4; c.335_352del, p.P112_L117del in exon 5; c.C625T, p.R209X in exon 7; c.C1530A, p.C510X in exon 13; c.G1532C, p.R511P in exon 13; c.2007del, p.E669Dfs*14 in exon 17; c.2050_2052del, p.E684del in exon 17; c.2075_2077del, p.Q692del in exon 17) |
| Kosaki et al., 2004 [136] | Absent inner dynein arms in a fetus with familial hydrocephalus-situs abnormality | Keio University School of Medicine, Tokyo, Japan | 3 Subjects | - | Case study | isolated hydrocephalus | situs inversus, micrognathia, ulnar deviation of the fingers with absent distal interphalangeal creases on fingers 2–4, lung abnormalities, and rocker-bottom feet | - | TES | Direct sequencing | AR | - |
| Mei et al., 2021 [137] | Genetic etiologies associated with infantile hydrocephalus in a Chinese infantile cohort | Children's Hospital of Fudan University, National Children's Medical Center, Shanghai, China | 110 Subjects, 300 Controls | Chinese | Case series | isolated hydrocephalus | - | - | WES | NGS; variant analysis | - | 11p15.4 (SBF2 mutations: c.G1171A, p.A391T in exon 12; c.A3877G, p.K1293E in exon 29; c.A3754T,<br>p.S1252C in exon 28;<br>c.A3056T, p.Q1019L in exon 24;<br>c.C5037T, p.R1679R in exon 36;<br>c.C1066T, p.R356X in exon 11;<br>c.G1067T, p.R356L in exon 11;<br>c.A2390G, p.Y797C in exon 20;<br>c.A2813G, p.E938G in exon 23;<br>c.T527G, p.L176W in exon 6;<br>c.A4328C, p.E1443A in exon 32);<br>10p11.22 (ZEB1 mutations:<br>c.444_461delinsG,<br>p.G150Wfs*3 in exon 4;<br>c.479_480delinsA,<br>p.N160Kfs*26 in exon 4;<br>c.G2995C, p.E999Q in exon 9);<br>3p21.31 (c.A465-8C in exon 5 of GNAI2) |
| Nabhan et al., 2014 [138] | Case Report: Whole-exome analysis of a child with polycystic kidney disease and ventriculomegaly | Kasr Al Ainy School of Medicine, Center of Pediatric Nephrology and Transplantation, Cairo University, Egyptian Group for Orphan Renal Diseases, Cairo, Egypt | 1 Subject, 2 Parents | - | Case study | Macrocephaly | bilateral enlarged and palpable kidneys, systemic hypertension | - | WES | Sanger sequencing , segregation analysis | AR | 6p12.3 (c.G3367A, p.G1123S in PKHD1);<br>17q22 (c.G368A, p.R123Q in MKS1) |
| Oud et al., 2016 [139] | A novel ICK mutation causes ciliary disruption and lethal endocrine- | Radboud Institute for Molecular Life | 4 Subjects, 2 Controls | Turkish | Case study | absence of septum pellucidum | genital anomalies, ventral septal defect, renal | - | WES; genotyping | IBD mapping; CNV; | - | 6p12.1 (c.G358T, p.G120C in ICK) |
|  | cerebro-osteodysplasia syndrome | Sciences, Radboud University Medical Centre, PO-Box 9101, 6500 HB Nijmegen, The Netherlands |  |  |  |  | abnormalities, cystic hygroma, scalp edema ascites, very short tubular bones and polydactyly of hands and feet, and short ribs |  |  | Sanger sequencing |  |  |
| Rocca et al., 2020 [140] | A novel genetic variant in DNAI2 detected by custom gene panel in a newborn with Primary Ciliary Dyskinesia: case report | University of Padova, Via Giustiniani, Padova, Italy | 1 Subject | Moroccan | Case study | isolated hydrocephalus | situs inversus, respiratory infections | Communicating | TGS, cytogenetics | NGS; aCGH; CNV | - | 17q25.1 (6.9kb deletion in of DNAI2) |
| Wallmeier et al., 2019 [141] | De Novo Mutations in FOXJ1 Result in a Motile Ciliopathy with Hydrocephalus and Randomization of Left/Right Body Asymmetry | University Children's Hospital Muenster, 48149 Muenster, Germany | 6 Subjects, Controls used | Germany, USA | Case series | isolated hydrocephalus | chronic destructive airway disease, and randomization of left/right body asymmetry | Obstructive | WES | Sequencing, unspecified | De novo | 17q25.1 (FOXJ1 mutations: c.G901T, p.E301X; c.868_871dup, p.T291Kfs*12; c.C826T, p.Q276X; c.967delG, p.E323Sfs*10; c.939delC, p.I314Sfs*19) |

#### PI3K-Akt-mTOR

Genes involved in PI3K-Akt-mTOR cell signaling pathway underlying HC are summarized in **Table 6**. Missense mutations were identified in ring finger protein 125 (*RNF125*), HECT and RLD domain containing E3 ubiquitin protein ligase family member 1 (*HERC1*), AKT serine/threonine kinase 3 (*AKT3*), mechanistic target of rapamycin kinase (*mTOR*), phosphatase and tensin homolog (*PTEN*), cyclin D2 (*CCND2*), phosphatidylinositol-4,5-bisphosphate 3-kinase catalytic subunit alpha (*PIK3CA*) (exon 18 and others), phosphoinositide-3-kinase regulatory subunit 2 (*PIK3R2*) (exon 13 and others), and platelet derived growth factor receptor beta (*PDGFRB*) (exon 12 and others). Deletions were observed in *PIK3CA*, and nonsense mutations were seen in *PTEN*. Deletions in chromosome 1 (1q42.3-q44) resulted in the deletion of AKT serine/threonine kinase 3 (*AKT3*). Additional genetic mutations implicated in this pathway included those in tripartite motif containing 71 (*TRIM71*), SWI/SNF related matrix associated, actin dependent regulator of chromatin (*SMARCC1*), forkhead box J1 (*FOXJ1*), formin 2 (*FMN2*), patched 1 (*PTCH1*), and FXYD domain containing ion transport regulator 2 (*FXYD2*). Multiple genes within the PI3K-AKT-MTOR pathway highlight convergence on molecular mechanisms conferring risk to HC. Murine models have demonstrated the role of *HERC1*, which codes for an E3 ubiquitin ligase, to affect Purkinje cell physiology and mTOR activity [54]. *TRIM71* and *SMARCC1* are expressed within the ventricles and epithelium of mice brains (determined via *in situ* hybridization) suggesting that a mutation within this gene locus can affect this region may lead to HC [9]. Mutations in *FOXJ1* and *FMN2* have been shown to alter neuroepithelial integrity and lead to HC in mice [55; 56]. Mice harboring mutations in *PTCH1* also display defects in ependymal cell integrity [57]. Thus, mutations within many genes converging on PI3K-Akt-mTOR signaling have been widely implicated in HC pathophysiology.

**Table 6:** PI3K-Akt-MTOR.

Abbreviations include the following: Array comparative genomic hybridization (aCGH).
| Citation | Title | Author Affiliation | Case # | Ancestry | Study Design | CNS Phenotype | Non-CNS Phenotype | Type of Hydrocephalus | Genetic Methodology | Genetic Analysis | Inheritance | Genetic Findings |
| --- | --- | --- | --- | --- | --- | --- | --- | --- | --- | --- | --- | --- |
| Cappuccio et al., 2019 [142] | Severe presentation and complex brain malformations in an individual carrying a CCND2 variant | Federico II University, Naples, Italy | 1 Subject, 2 Parents | - | Case study | infantile spasms, siezures, developmental delay, bilateral PMG, white matter hypoplasia, fenestration of the septum pellucidum and hypoplasia of the anterior and posterior commissures, hippocampal hypoplasia and malrotation, hypoplastic thalami and lentiform nuclei malrotation of the vermis, brainstem hypoplasia | bilateral postaxial polydactyly, patent foramen ovale and ductus arteriosus | - | TGS | NGS, sanger sequencing | De novo | 12p13.32 (c.C839T, p.T280I in CCND2) |
| Jin et al., 2020 [143] | Exome sequencing implicates genetic disruption of prenatal neuro-gliogenesis in sporadic congenital hydrocephalus | The Rockefeller University, New York, NY, USA | 381 Subjects , 1,798 Controls | - | Case-Control | Congenital hydrocephalus | - | Obstructive, Communicating | WES | CNV; sanger sequencing | De-novo | 3q26.32 (PIK3CA mutations: p.D350N; p.E365K; p.G914R; p.R770Q; p.N345S); 10q23.31 (PTEN mutations: p.Y16X; p.R130Q; p.R335X; p.S305N); 1p36.22 (MTOR mutations: p.E1799K; p.M304T; p.R769C; p.R1161G; p.R1170C; p.H1782R); Mutations in 3p22.3 (TRIM71), 3p21.31 (SMARCC1), 17q25.1 (FOXJ1), 1q43 (FMN2), 9q22.32 (PTCH1) and 11q23.3 (FXDY2) |
| Maguolo et al., 2018 [144] | Clinical pitfalls in the diagnosis of segmental overgrowth syndromes: a child with the c.2740G > A mutation in PIK3CA gene | University Hospital of Verona, Verona, Italy | 1 Subject | Italian | Case study | cerebellar tonsillar ectopia, a markedly thick corpus callosum, and white matter abnormalities | lateralized overgrowth (segmental overgrowth syndrome) | - | TGS | Targeted NGS | - | 3q26.32 (c.G2740A, pG914R in exon 18 of PIK3CA) |
| Maini et al., 2018 [145] | A Novel CCND2 Mutation in a Previously Reported Case of Megalencephaly and Perisylvian Polymicrogyria with Postaxial Polydactyly and Hydrocephalus | Azienda Unità Sanitaria Locale, Arcispedale Santa Maria Nuova, IRCCS, Reggio Emilia, Italy | 1 Subject, Controls used | - | Case study | intellectual disability, seizures | aphasia, postaxial polydactyly | - | WES | Sanger sequencing, direct sequencing | - | 12p13.32 (c.C839T, p.T280I in CCND2) |
| McDermott et al., 2018 [146] | Hypoglycaemia represents a clinically significant manifestation of PIK3CA- and CCND2-associated segmental overgrowth | St Mary's Hospital, Central Manchester University Hospitals, NHS Foundation Trust Manchester Academic Health Sciences Centre, Manchester, UK | 6 Subjects | - | Case series | Polymicrogyria | polydactyly, capillary malformation, endocrine abnormalities | - | TGS | NGS; sanger sequencing | - | 3q26.32 (PIK3CA mutations: c.G1048A, p.D350N; c.G2176A, p.E726K; c.G263A, p.R88Q); 12p13.32 (c.C841G, p.P281R in CCND2) |
| Mirzaa et al., 2015 [147] | Characterisation of mutations of the phosphoinositide-3-kinase regulatory subunit, PIK3R2, in perisylvian polymicrogyria: a next-generation sequencing study | University of Washington, Seattle, WA, USA; Center for Integrative Brain Research, Seattle Children's Research Institute, Seattle, WA, USA | 20 Subjects, Controls used | USA | Case series | polymicrogyria, seizures | oromotor weakness | - | WES | AD assay; smMIPs; amplicon sequencing; sanger sequencing | De novo, maternal, | 19p13.11 (c.G1117A, p.G373R and c.A1126G, p.K376E in PIK3R2) |
| Mirzaa, et al. 2013 [148] | Megalencephaly syndromes and activating mutations in the PI3K-AKT pathway: MPPH and MCAP | Center for Integrative Brain Research, University of Washington, Seattle Children's Research Institute, Seattle, WA, USA | 50 Subjects | - | Case series | cerebellar tonsillar ectopia or Chiari malformation, cortical brain abnormalities, macrocephaly | postaxial polydactyly | - | WES | Sanger sequencing; REF; targeted ultra-deep sequencing | De novo | 19p13.11 (p.G373R in PIK3R2); 1q43-q44 (p.R465W and p.N229S in AKT3); 3q26.32 (PIK3CA mutations: c.G241A, p.E81K; c.G263A, p.R88Q; c.G1090A, p.G364R; c.G1093A, p.E365K; c.G1133A, p.C378Y; c.1359_1361del, p.E453del; c.G1633A, p.E545K; c.G2176A, p.E726K; |
|  |  |  |  |  |  |  |  |  |  |  |  | c.G2740A, p.G914R; c.A3062G, p.Y1021C; c.A3073G, p.T1025A; c.C3104T, p.A1035V; c.G3129T, p.M1043I; c.C3139T, p.H1047Y; c.G3145A, p.G1049S) |
| Ortega-Recalde et al., 2015 [149] | Biallelic HERC1 mutations in a syndromic form of overgrowth and intellectual disability | Escuela de Medicina y Ciencias de la Salud, Universidad del Rosario, Bogotá, Colombia | 2 Subjects | Colombian | Case study | intellectual disability | overgrowth, kyphoscoliosis and facial dysmorphism | Communicating | WES | NGS, sanger sequencing | AR | 15q22.31 (c.G2625A, p.W875X and c.G13559A, p.G4520E in HERC1) |
| Poduri et al., 2012 [150] | Somatic activation of AKT3 causes hemispheric developmental brain malformations | Children's Hospital Boston, 300 Longwood Avenue, Boston, MA 02115, USA | 8 Subjects, Controls used | - | Case series | intellectual disability and severe, intractable epilepsy | - | - | TGS | CNV; SNP; Karyotyping | *De novo* | 1q43-q44 (c.G49A, p.E17K in AKT3) |
| Riviere et al., 2012 [151] | De novo germline and postzygotic mutations in AKT3, PIK3R2 and PIK3CA cause a spectrum of related megalencephaly syndromes | Seattle Children's Hospital, Seattle, Washington, USA | 52 Subjects, 95 Controls | European | Case series | megalocephaly, variable cortical malformation | growth dysregulation with variable asymmetry, developmental vascular anomalies, distal limb malformations (syndactyls and polydactyls), and a mild connective tissue dysplasia | - | TES | Sanger sequencing; REF; targeted deep sequencing | *De novo* | 1q43-q44 (c.C1393T, p.R465W and c.A686G, p.N229S in AKT3); 19p13.11 (c.G1117A; p.G373R in PIK3R2); 3q26.32 (PIK3CA mutations: c.G241A, p.E81K; c.G263A, p.R88Q; c.G1090A, p.G364R; c.G1093A, p.E365K; c.G1133A, p.C378Y; c.1359_1361del, p.E453del; c.G1633A, p.E545K; c.G2176A, p.E726K; c.G2740A, p.G914R; c.A3062G, p.Y1021C; c.A3073G, p.T1025A;<br>c.C3104T,<br>p.A1035V;<br>c.G3129T,<br>p.M1043I;<br>c.C3139T,<br>p.H1047Y;<br>c.G3145A,<br>p.G1049S) |
| Sameshi<br>ma et al.,<br>2019<br>[152] | MPPH syndrome<br>with aortic<br>coarctation and<br>macrosomia due<br>to CCND2<br>mutations | Hyogo<br>Prefectural<br>Awaji<br>Medical<br>Center,<br>Sumoto,<br>Hyogo,<br>Japan | 1<br>Subject,<br>2<br>Parents | Japanese | Case<br>study | polymicrogyria,<br>seizures | forehead<br>protrusion,<br>sacral cusp<br>depression,<br>low auricle,<br>depressed<br>nasal bridge<br>and postaxial<br>polydactyly,<br>aortic<br>coarctation | - | TGS | NGS,<br>sanger<br>sequencing | - | 12p13.32<br>(c.C842G,<br>p.P281R in<br>CCND2) |
| Szalai et<br>al., 2020<br>[153] | Maternal<br>mosaicism<br>underlies the<br>inheritance of a<br>rare germline<br>AKT3 variant<br>which is<br>responsible for<br>megalencephaly-polymicrogyria-polydactyly-hydrocephalus<br>syndrome in two<br>Roma half-<br>siblings | University<br>of Pecs,<br>Medical<br>School,<br>Departmen<br>t of Medical<br>Genetics,<br>Pecs,<br>Hungary | 2<br>Subjects | Hungarian<br>Roma | Case<br>study | intellectual disability,<br>epilepsy, brain<br>malformations, and<br>megalencephaly | dysmorphic<br>features,<br>visual<br>impairment | - | WES,<br>cytogenetics | Karyotyping<br>; aCGH;<br>sanger<br>sequencing | Maternal<br>mosaicism | 1q43-q44<br>(c.C1393T,<br>p.R465W in AKT3) |
| Tapper<br>et al., 2014<br>[154] | Megalencephaly<br>syndromes:<br>exome pipeline<br>strategies for<br>detecting low-level mosaic<br>mutations | University<br>of Southampt<br>on,<br>Southampt<br>on,<br>Hampshire,<br>United<br>Kingdom ;<br>Wessex<br>Regional<br>Genetics<br>Laboratory,<br>Salisbury<br>District<br>Hospital,<br>Salisbury,<br>Wiltshire,<br>United<br>Kingdom | 3<br>Subjects<br>, 4<br>Parents | - | Case<br>series | macrocephaly,<br>dysmorphic<br>cerebellum, hypotonia | capillary<br>malformation<br>s,<br>overgrowth<br>and<br>asymmetry,<br>development<br>al delay | - | WES,<br>cytogenetics | aCGH;<br>sanger<br>sequencing | - | 3q26.32<br>(c.G2176A,<br>p.E726K in<br>PIK3CA);<br>19p13.11<br>(c.G1117A,<br>p.G373R in<br>PIK3R2) |
| Tenorio<br>et al., 2014<br>[155] | A new<br>overgrowth<br>syndrome is due<br>to mutations in<br>RNF125 | Hospital<br>Universitari<br>o La Paz,<br>Universida<br>d<br>Autónoma<br>de Madrid<br>(UAM), | 6<br>Subjects<br>, 350<br>Control | Spanish | Case<br>series | macrocephaly,<br>intellectual disability | overgrowth,<br>hypoglycemi<br>a,<br>inflammatory<br>diseases<br>resembling<br>Sjögren<br>syndrome | - | TGS,<br>cytogenetics | Karyotyping<br>; aCGH;<br>SNP array;<br>MLPA;<br>high-<br>resolution<br>melting;<br>sanger<br>sequencing; | *De novo* | 18q12.1 (RNF125<br>mutations:<br>c.G336A, p.M112I;<br>c.C488T, p.S163L;<br>c.C520T,<br>p.R174C) |
|  |  | Madrid,<br>Spain |  |  |  |  |  |  |  | pyrosequencing |  |  |
| Terrone et al., 2016 [156] | De novo PIK3R2 variant causes polymicrogyria, corpus callosum hyperplasia and focal cortical dysplasia | Federico II University, Naples, Italy | 1 Subject | Italian | Case study | left spastic hemiplegia, megalencephaly, perisylvian polymicrogyria, and mega corpus callosum | synophrys, depressed nasal bridge, anteverted nares, pectus excavatum, broad thumb and hallux | - | WES | Sanger sequencing | *De novo* | 19p13.11 (c.G1669C, p.D557H in exon 13 of PIK3R2) |
| Zarate et al., 2019 [157] | Constitutive activation of the PI3K-AKT pathway and cardiovascular abnormalities in an individual with Kosaki overgrowth syndrome | University of Arkansas for Medical Sciences, Little Rock, Arkansas | 1 Subject, 1 Control | - | Case study | Dandy-Walker malformation, cervical spine arachnoid cyst, progressive scoliosis, white matter lesions, spastic diplegia | craniofacial dysmorphism, hyperextensible skin, cardiac saccular aneurysms, developmental delay, low-frequency hearing loss | Obstructive | TES | Exome sequencing trio analysis, sanger sequencing | *De novo* | 5q32 (c.T1696C, p.W566R in exon 12 of PDGFRB) |

### Vesicle Regulation & Cell Adhesion

**Table 7** details mutations in genes responsible for vesicle regulation and cell adhesion that contribute to the development of HC. Missense mutations were found in and glial fibrillary acidic protein (*GFAP*). Sorting nexin 10 (*SNX10*) displayed a nonsense mutation and clathrin heavy chain (*CLTC*) displayed a frameshift mutation. Additional mutations include ArfGAP with FG repeats 1 (*RAB*), multiple PDZ domain crumbs cell polarity complex component (*MPDZ*), beta 1,3-glucosyltransferase (*B3GALTL*), SEC24 homolog D, COPII coat complex component (*SEC24D*), and actin beta (*ACTB*).

**Table 7:** Vesicle Regulation & Cell Adhesion.

Abbreviations include the following: Array comparative genomic hybridization (aCGH).
| Citation | Title | Author Affiliation | Case # | Ancestry | Study Design | CNS Phenotype | Non-CNS Phenotype | Type of Hydrocephalus | Genetic Methodology | Genetic Analysis | Inheritance | Genetic Findings |
| --- | --- | --- | --- | --- | --- | --- | --- | --- | --- | --- | --- | --- |
| Al-Dosari et al., 2013 [158] | Mutation in MPDZ causes severe congenital hydrocephalus | King Faisal Specialist Hospital and Research Center, Riyadh, Saudi Arabia | 1 Subject, 50 Controls | Saudi | Case series | callosal agenesis, hypotonia | chorioretinal coloboma, atrial septal defect | Communicating | TGS, genotyping | Autozygosity mapping, linkage analysis, sanger sequencing | AR | 9p23 (MPDZ) |
| Al-Jezawi et al., 2018 [159] | Compound heterozygous variants in the multiple PDZ domain protein (MPDZ) cause a case of mild non-progressive communicating hydrocephalus | College of Medicine and Health Sciences, United Arab Emirates University | 1 Subject, 2 Parents, 100 Controls | United Arab Emirates | Case study | Isolated hydrocephalus | large head with frontal bossing and high arched palate | Communicating | WES | Variant analysis, sanger sequencing | AR | 9p23 (MPDZ) |
| DeMari et al., 2016 [160] | CLTC as a clinically novel gene associated with multiple malformations and developmental delay | SUNY Upstate Medical University, Syracuse, New York | 1 Subject, 2 Parents | Caucasian | Case study | Hypotonia | prominent jaw, large anterior fontanel, bilateral hip laxity, and jaundice, low-set ears, depressed nasal bridge, anteverted nares, widely set involutioned nipples | Communicating | WGS, cytogenetics | Karyotype, SNP microarray, co-segregation analysis, sanger sequencing | De novo | 17q23.1 (A heterozygous de novo frameshift mutation, c.2737_2738dupGAp.D913Efs*59) |
| Mégarbane et al., 2013 [161] | Homozygous stop mutation in the SNX10 gene in a consanguineous Iraqi boy with osteopetrosis and corpus callosum hypoplasia | Unité de Génétique Médicale et laboratoire associé INSERM à l'Unité UMR_S 910, Pôle Technologie Santé, Université Saint-Joseph, Beirut, Lebanon | 1 Subject, 1 Control | Iraqi | Case study | macrocephaly, brain atrophy, thin corpus callosum | proptosis of the eyes, skeletal abnormality, strabismus, splenomegaly and joint hyperlaxity | Communicating | TGS | Direct sequencing | AR | 7p15.2 (SNX10 gene) |
| Rajadhyax et al., 2007 [162] | Neurological presentation of Griscelli syndrome: obstructive hydrocephalus without haematological abnormalities or organomegaly | Genetics and Neurosurgery, Leeds General Infirmary, UK | 1 Subject | Asian | Case study | sixth nerve palsy, increased muscle tone | patchy hyperpigmentation on the lower limbs, hemophagocytic lymphohistiocytosis | Obstructive | TGS | - | AR | 2q36.3 (RAB27A) |
| Reis et al., 2008 [163] | Mutation analysis of B3GALT1 in Peters Plus syndrome | Medical College of Wisconsin and Children's Hospital of Wisconsin, Milwaukee, Wisconsin, USA | 8 Subjects, 180 Controls | Dutch | Case series | intellectual disability | central corneal opacity, defects in the posterior layers of the cornea, and lenticulo-corneal and/or irido-corneal adhesions, short stature, short broad hands with fifth finger clinodactyly, distinctive facial features, cleft lip and/or cleft palate, hearing loss, abnormal ears, heart defects, genitourinary anomalies | Communicating | TGS | Direct sequencing, | AR | 13q12.3 (beta1,3-glucosyltransferase gene (B3GALT1)) |
| Rodriguez et al., 2001 [164] | Infantile Alexander disease: spectrum of GFAP mutations and genotype-phenotype correlation | Laboratoire de Neurogénétique Moléculaire, INSERM U546, Université Paris VI, France | 15 Subjects, 50 Controls | - | Case series | macrocephaly, psychomotor regression, seizures, and spasticity | respiratory difficulties | Communicating | TES | - | *De novo* | 17q21.31 (Missense, heterozygous, de novo GFAP mutations (R79H; four had R239C; and one had R239H)) |
| Sakakibara et al., 2007 [165] | A case of infantile Alexander disease diagnosed by magnetic resonance imaging and genetic analysis | Nara Medical University, Japan | 1 Subject | - | Case study | megalencephalic, seizures, white matter abnormalities | bulbar paralysis | Obstructive | TGS | - | AD | 17q21.31 (R239H mutation of glial fibrillary acidic protein(GFAP)) |
| Saugier-Weber et al., 2017 [166] | Hydrocephalus due to multiple ependymal malformations is caused by mutations in the MPDZ gene | Normandie Univ, UNIROUEN, INSERM U1245, Normandy Centre for Genomic and Personalized Medicine, Rouen University Hospital, F76000, Rouen, France | 5 Subjects, 3 Controls | - | Case series | Multifocal ependymal malformations | - | Obstructive | TGS, cytogenetics | Karyotyping, variant analysis, sanger sequencing, targeted NGS | AR | 9p23 (MPDZ gene) |
| Takeyari et al., 2018 [167] | Japanese patient with Cole-carpen-ter syndrome with compound heterozygo- us variants of SEC24D | Osaka University Graduate School of Medicine, Osaka, Japan | 1 Subject | Japanese | Case study | craniosynostosis | prominent eye and micrognathia, short neck, scoliosis, and chest deformity, bone fractures, Wormian bones, lordosis, and long thin bones | - | TES | Variant analysis, sanger sequencin- g | - | 4q26 (SEC24D) |
| Van der Knaap et al., 2005 [168] | Unusual variants of Alexander's disease | VU University Medical Center, De Boelelaan 1117, 1081 HV Amsterdam, the Netherlands | 10 Subjects, 100 Controls | - | Case series | cerebral white matter abnormalities, brainstem lesions | scoliosis, dysphagia, gait disturbances | Obstructive | TGS | - | *De novo* | 17q21.31 (GFAP) |
| Zhang et al., 2020 [169] | Prenatal presentatio- n and diagnosis of Baraitser- Winter syndrome using exome sequencing | Virginia Tech Carilion School of Medicine, Roanoke, Virginia, USA | 1 Subject, 2 Parents | - | Case study | interhemispheric cyst | cystic hygroma and omphalocele, ocular coloboma, hypertelorism, heart, renal, musculoskeletal system defects | - | TGS | NGS, variant analysis, | AD | 7p22.1 (ACTB) |

GFAP is required for white-matter architectural development and myelination, perhaps accounting for the neurodevelopmental comorbidities frequently observed in patients with HC [58]. Mutations in the phosphoinositide binding domain of *SNX10* alters endosomal integrity, suggesting a potential pathogenic mechanism in vesicular trafficking [59]. Additionally, mutations in this gene locus can disrupt interactions between sorting nexins and the V-ATPase complex further contributing to vesicle dysfunction and ciliopathy [60]. *CLTC* contributes to the development of the vesicular coat, and a mutation within this gene locus may disrupt vesicle stability [61]. Mutations in *RAB27A* are associated with Griscelli syndrome, characterized by albinism, hematological abnormalities, and organ malformation which can also present with HC [62]. *SEC24* is also involved in intracellular trafficking by interacting with export signals from the endoplasmic reticulum and regulating cargo transport [63]. In addition, *MPDZ* is highly expressed in tight junctions suggesting that a mutation within this gene locus may disrupt alter tissue permeability [64]. Finally, *B3GALTL* interacts with the thrombospondin type 1 repeat (TSR) protein family which play varied roles in maintain and regulating cell-cell adhesion [65].

### Glycosylation defects

**Table 8** summarizes genes implicated in human HC associated with defects in glycosylation. Nonsense mutations were seen in protein O-mannose kinase (*POMK*), and protein O-mannosyltransferase 1 (*POMT1*). Loss of function mutations were identified in dystroglycan 1 (*DAG1*) and isoprenoid synthase domain containing gene (*ISPD*). Additional mutations included those in protein C, inactivator of coagulation factors Va and VIIIa (*PROC*), fukutin related protein (*FKRP*), protein O-mannosyltransferase 2 (*POMT2*), protein O-linked mannose N-acetylglucosaminyltransferase 1 (beta 1,2-) (*POMGNT1*), LARGE xylosyl and glucuronyltransferase 1 (*LARGE1*) and a translocation between chromosome 5 and 6, t(5;6) (q35;q21). In addition, *DAG1* codes for dystroglycan, a protein involved in extracellular matrix integrity and the genetic etiology of many neurological syndromes. Mutations in *DAG1* have been found to contribute to Walker-Warburg syndrome and other muscular dystrophy-dystroglycanopathies which can be associated with HC [66]. Dystroglycan may also be affected through defects in its glycosylation patterns. For instance, mutations in *POMK* have been shown to impair the glycosylation of a-dystroglycan affecting cytoskeleton stability [67]. Other genes contributing to dystroglycanopathies through glycosylation errors include *POMT1*, *POMT2*, *POMGNT1*, *FKRP*, *ISPD* and *LARGE1* [68].

**Table 8:** Glycosylation defects.

Abbreviations include the following: Array comparative genomic hybridization (aCGH).
| Citation | Title | Author Affiliation | Case # | Ancestry | Study Design | CNS Phenotype | Non-CNS Phenotype | Type of Hydrocephalus | Genetic Methodology | Genetic Analysis | Inheritance | Genetic Findings |
| --- | --- | --- | --- | --- | --- | --- | --- | --- | --- | --- | --- | --- |
| Beltran-Valero de Bernabé et al., 2004 [170] | Mutations in the FKRP gene can cause muscle-eye-brain disease and Walker-Warburg syndrome | University Medical Centre Nijmegen, Nijmegen, The Netherlands | 2 Patients, 200 Controls | German, Asian | Case series | dandy walker-like malformation , intellectual disability | muscular dystrophy, left ventricular hypertrophy, retinal and eye developmental issues | Communicating | TES | Direct sequencing, linkage analysis | - | 19q13.32 (FKRP) |
| Beltrán-Valero de Bernabé et al., 2002 [171] | Mutations in the O-mannosyltransferase gene POMT1 give rise to the severe neuronal migration disorder Walker-Warburg syndrome | University Medical Centre Nijmegen, Nijmegen, The Netherlands | 30 Subjects, 105 Controls | Turkish, Italy, Dutch, Australian | Case series | cobblestone lissencephaly , occipital encephalocele | eye malformations , congenital muscular dystrophy or elevated creatine kinase | Obstructive | TES | Linkage analysis, SSCP, restriction enzyme analysis | AR | 9q34.13 (POMT1) |
| Biancheri et al., 2006 [172] | POMGnT1 mutations in congenital muscular dystrophy: genotype-phenotype correlation and expanded clinical spectrum | University of Genova, Italy | 3 Subjects, 192 Controls | Italian | Case series | intellectual disability, epilepsy, and lissencephaly | congenital muscular dystrophy, ocular abnormalities | Communicating | TGS | Direct sequencing | AR | 1p34.1 (POMGnT1) |
| Bouchet et al., 2007 [173] | Molecular heterogeneity in fetal forms of type II lissencephaly | Bichat-Claude Bernard Hospital, Biochimie Métabolique, Paris, France | 47 Subjects, 100 Controls | French | Case series | agyria, thick leptomeninges, disorganized cortical ribbon, cerebellar dysplasia | - | Communicating | TGS | - | AR | 9q34.13 (15 in POMT1); 14q24.3 (five in POMT2); 1p34.1 (POMGNT1) |
| Cormand et al., 2001 [174] | Clinical and genetic distinction between Walker-Warburg syndrome and muscle-eye-brain disease | University of Helsinki, Finland | 29 Subjects | Turkish, Netherlands, German, Pakistani, Swedish, Palestinian, Dutch, and American | Case series | malformation of neuronal migration compatible with cobblestone complex | elevated serum creatine kinase level or abnormal muscle biopsy, and ocular abnormalities | - | Genotyping | Linkage analysis | AR | MEB gene locus localized to 1p32-p34 |
| Currier et al., 2005 [175] | Mutations in POMT1 are found in a minority of patients with Walker-Warburg syndrome | Beth Israel Deaconess Medical Center, Boston, Massachusetts, USA | 30 Subjects, 110 Controls | Asian, African, and Caucasian | Case series | cerebellar hypoplasia, brainstem hypoplasia, agenesis of the corpus callosum, agenesis of the septum pellucidum, interhemispheric fusion, and the presence of an encephalocel<br>e | ocular abnormalities, congenital muscular dystrophy | Obstructive | TES | Microsatellite marker assay | AR | 9q34.13 (POMT1) |
| Geis et al.,<br>2019 [176] | Clinical long-time course, novel mutations and genotype-phenotype correlation in a cohort of 27 families with POMT1-related disorders | Klinik St. Hedwig, University Children's Hospital Regensburg (KUNO), Steinmetzstr. 1-3, 93049, Regensburg, Germany | 35 Subjects | German, Turkish, Indonesian, Gipsy, African | Case series | lissencephaly type II, hypoplasia of the pons and/or brainstem, cerebellar hypoplasia, hypoplasia of the corpus callosum, encephalocel<br>e | muscle weakness, muscular dystrophy, GI malformations | Communicating | TGS | Direct sequencing, sanger sequencing, massive parallel sequencing | AR | 9q34.13 (POMT1) |
| Godfrey et al.,<br>2007 [177] | Refining genotype phenotype correlations in muscular dystrophies with defective glycosylation of dystroglycan | Hammersmith Hospital, Imperial College, London, UK | 92 Subjects | Australia, Turkey | Case series | cobblestone lissencephaly | limb girdle muscular dystrophy, Congenital muscular dystrophy, elevated serum CK | Communicating | TGS | Unidirectional sequencing, HA, segregation analysis, | AR, *De novo* | 9q34.13 (POMT1); 14q24.3 (POMT2); 1p34.1 (POMGnT1); 9q31.2 (FKTN); and 22q12.3 (LARGE) |
| Hehr et al.,<br>2007 [178] | Novel POMGnT1 mutations define broader phenotypic spectrum of muscle-eye-brain disease | University of Regensburg, Universitätsklinikum D3, Franz-Josef-Strauss-Allee 11, Regensburg, Germany | 9 Subjects | German, Turkish, English | Case series | global developmental delay, seizures, cerebellar cysts, intellectual disability | congenital muscular dystrophy, severe congenital myopia, glaucoma, retinal hypoplasia | Communicating | TGS | Cycle sequencing, linkage analysis, restriction enzyme analysis | AR | 1p34.1 (POMGnT1) |
| Ichiyama et al.,<br>2016 [179] | Fetal hydrocephalus and neonatal stroke as the first presentation of protein C deficiency | Kyushu University, Fukuoka, Japan | 1 Subject | Asian | Case study | isolated hydrocephalus | Slight developmental delay | - | TES | Direct sequencing | - | 2q14.3 (PROCc.574_576 delAAG) |
| Kano et al.,<br>2002 [180] | Deficiency of alpha-dystroglycan in muscle-eye-brain disease | Osaka University Graduate School of Medicine, 2-2 B9, Yamadaoka, Suita, Osaka, Japan | 3 Subjects, 1 Control | Turkish, French | Case series | type II lissencephaly, Intellectual disability | congenital muscular dystrophy, congenital myopia, congenital glaucoma, pallor of the optic discs, retinal hypoplasia, hydrocephalus, myoclonic jerks | Communicating | TGS | - | AR | 1p34.1 (POMGnT1) |
| Karadeniz et al.,<br>2002 [181] | De novo translocation t(5;6)(q35;q21) in an infant with Walker-Warburg syndrome | Burak Woman's Hospital, Department of Medical Genetics, | 1 Subject, 2 Parents | - | Case study | hypoplasia of cerebellar vermis, enlargement of cisterna magna, | eye abnormalities with microphthalmia cataract, congenital | Communicating | - | G-banding | *De novo* | translocation t(5;6)(q35;q21) |
|  |  | Ankara,<br>Turkey |  |  |  | bilateral<br>dilatation of<br>lateral<br>ventricles,<br>widespread<br>agyria, and<br>irregularity of<br>the white<br>matter-gray<br>matter line | muscular<br>dystrophy |  |  |  |  |  |
| Preiksaitiene<br>et al., 2020<br>[182] | Pathogenic homozygous<br>variant in POMK gene is the<br>cause of prenatally detected<br>severe ventriculomegaly in two<br>Lithuanian families | Vilnius<br>University,<br>Vilnius,<br>Lithuania | 4<br>Subjects,<br>98<br>Controls | Lithuania<br>n | Case<br>series | Isolated<br>hydrocephalus | highly<br>variable | Dependent on<br>phenotype of<br>dystroglycanopathy | WES | Sanger<br>sequencing | *De novo*,<br>AR | 8p11.21<br>(homozygous<br>nonsense<br>variant in the<br>POMK) |
| Van Reeuwijk<br>et al., 2005<br>[183] | POMT2 mutations cause<br>alpha-dystroglycan<br>hypoglycosylation and Walker-Warburg syndrome | Radboud<br>University<br>Nijmegen<br>Medical<br>Centre,<br>Nijmegen,<br>The<br>Netherlands | 3<br>Subjects,<br>Controls<br>used | Moroccon,<br>Pakistani,<br>Bengali | Case<br>series | lissencephaly<br>, agenesis of<br>the corpus<br>callosum,<br>fusion of the<br>hemispheres,<br>cerebellar<br>hypoplasia,<br>and neuronal<br>overmigration | eye<br>malformations<br>(cataract,<br>microphthalmia,<br>buphthalmos,<br>and Peters<br>anomaly) | - | TGS | Homozygosity<br>mapping,<br>direct<br>sequencing | AR | 14q24.3<br>(POMT2) |
| Van Reeuwijk<br>et al., 2006<br>[184] | The expanding phenotype of<br>POMT1 mutations: from<br>Walker-Warburg syndrome to<br>congenital muscular dystrophy,<br>microcephaly, and mental<br>retardation | Radboud<br>University<br>Nijmegen<br>Medical<br>Centre,<br>Nijmegen,<br>The<br>Netherlands | 28<br>Subjects,<br>100<br>Controls | Italy,<br>Netherlands,<br>Pakistan,<br>Lebanon,<br>India,<br>Qatar,<br>Ireland,<br>turkey | Case<br>series | lissencephaly<br>, agenesis of<br>the corpus<br>callosum,<br>fusion of the<br>hemispheres,<br>cerebellar<br>hypoplasia,<br>and neuronal<br>overmigration | myopia, gait<br>disturbances | Communicating | TGS | Linkage<br>analysis | - | 9q34.13<br>(POMT1) |
| Van Reeuwijk<br>et al., 2010<br>[185] | A homozygous FKRP start<br>codon mutation is associated<br>with Walker-Warburg<br>syndrome, the severe end of<br>the clinical spectrum | Radboud<br>University<br>Nijmegen<br>Medical<br>Centre,<br>Nijmegen,<br>The<br>Netherlands | 2<br>Subjects,<br>2 Parents | Caucasian | Case<br>series | lissencephaly<br>, agenesis of<br>the corpus<br>callosum,<br>fusion of the<br>hemispheres,<br>cerebellar<br>hypoplasia,<br>and neuronal<br>overmigration | cataracts,<br>muscular<br>dystrophy | Communicating | TGS | SNP | - | 19q13.32<br>(FKRP) |
| Riemersma et<br>al., 2015 [186] | Absence of α- and β-<br>dystroglycan is associated with<br>Walker-Warburg syndrome | Leiden<br>University<br>Medical<br>Center, the<br>Netherlands,<br>Sydney<br>Children's<br>Hospital,<br>University of<br>New South<br>Wales,<br>Sydney, | 5<br>Subjects,<br>Controls<br>used | Israeli-Arab | Case<br>series | hypotonia,<br>posterior<br>fossa, a<br>small midline<br>encephalocele, a<br>hypoplastic<br>vermis,<br>intracranial<br>calcifications | elevated CK,<br>elevated<br>LFTs,<br>respiratory<br>failure,<br>bilateral<br>corneal<br>opacities, and<br>glaucoma | Communicating | TES | Homozygosity<br>mapping,<br>CNV,<br>sanger<br>sequencing | - | 3p21.31<br>(homozygous<br>loss-of-function<br>frameshift<br>mutation in the<br>DAG1<br>gene) |
|  |  | Australia, Rambam Health Care Campus, Haifa, Weizmann Institute of Science, Rehovot, Israel |  |  |  |  |  |  |  |  |  |  |
| Saredi et al., 2012 [187] | Novel POMGNT1 point mutations and intragenic rearrangements associated with muscle-eye-brain disease | Foundation Neurological Institute C. Besta, Milano, Italy | 3 Subjects, 1 Control | Italian | Case series | Microcephaly, spastic tetraparesis | rounded forehead, thin lips, short neck, micrognathia, motor disability, eye abnormalities | Communicating | TGS | Cycle sequencing, MLPA | AR | 1p34.1 (c.643C>T, c.1863del C in POMGNT1) |
| Vervoort et al., 2004 [188] | POMGNT1 gene alterations in a family with neurological abnormalities | J. C. Self Research Institute of Human Genetics, Greenwood Genetic Center, Greenwood, SC, USA | 2 Subjects, 2 Parents, 500 Controls | Caucasian | Case series | hypotonia, bilateral frontal polymicrogyria, abnormal cerebellum, and characteristic flattened dystrophic pons | congenital muscular dystrophy, congenital glaucoma and severe myopia | - | TGS | Haplotype analysis, SSCP, cycle sequencing | AR | 1p34.1 (POMGNT1) |
| Willer et al., 2012 [189] | ISPD loss-of-function mutations disrupt dystroglycan O-mannosylation and cause Walker-Warburg syndrome | University of Iowa Roy J and Lucille A Carver College of Medicine, Iowa City, Iowa, USA | 7 Subjects, Controls used | - | Case series | cobblestone lissencephaly, severe brainstem hypoplasia with a kink at the isthmus and severe hypoplasia of the cerebellum | muscular dystrophy, bilateral microphthalmia with cataracts and arrested retinal development | Communicating | TGS, cytogenetics | Linkage analysis, targeted NGS, aCGH, CNV, Sanger sequencing | AR | 7p21 (ISPD mutation) |
| Yis et al., 2007 [190] | A case of Walker-Warburg syndrome resulting from a homozygous POMT1 mutation | University of Dokuz Eylul, 35340 Izmir, Turkey | 1 Subject, 2 Parents | - | Case study | type II lissencephaly and pontocerebellar hypoplasia | severe ocular malformations and congenital muscular dystrophy | Communicating | TGS | Linkage analysis, direct sequencing | AR | 9q34.13 (mutation (R514X) in the POMT1 gene) |
| Yoshida et al., 2001 [191] | Muscular dystrophy and neuronal migration disorder caused by mutations in a glycosyltransferase, POMGNT1 | Central Laboratories for Key Technology, Kirin Brewery Co., Ltd., Kanazawa-ku, Yokohama, Japan | 6 Subjects | Turkish, French | Case series | lissencephaly | congenital muscular dystrophy, ocular abnormalities | Communicating | TGS | Direct sequencing | AR | 1p34.1 (POMGNT1) |

### Growth factor related signaling

**Table 9** summarizes genetic mutations associated with growth factor related signaling dysfunction. Mutations were observed in fibroblast growth factor receptor 1 (*FGFR1*), fibroblast growth factor receptor 2 (*FGFR2*), fibroblast growth factor receptor 3 (*FGFR3*), ZPR1 zinc finger (*ZPR1*), and fibrillin 1 (*FBN1*). Specifically, exon 7 displayed a missense mutation in *FGFR2* and exon 64 displayed a mutation in *FBN1*. Mutations in *FGFR* play pleiotropic roles in numerous syndromes including Crouzon syndrome, Jackson-Weiss syndrome, Apert syndrome and Pfeiffer syndrome [69; 70; 71; 72]. These craniosynostoses have been associated with HC and *FGFR* mutations contributing to bony abnormalities, which may explain the venous and CSF outflow obstructions leading to this phenotype [73]. The FGFR mutations identified are predominantly gain of function mutations altering ligand binding and tyrosine kinase activity [74]. In addition, *ZPR1* contributes to cell proliferation and *FBN1* is associated with TGF beta signaling suggesting their mechanistic contributions to the HC phenotype seen in patients with these phenotypes [75].

**Table 9:** Growth factor signaling.

Abbreviations include the following: Autosomal Dominant (AD).
| Citation | Title | Author Affiliation | Case # | Ancestry | Study Design | CNS Phenotype | Non-CNS Phenotype | Type of Hydrocephalus | Genetic Methodology | Genetic Analysis | Inheritance | Genetic Findings |
| --- | --- | --- | --- | --- | --- | --- | --- | --- | --- | --- | --- | --- |
| Abdel-Salam et al., 2011 [192] | Muenke syndrome with pigmentary disorder and probable hemimegalencephaly: An expansion of the phenotype | National Research Centre, Cairo, Egypt | 1 Patient, 2 parents | - | Case study | left HME, inadequate differentiation of white and gray matter, underdeveloped corpus callosum, abnormal hippocampus configuration, right coronal, sagittal, and lambdoid suture synostoses | frontal bossing, sparse, hypopigmented, curly hair, prominent eyes, low-set ears, hypoplastic maxilla, long philtrum, brachydactyly with fusiform fingers, skin hyperpigmentation | Obstructive | TES | MLPA, DHPLC | AD | 4p16.3 (FGFR3 showed a c.749C > G, p.Pro250Arg substitution) |
| Arnaud-López et al., 2007 [193] | Crouzon with acanthosis nigricans. Further delineation of the syndrome | Instituto Mexicano del Seguro Social, Guadalajara, México | 2 Subjects | - | Case series | Craniosynostosis | laryngomalacia, acanthosis nigricans, choanal stenosis, double collecting system and dysplastic kidney | Communicating | TGS, cytogenetics | Karyotyping | AD | 4p16.3 (FGFR3) |
| Chen et al., 2001 [194] | Prenatal diagnosis and genetic analysis of type I and type II thanatophoric dysplasia | Mackay Memorial Hospital, Taipei, Taiwan | 4 Subjects, control matched sampling | Chinese | Case series | cloverleaf skull, macrocephaly, synostosis | short-limbed dwarfism, multiple skeletal dysplasias | - | TES | Direct sequencing | - | 4p16.3 (FGFR3) |
| Chen et al., 2008 [195] | Craniosynostosis and congenital tracheal anomalies in an infant with Pfeiffer syndrome carrying the W290C FGFR2 mutation | Mackay Memorial Hospital, Taipei, Taiwan | 1 Subject | - | Case study | dandy walker - cerebellar malformations | turricephalic prominent forehead, hypertelorism, low-set ears, a flat nasal bridge, mid-face hypoplasia, bilateral cleft lip and palate, a thick nuchal fold, and a distended abdomen, and multicystic kidneys | Communicating | - | - | *De novo* | 10q26.13 (c.870 G>T (TGG>TGT) in the FGFR2) |
| Chen et al., 2017 [196] | Pfeiffer syndrome with FGFR2 C342R mutation presenting extreme proptosis, craniosynostosis, hearing loss, ventriculomegaly, broad great toes and thumbs, maxillary hypoplasia, and laryngomalacia | Mackay Memorial Hospital, Taipei, Taiwan | 1 Subject | - | Case study | multisynostoses of sagittal and coronal sutures | bilateral hearing loss, brachycephaly, hypertelorism, broad big toes and thumbs, low-set ears, laryngeomalacia and midface hypoplasia | Obstructive | Cytogenetics | Karyotyping | AD | 10q26.13 (FGFR2 C342R mutation) |
| Fonseca et al., 2008 [197] | Second case of Beare-Stevenson syndrome with an FGFR2 Ser372Cys mutation | Universidade Federal do Rio de Janeiro, Rio de Janeiro, Brazil. | 1 Subject | - | Case study | craniosynostosis, crouzonoid-like features, and cloverleaf skull | cutis gyrata, acanthosis nigricans, skin furrows, skin tags, anogenital anomalies, and prominent umbilical stump | Communicating | TES | Direct sequencing | AD | 10q26.13 (FGFR2 Ser372Cys mutation.) |
| González-Del Angel et al., 2016 [198] | Expansion of the variable expression of Muenke syndrome: Hydrocephalus without craniosynostosis | Instituto Nacional de Pediatría, Mexico City, Mexico | 56 Subjects | Mexican | Case series | uni- or bicoronal craniosynostosis | wide variability | Obstructive | TES | Direct sequencing, restriction enzyme analysis | AD | 4p16.3 (FGFR3) |
| Gripp et al., 1998 [199] | Phenotype of the fibroblast growth factor receptor 2 Ser351Cys mutation: Pfeiffer syndrome type III | The Children's Hospital of Philadelphia, Pennsylvania, USA | 1 Subject | Caucasian | Case study | seizures, developmental delay, pansynostosis | bilateral elbow ankylosis, radial head dislocation, extreme proptosis with luxation of the eyes out of the lids, apnea and airway obstruction, intestinal non-rotation | Communicating | TES | SSCP, cycle sequencing | AD | 10q26.13 (Ser351Cys in FGFR2) |
| Gupta et al., 2020 [200] | Crouzon Syndrome in a Ten-week-old Infant: A Case Report | All India Institute of Medical Sciences, Patna, Bihar, India | 1 Subject | Japanese | Case study | neurologic and neuromuscular impairment, Craniosynostosis | airway obstruction, craniofacial dysostosis with abnormal shape of the skull, proptosis, hypertelorism, curved nose and frontal bossing | Communicating | - | - | AR | 10q26.13 (FGFR2) |
| Ito et al., 2018 [201] | A ZPR1 mutation is associated with a novel syndrome of growth restriction, distinct craniofacial features, alopecia, and hypoplastic kidneys | University of Ottawa, Ottawa, Canada | 4 Subjects, 3 Controls | New Mexican Hispanic heritage | Case series | microcephaly | growth restriction, distinctive craniofacial features, congenital alopecia, hypoplastic kidneys with renal insufficiency, global developmental delay, severe congenital sensorineural hearing loss, and genital hypoplasia | - | WES | Sanger sequencing | AR | 11q23.3 (ZPR1 Zinc Finger) |
| Kan et al., 2002 [202] | Genomic screening of fibroblast growth-factor receptor 2 reveals a wide spectrum of mutations in patients with syndromic craniosynostosis | The John Radcliffe Hospital, Oxford, United Kingdom | 259 Subjects, 128 Controls | - | Case series | Cloverleaf skull, craniosynostosis | - | Communicating | TES | HA | AD | 10q26.13 (FGFR2) |
| Lajeunie et al., 2006 [203] | Mutation screening in patients with syndromic craniosynostoses indicates that a limited number of recurrent FGFR2 mutations accounts for severe forms of Pfeiffer syndrome | Hôpital Necker-Enfants malades, Paris, France | 129 Subjects, 65 Controls | - | Case series | synostosis of one or several cranial sutures | ocular proptosis, maxillary hypoplasia and midface retrusion | Communicating | TES | Direct sequencing | AD | 8p11.23 (FGFR 1); 10q26.13 (FGFR2); 4p16.3 (FGFR 3 mutation) |
| Priolo et al., 2000 [204] | Pfeiffer syndrome type 2 associated with a single amino acid deletion in the FGFR2 gene | G. Gaslini Institute, Genova, Italy | 1 Subject, 60 Controls | - | Case study | acrocephalo-trigonocephaly with cloverleaf skull, callosal dysgenesis and Chiari I malformation | facial dysmorphism, radial clinodactyly of the thumbs and valgus deviation of the halluces | Unclear | TGS | Cycle sequencing | AD | 10q26.13 (FGFR2) |
| Przylepa et al., 1996 [205] | Fibroblast growth factor receptor 2 mutations in Beare-Stevenson cutis gyrata syndrome | The Johns Hopkins University School of Medicine, Baltimore, Maryland, USA | 5 Subjects, 3 Parents, and 50 Controls | - | Case series | craniosynostosis | cutis gyrata, acanthosis nigricans, craniofacial dysmorphism, digital anomalies, umbilical and anogenital abnormalities | Communicating | TGS | HA, fluorescent dideoxy terminator method, restriction enzyme analysis | AD | 10q26.13 (FGFR2) |
| Rump et al., 2006 [206] | Severe complications in a child with achondroplasia and two FGFR3 mutations on the same allele | University Medical Center Groningen, University of Groningen, The Netherlands | 1 Subject, 2 Parents | Dutch | Case study | megalecephalic | midface hypoplasia, lordotic lumbar spine, trident hand configuration, achondroplasia, respiratory failure | Communicating | TGS | Variant analysis | AD | 4p16.3 (p.G380R mutation of FGFR3) |
| Rutland et al., 1995 [207] | Identical mutations in the FGFR2 gene cause both Pfeiffer and Crouzon syndrome phenotypes | Institute of Child Health, London, UK | 12 Subjects | - | Case series | Cloverleaf skull, craniosynostosis | digital abnormalities | Communicating | TES | SSCP, direct sequencing, restriction endonuclease analysis | *De novo* | 10q26.13 (FGFR2) |
| Schaefer et al., 1998 [208] | Novel mutation in the FGFR2 gene at the same codon as the Crouzon syndrome mutations in a severe Pfeiffer syndrome type 2 case | H.A. Chapman Research Institute of Medical Genetics, Tulsa, Oklahoma, USA | 1 Subject | - | Case study | cloverleaf skull | proptosis, radioulnar synostosis and broad thumbs and great toes | Communicating | TES | Cycle sequencing | - | 10q26.13 (G to T mutation in codon 290 exon 7 of the FGFR2) |
| Takenouchi et al., 2013 [209] | Severe congenital lipodystrophy and a progeroid appearance: Mutation in the penultimate exon of FBN1 causing a recognizable phenotype | Keio University School of Medicine, Tokyo, Japan | 1 Subject | Japanese | Case study | craniosynostosis | progeroid appearance, wide-open anterior fontanelle, low-set ears, long arms and legs, arachnodactyly, and arthrogryposis, hydronephrosis | Communicating | TGS | NGS, sanger sequencing | AD | 15q21.1 (exon 64 of the FBN1 gene) |

### Extracellular matrix defects

**Table 10** highlights the genetic mutations contributing to extracellular matrix defects. Mutations were found in fukutin (*FKTN*), cartilage associated protein (*CRTAP*), collagen type VIII alpha 2 chain (*COL8A2*), collagen type III alpha 1 chain (*COL3A1*), collagen type IV alpha 1 chain (*COL4A1*), vascular cell adhesion molecule 1 (*VCAM1*), protein tyrosine phosphatase receptor type F (*PTPRF*), fibrillin 1 (*FBN1*), laminin subunit beta 1 (*LAMB1*), FRAS1 related extracellular matrix 1 (*FREM1*), and the plasminogen gene. *CRTAP* is involved in proline hydroxylation which ultimately contributes to collagen stability and functionality [76]. Mutations within the *CRTAP* gene locus can lead to Cole-Carpenter syndrome, which is associated with HC [76]. Other basement membrane proteins encoded by *COL8A2*, *COL3A1*, *COL4A1*, *VCAM1*, and *PTPRF* may alert the extracellular matrix and contribute to HC. For instance, a mutation in *COL3A1* affects its triple helix stability leading to degradation and further defects in the basement membrane [77]. *LAMB1* knockdown in zebrafish disrupted laminin integrity, a component of the basal lamina, leading to brain structural abnormalities [78], suggesting a potential pathogenic link to HC.

**Table 10:** Extracellular matrix defects.

Abbreviations include the following: Array comparative genomic hybridization (aCGH).
| Citation | Title | Author Affiliation | Case # | Ancestry | Study Design | CNS Phenotype | Non-CNS Phenotype | Type of Hydrocephalus | Genetic Methodology | Genetic Analysis | Inheritance | Genetic Findings |
| --- | --- | --- | --- | --- | --- | --- | --- | --- | --- | --- | --- | --- |
| Balasubramanian et al., 2015 [210] | CRTAP mutation in a patient with Cole-Carpenter syndrome | Sheffield Children's NHS Foundation Trust, UK | 1 Subject | Asian Pakistan | Case subject | thoraco-lumbar scoliosis and sutural craniosynostosis | Osteogenesis Imperfecta, bilateral limb deformities, joint hypermobility, prominent eyes with a proptotic appearance, greyish blue sclerae, and dentinogenesis imperfecta | Communicating | TGS | Variant analysis | - | 3p22.3 (c.118G>T mutation in exon 1 of the CRTAP gene) |
| Çiftçi et al., 2003 [211] | Ligneous conjunctivitis, hydrocephalus, hydrocele, and pulmonary involvement in a child with homozygous type I plasminogen deficiency | University of Ankara Medical School, 06100, Dikimevi Ankara, Turkey | 1 Subject, 2 Parents , 1 Control | Turkish | Case study | isolated hydrocephalus | tracheal pseudomembranes, bilateral hydrocele and unilateral inguinal hernia | Obstructive | TGS | SSCP, direct sequencing | AR | 6q26 (L650fsX652 mutation (deletion of 2081C)) |
| Cormand et al., 1999 [212] | Assignment of the muscle-eye-brain disease gene to 1p32-p34 by linkage analysis and homozygosity mapping | University of Helsinki, Finland | 12 Subjects, 27 Controls | Finnish, Turkish | Case series | intellectual disability, polymicrogyria-pachygyria-type neuronal migration disorder of the brain | ocular abnormalities, congenital muscular dystrophy | Communicating | Genotyping | Linkage analysis, haplotype analysis | AR | 1p34.3 (COL8A2), 1p21.2 (VCAM1), 1p34.2 (PTPRF) |
| Cotarelo et al., 2008 [213] | Two new patients bearing mutations in the fukutin gene confirm the relevance of this gene in Walker-Warburg syndrome | Universidad Autónoma de Madrid, Madrid, Spain | 2 Subjects, 3 Family Members | Ashkanazi Jewish, Spanish | Case series | overriding cranial bones, monolobar holoprosencephaly, interhemispheric cyst, incomplete cleavage of the thalamus and corpora quadrigemina, an absent corpus callosum and rhombencephalic hypoplasia | microphthalmia, atrial septal defect , double subaortic ventricular defect, hypoplastic left ventricle outlet, stenotic pulmonary valve and infundibular transposition of great vessels with no innominate vein, and retinal dysplasia | External and internal | TGS | Restriction endonuclease enzyme analysis, PCR | AR | 9q31.2 (FKTN) |
| de Bernabé et al., 2003 [214] | A homozygous nonsense mutation in the fukutin gene causes a Walker-Warburg syndrome phenotype | University Medical Centre Nijmegen, Nijmegen, Netherlands | 30 Subjects, 105 Controls | Japanese | Case series | cobblestone lissencephaly with agenesis of the corpus callosum, fusion of hemispheres, hydrocephalus, dilatation of the fourth ventricle, cerebellar hypoplasia, hydrocephalus, and sometimes encephalocele | eye malformations and congenital muscular dystrophy | Communicating | TGS | Linkage analysis, direct sequencing, SSCP | AR | 9q31.2 (FKTN) |
| Horn et al., 2011 [215] | Progeroid facial features and lipodystrophy associated with a novel splice site mutation in the final intron of the FBN1 gene | Charité-Universitätmedizin Berlin, Berlin, Germany | 1 Subject, 150 Controls | German | Case study | Psychomotor delay, hypotonia | triangular facial shape, large head with a broad and prominent forehead, deep set eyes with proptosis, downward slanting palpebral fissures, and a high nasal bridge, highly arched palate and mild retrognathia, generalized lipodystrophy, long fingers and toes, bilateral pes valgus | - | TGS, cytogenetics | Karyotyping, aCGH | AD | 15q21.1 (FBN1) |
| Kondo-lida et al., 1999 [216] | Novel mutations and genotype-phenotype relationships in 107 families with Fukuyama-type congenital muscular dystrophy (FCMD) | Human Genome Center, Institute of Medical Science, University of Tokyo, Japan | 19 Subjects, 50 Controls | Japanese | Case series | Intellectual delay micropolygyria, pachygyria and agyria | congenital muscular dystrophy, eye abnormalities | Communicating | TGS | SSCP, direct sequencing | *De novo* | 9q31, gene FCMD |
| Kroes et al., 2003 [217] | Ehlers-Danlos syndrome type IV: unusual congenital anomalies in a mother and son with a COL3A1 mutation and a normal collagen III protein profile | University Medical Center WKZ, Internal mail KC 04.084.2, Lundlaan 6, 3584 EA Utrecht, the Netherlands | 2 Subjects | - | Case series | Macrocephaly | blue sclerae, unilateral clubfoot, esophageal atresia, joint hyperlaxity | Communicating | TGS | - | - | 2q32.2 (COL3A1) |
| Radmanesh et al., 2013 [218] | Mutations in LAMB1 cause cobblestone brain malformation without muscular or ocular abnormalities | University of California, San Diego, CA, USA | 2 Subjects, 200 Controls | Egyptian and Turkish | Case series | cortical gyral and white-matter signal abnormalities, severe cerebellar dysplasia, brainstem hypoplasia, and occipital encephalocele | minor optic atrophy | Communicating | WES | Sanger sequencing | AR | 7q31.1 (LAMB1) |
| Saito et al., 2000 [219] | Haplotype-phenotype correlation in Fukuyama congenital muscular dystrophy | Tokyo Women's Medical University, School of Medicine, Japan | 56 Subjects, 82 Controls | Japanese | Case series | cobblestone lissencephaly with cerebral and cerebellar cortical dysplasia | congenital muscular dystrophy, eye abnormalities | Communicating | Allelotyping | Haplotype analysis, microsatellite marker assay | AR | FCMD gene |
| Schott et al., 1998 [220] | Therapy with a purified plasminogen concentrate in an infant with ligneous conjunctivitis and homozygous plasminogen deficiency | Klinikum Mannheim, University of Heidelberg, Germany | 1 Subject, 1 Control, 2 Parents, 1 Brother | Turkish | Case study | macrocephalus | pseudomembranous conjunctivitis, ligneous conjunctivitis | - | TGS | SSCP, cycle sequencing, restriction enzyme analysis | AR | 6q26 (plasminogen gene (Glu460Stop mutation)) |
| Schuster et al., 1997 [221] | Homozygous mutations in the plasminogen gene of two unrelated girls with ligneous conjunctivitis | Children's Hospital, University of Würzburg, Germany | 2 Subjects, 2 Parents, 1 Sister, 1 Control | Turkish | Case study | macrocephaly | pseudomembranous lesions of other mucous membranes in the mouth, nasopharynx, trachea, and female genital tract | Obstructive | TES | SSCP, restriction enzyme analysis | AR | 6q26 (Plasminogen gene) |
| Schuster et al., 1999 [222] | Prenatal diagnosis in a family with severe type I plasminogen deficiency, ligneous conjunctivitis and congenital hydrocephalus | Children's Hospital, University of Würzburg, Germany | 1 Subject, 2 Parents, 1 Control | Turkish | Case study | Isolated hydrocephalus | pseudomembranous conjunctivitis, ligneous conjunctivitis | Obstructive | TES | SSCP | AR | 6q26 (Plasminogen gene) |
| Tonduti et al., 2015 [223] | Cystic leukoencephalopathy with cortical dysplasia related to LAMB1 mutations | Université Paris Diderot-Sorbonne Paris Cité and INSERM U1141-DHU Protect, Paris, France | 2 Subjects, 100 Controls | - | Case series | cerebral palsy, epilepsy, spastic tetraplegia, intellectual disability | lens opacification, optic atrophy | Unclear | WES | Sanger sequencing, segregation analysis | - | 7q31.1 (LAMB1) |
| Van der Knaap et al., 2006 [224] | Neonatal porencephaly and adult stroke related to mutations in collagen IV A1 | VU University Medical Center, Amsterdam, the Netherlands | 3 Subjects, 192 Controls | Dutch | Case series | Leukoencephalopathy, porencephalic cysts, cerebral microangiopathies | cataracts, blood vessel defects | Obstructive (blood, calcifications) vs. Porencephaly | - | - | AD | 13q34 (mutation in the COL4A1) |
| Yang et al., 2017 [225] | Novel FREM1 mutations are associated with severe hydrocephalus and shortened limbs in a prenatal case | The First Affiliated Hospital of Jinan University, Guangzhou, Guangdong, China | 1 Subject, 200 Controls | Chinese | Case study | isolated hydrocephalus | short limbs | - | WES | Sanger sequencing | - | 9p22.3 (FREM1) |

### Neurogenesis and neural stem cell biology

**Table 11** summaries gene mutations implicating neurogenesis. Mutations were identified in SRY-box transcription factor 9 (*SOX9*), solute carrier family 29 member 3 (*SLC29A3*), adhesion G protein-coupled receptor (*ADGRG1*), katanin interacting protein (*KIAA0556*), G protein signaling modulator 2 (*GPSM2*), tripartite motif containing 71 (*TRIM71*), SWI/SNF related, matrix associated, actin dependent regulator of chromatin subfamily c member 1 (*SMARCC1*), patched 1 (*PTCH1*), FLVCR heme transporter 2 (*FLVCR2*), intestinal cell kinase (*ICK*), cystathionine beta-synthase (*CBS*), 5-methyltetrahydrofolate-homocysteine methyltransferase reductase (*MTRR*), interleukin 4 induced 1 (*IL4I1*), scribble planar cell polarity protein (*SCRIB1*), protein tyrosine kinase 7 (*PTK7*), frizzled class receptor 1 (*FZD1*), VANGL planar cell polarity protein 2 (*VANGL2*), dishevelled segment polarity protein (*DVL2*), transcription elongation factor B polypeptide 3B (*TCEB3B*), phospholipase C delta 4 (*PLCD4*), Ras associated domain family member 4 (*RASSF4*), phenylalanyl-tRNA synthetase 2, mitochondrial (*FARS2*), tubulin beta 3 class III (*TUBB3*), and discs large MAGUK scaffold protein 5 (*DLG5*). Frameshift mutations were seen in WD repeat domain 81 (*WDR81*), kinase D interacting substrate 220 (*KIDINS220*). Deletions were seen in chromosome 6 (6q25.3 and 6p25), chromosome 13 (13q), chromosome 16 (16p12.2), and chromosome 22 (22q11.2). The 6p25 deletion resulted in the deletion of forkhead box C1 (*FOXC1*), forkhead box F2 (*FOXF2*), and forkhead box Q1 (*FOXQ1*).

**Table 11:** Neurogenesis.

Abbreviations include the following: Array comparative genomic hybridization (aCGH).
| Citation | Title | Author Affiliation | Case # | Ancestry | Study Design | CNS Phenotype | Non-CNS Phenotype | Type of Hydrocephalus | Genetic Methodology | Genetic Analysis | Inheritance | Genetic Findings |
| --- | --- | --- | --- | --- | --- | --- | --- | --- | --- | --- | --- | --- |
| Antwi et al., 2018 [226] | A novel association of campomelic dysplasia and hydrocephalus with an unbalanced chromosomal translocation upstream of SOX9 | Yale University, New Haven, CT, United States | 1 Subject | - | Case study | hypoplastic C6 vertebral body, exaggerated cervical lordosis, and exaggerated thoracic kyphosis | tracheobronchom alacia, cleft palate, retrognathia, hypertelorism, hypoplastic mandible | Communicating | WES, cytogenetics | Karyotyping, FISH, aCGH | *De novo* | 17q24.3 (SOX9) |
| Avitan-Hersh et al., 2011 [227] | A case of H syndrome showing immunophenotype similarities to Rosai-Dorfman disease | Technion Institute of Technology, Haifa, Israel | 1 Subject, 2 Parents | Arab | Case study | isolated hydrocephalus | pulmonic stenosis, skin hyperpigmentation, hepatomegaly, splenomegaly, dilatation of the right renal pelvis | Communicating | TES | - | AR | 10q22.1 (SLC29A3 gene, encodes human equilibrative nucleoside transporter hENT3) |
| Cauley et al., 2019 [228] | Overlap of polymicrogyria, hydrocephalus, and Joubert syndrome in a family with novel truncating mutations in ADGRG1/GPR56 and KIAA0556 | The George Washington University School of Medicine and Health Sciences, Washington, DC, USA | 2 Subjects, 2 Siblings, 2 Parents, Controls used | Sudanese | Case series | psychomotor delay, intellectual disability, seizures, severe brain malformations, spasticity, hyperreflexia | ptosis, unilateral ophthalmoplegia, and bilateral vertical ophthalmoplegia, muscle wasting | - | WES | Variant analysis, Sanger sequencing | AR | 16q21 (ADGRG1) And 16p12.1 (KIAA0556) |
| Christofolini et al., 2006 [229] | Hydrocephaly, penoscrotal transposition, and digital anomalies associated with de novo pseudodicentric rearranged chromosome 13 characterized by classical cytogenetic methods and mBAND analysis | Departamento de Morfologia, Disciplina de Genética, Universidade Federal de São Paulo, São Paulo, Brazil | 1 Subject, 2 Parents | - | Case study | corpus callosum agenesis | imperforate anus with anocutaneous fistula, penoscrotal transposition, and digital reduction defects, short palpebral fissures, telecanthus, epicanthic folds, short nose with depressed nasal bridge and anteverted nostrils, posteriorly rotated ears, short neck | Obstructive | Cytogenetics | G-banding | *De novo* | 13q deletion |
| Doherty et al., 2012 [230] | GPSM2 mutations cause the brain malformations and hearing loss in Chudley-McCullough syndrome | University of Washington, Seattle Children's Hospital, USA | 12 Subjects, Controls used | Mennonite, European American, Dutch | Case series | bilateral sensorineural deafness, corpus callosum agenesis, arachnoid cysts, posterior agenesis of the corpus callosum, frontal polymicrogyria, frontal heterotopia, cerebellar dysplasia | down slanting palpebral fissures and low-set, posteriorly rotated ears | Communicating, Obstructive | Genotyping, WES | SNP, sanger sequencing | AR | 1p13.3 (G protein-signaling modulator 2 gene, GPSM2) |
| Forrester et al., 2002 [231] | Kousseff syndrome caused by deletion of chromosome 22q11-13 | Southern Illinois University School of Medicine, Springfield, Illinois, USA | 3 Subjects, 2 Controls | - | Case series | intellectual disability | lumbosacral myelomeningocele, cleft palate, and dysmorphic features consisting of low-set and posteriorly rotated ears, retrognathia, and clinodactyly of the fifth toes, cardiac anomalies | Obstructive | Genotyping, cytogenetics | FISH, karyotyping, microsatellite marker assay | AR | 22q11.2-microdeletion |
| Furey et al., 2018 [232] | De Novo Mutation in Genes Regulating Neural Stem Cell Fate in Human Congenital Hydrocephalus | Yale University School of Medicine, New Haven, CT 06510, USA | 177 subjects, 1,789 controls | - | Case series | isolated hydrocephalus | - | Communicating, Obstructive | WES | Direct sequencing | *De novo* | 3p22.3 (TRIM71)<br>3p21.31 (SMARCC1)<br>9q22.32 (PTCH1) |
| Grosso et al., 2002 [233] | De novo complete trisomy 5p: clinical and neuroradiological findings | University of Siena, Siena, Italy | 1 Subject | - | Case study | isolated hydrocephalus | Low-set, posteriorly rotated ears with reduced cartilage, up slanted palpebral fissures, epicanthus, hypertelorism, a wide and depressed nasal root, a short nose with anteverted nostrils, a long philtrum, retrognathia, an ogival palate, a short neck, abnormal palmar creases, and a bell-shaped trunk | - | Cytogenetics | FISH w/ WCP | *De novo* | trisomy 5p |
| Jacquemin et al., 2020 [234] | TrkA mediates effect of novel KIDINS220 mutation in human brain ventriculomegaly | Université Libre de Bruxelles, 1070 Brussels, Belgium | 3 Subjects, 1 Control | Pakistan | Case series | isolated hydrocephalus | limb contractures, club feet | - | WES | Variant analysis | AR | 2p25.1 (homozygous variant of KIDINS220) |
| Kline-Fath et al., 2018 [235] | Fowler syndrome and fetal MRI findings: a genetic disorder mimicking hydranencephaly/hydrocephalus | Cincinnati Children's Hospital Medical Center, 3333 Burnet Ave., Cincinnati, OH, USA | 1 Subject | - | Case study | thin cerebral cortex, cerebellum, brainstem, and spinal cord | Arthrogryposis, proliferative glomeruloid vasculopathy | Obstructive | WES | - | AR | 14q24.3 (FLVCR2 gene) |
| Koenigstein et al., 2016 [236] | Chudley-McCullough Syndrome: Variable Clinical Picture in Twins with a Novel GPSM2 Mutation | Justus-Liebig-University, Giessen, Germany | 2 Subjects | Turkish | Case series | callosal agenesis, interhemispheric cyst, frontal polymicrogyria | sensorineural deafness | Communicating | - | - | AR | 1p13.3 (c.C1093T; p.R365X in GPSM2) |
| Lahiry et al., 2009 [237] | A multiplex human syndrome implicates a key role for intestinal cell kinase in development of central nervous, skeletal, and endocrine systems | Robarts Research Institute, London, Ontario N6A 5K8, Canada | 6 Subjects , 3112 Controls | Amish | Case series | Cerebral anomalies | facial dysmorphisms, eye anomalies, skeletal anomalies, pulmonary/GI/GU dysplasia | Communicating | Genotyping, TGS | SNP, autozygosity mapping, direct sequencing | AR | 6p12.1 (ICK p.R272Q mutation) |
| Li et al., 2015 [238] | Congenital hydrocephalus and hemivertebrae associated with de novo partial monosomy 6q (6q25.3→qter) | The Third Affiliated Hospital of Guangzhou Medical University, Guangzhou, Guangdong, People's Republic of China | 1 Subject | - | Case study | Isolated Hydrocephalus | Lumbar hemivertebrae | - | Cytogenetics | CNV, aCGH, Karyotyping, FISH | - | deletion in chromosome region 6q25.3→qter |
| Maclean et al., 2004 (258) | Kousseff syndrome: a causally heterogeneous disorder | Sydney Children's Hospital, Sydney, Australia | 2 Subjects | Indonesian | Case series | myelomeningocele, callosal hypoplasia, intellectual delay | posteriorly rotated ears, a large nose, a smooth featureless philtrum, hypertrichosis and restricted ankle dorsiflexion, tetralogy of fallot | Obstructive | TES | Cycle sequencing | AR | 22q11.2-microdeletion |
| Maclean et al., 2005 [239] | Axenfeld-Rieger malformation and distinctive facial features: Clues to a recognizable 6p25 microdeletion syndrome | The Children's Hospital at Westmead, Sydney, New South Wales, Australia | 1 Subject | Caucasian | Case study | cerebellar hypoplasia, a deficient inferior vermis, hypoplasia of the pons, medulla, and posterior corpus callosum, and absent septum pellucidum | Axenfeld-Rieger malformation, hearing loss, congenital heart disease, dental anomalies, developmental delay, and a characteristic facial appearance | Communicating | Cytogenetics, genotyping | Karyotyping, FISH, microsatellite marker assay, | *De novo* | 6p25 (deletion of the FOXC1/FOXF2/FOXQ1 forkhead gene cluster) |
| Mero et al., 2017 [240] | Homozygous KIDINS220 loss-of-function variants in fetuses with cerebral ventriculomegaly and limb contractures | Oslo University Hospital, Oslo, Norway | 4 Subjects , 2 Parents | - | Case series | callosum agenesis, small cerebellum | limb contractures | Communicating | WES | Sanger sequencing, autozygosity mapping, | AR | 2p25.1 (homozygous frameshift variant in exon 24 in KIDINS220) |
| Pappa et al., 2017 [241] | Exome analysis in an Estonian multiplex family with neural tube defects-a case report | University of Tartu, Riia 23b, 51010, Tartu, Estonia | 3 Subjects , 2 Parents | Estonian | Case series | spina bifida, aqueductal stenosis, intellectual delay | gait and motor abnormalities | Obstructive | WES | Variant analysis | Maternal | 21q22.3 (CBS), 5p15.31 (MTRR), 1p36.22 (MTHFR), 19q13.33 (IL4I1), 8q24.3 (SCRIB1), |
|  |  |  |  |  |  |  |  |  |  |  |  | 6p21.1 (PTK7), 7q21.13 (FZD1), 1q23.2 (VANG12), 17p13.1 (DVL2), 18q21.1 (TCEB3B), 2q35 (PLCD4), 10q11.21 (RASSF4), and 6p25.1 (FARS2) |
| Powis et al., 2018 [242] | Postmortem Diagnostic Exome Sequencing Identifies a De Novo TUBB3 Alteration in a Newborn with Prenatally Diagnosed Hydrocephalus and Suspected Walker-Warburg Syndrome | Ambry Genetics, Aliso Viejo, California, USA | 1 Subject, 2 Parents | Caucasian | Case study | posterior fossa cyst, dandy walker malformation, seizures | optic nerve abnormalities, abnormal renal function | - | Diagnostic exome sequencing | - | *De novo* | 16q24.3 (TUBB3) |
| Rai et al., 2015 [243] | Cervicomedullary spinal stenosis and ventriculomegaly in a child with developmental delay due to chromosome 16p12.1 microdeletion syndrome | Midland Regional Hospital, Mullingar Westmeath, Ireland | 1 Subject | - | Case study | macrocephaly | significant delay in gross motor skills | - | Cytogenetics | aCGH | - | Chr. 16p12.2 deletion |
| Su et al., 2021 [244] | Novel compound heterozygous frameshift variants in WDR81 associated with congenital hydrocephalus 3 with brain anomalies: First Chinese prenatal case confirms WDR81 involvement | Guangxi Health Commission Key Laboratory of Precise Diagnosis and Treatment of Genetic Diseases, Maternal and Child Health Hospital of Guangxi Zhuang Autonomous Region, Nanning, China | 2 Subjects | Chinese | Case series | cerebellar hypoplasia | cleft lip and palate, hydrops fetalis, hepatomegaly | HYC3 | WES | Sanger sequencing, variant analysis | AR | 17p13.3 (WDR81) |
| Yüksel et al., 2019 [245] | A homozygous frameshift variant in an alternatively spliced exon of DLG5 causes hydrocephalus and renal dysplasia | Centogene AG, Rostock, Germany | 1 Subject, 1 Control | - | Case study | isolated hydrocephalus | atrial and ventricular septal defects, cleft lip and palate, and a renal phenotype including multi-cystic dysplasia | Obstructive | WES | Variant analysis, sanger sequencing | *De novo* | 10q22.3 (DLG5) |

The heterogeneity of neurogenesis-associated HC suggests that numerous genes involved in development may confer susceptibility to this phenotype. *SOX9* knockdown in mice suggest a role in neural stem cell development and ependymal cell maintenance as a pathogenic mechanism that causes HC [79]. In addition, mutations in *ADGRG1* have been shown to impact cerebral cortex development and neuronal migration via the perturbation of the RhoA pathway [80]. In addition, *GPSM2* has been shown to alter neuroepithelial function through disruption of cellular orientation and planarity leading to aberrant brain development [81]. Finally, mice lacking *KIDINS220* display attenuated responses to neurotrophic factors and have impaired development in multiple signaling pathways [82]. Understanding the genetic influence of neurogenesis may elucidate a better understanding of patient characteristics and poor outcomes in the HC phenotype.

### Inherited cancer syndromes

**Table 12** summarizes genes that contributed to tumor pathogenesis, and which result in the development of HC. Mutations are seen in NRAS proto-oncogene, GTPase (*NRAS*), von Hippel-Lindau tumor suppressor (*VHL*), patched 1 (*PTCH1*) and FA complementation group C (*FANCC*). Germline mutations are seen in phosphatase and tensin homolog (*PTEN*) and SUFU negative regulator of hedgehog signaling (*SUFU*). Deletions within chromosome 11 (11p13) and chromosome 9 (9q22.3 and 9q22-q31) were also identified. *NRAS* is an oncogene contributing to the development of congenital melanocytic nevi, a condition associated with HC [83]. Clinically relevant mutations in Von Hippau Lindau (*VHL*) affect protein expression and degradation where patients with or without a mass lesion (i.e., hemangioblastoma) develop HC [84]. Gorlin syndrome is disorder characterized with bony abnormalities and an increased risk for multiple CNS and non-CNS tumors. Previous studies have mapped this syndrome to deletions in the 9q22 locus which is consistent with the patients identified in this review with mutations specifically affecting *PTCH1* and *FANCC* genes [85; 86]. Finally, mutations in SUFU have also been associated with Gorlin syndrome [87; 88].

**Table 12:** Inherited cancer syndromes.

Abbreviations include the following: Autosomal Dominant (AD). Fluorescence In Situ Hybridization (FISH). Multiplex ligation dependent probe amplification (MLPA). Single nucleotide polymorphisms (SNP). Targeted genome sequencing (TGS).
| Citation | Title | Author Affiliation | Case # | Ancestry | Study Design | CNS Phenotype | Non-CNS Phenotype | Type of Hydrocephalus | Genetic Methodology | Genetic Analysis | Inheritance | Genetic Findings |
| --- | --- | --- | --- | --- | --- | --- | --- | --- | --- | --- | --- | --- |
| Demir et al., 2011 [246] | WAGR syndrome with tetralogy of Fallot and hydrocephalus | Hacettepe University, Ankara, Turkey | 1 Subject | - | Case study | isolated hydrocephalus | Wilms tumor, aniridia, genitourinary abnormalities, and intellectual disability | Communicating | Cytogenetics | G-banding | *De novo* | deletion of chromosome 11p13 |
| Fukino et al., 2000 [247] | A family with hydrocephalus as a complication of cerebellar hemangioblastoma: identification of Pro157Leu mutation in the VHL gene | Nippon Medical School, Kawasaki-shi, Japan | 2 Subjects | Japanese | Case series | isolated hydrocephalus | retinal angioma, cerebellar, hemangioblastomas, pancreatic cysts | Obstructive | TES | Direct sequencing, restriction enzyme analysis | - | 3p25.3 (VHL) |
| Kinsler et al., 2013 [248] | Multiple congenital melanocytic nevi and neurocutaneous melanosis are caused by postzygotic mutations in codon 61 of NRAS | Great Ormond Street Hospital for Children, London, UK | 5 Subjects, Controls used | - | Case series | arachnoid cysts, syringomyelia, tumors (including astrocytoma, choroid plexus papilloma, ependymoma, and pineal germinoma), Dandy–Walker, and Chiari malformation | widespread melanocytic nevi | Communicating, Obstructing | TGS, cytogenetics | aCGH, direct sequencing | Non-mendelian inheritance | 1p13.2 (c.181C>A, p.Q61K NRAS mutations) |
| Kusakabe et al., 2018 [249] | Combined morphological, immunohistochemical and genetic analyses of medullopithelioma in the posterior cranial fossa | Ehime University School of Medicine, Toon, Japan | 1 Subject | - | Case study | Medullopithelioma | - | Obstructive | Cytogenetics | FISH | - | No C19MC mutations |
| Pastorino et al., 2009 [250] | Identification of a SUFU germline mutation in a family with Gorlin syndrome | Università degli Studi di Genova, Genova, Italy | 1 Subject, 1 Control | Caucasian | Case study | spina bifida | pits in hands and soles, coarse facies, strabismus, cleft lip and palate, bifid ribs | Obstructive | TGS | MPLA, direct sequencing | AD | 10q24.32 (c.1022 + 1G>A SUFU germ line splicing mutation) |
| Reardon et al., 2001 [251] | A novel germline mutation of the PTEN gene in a patient with macrocephaly, ventricular dilatation, and features of VATER association | Our Lady's Hospital for Sick Children, Crumlin, Dublin 12, Ireland | 1 Subject, 2 Parents | - | Case study | macrocephaly | hypoplasia of the thumbs bilaterally with radial deviation of the hands, 13 pairs of ribs | Communicating | Cytogenetics, TGS | Karyotyping | AD | 10q23.31 PTEN |
| Reichert et al., 2015 [252] | Diagnosis of 9q22.3 microdeletion syndrome in utero following identification of craniosynostosis, overgrowth, and skeletal anomalies | Children's Hospital of Philadelphia, Philadelphia, Pennsylvania | 1 Subject, 2 Parents | - | Case study | metopic craniosynostosis, intellectual disability, trigonocephaly | macrosomia, hepatomegaly, nephromegaly, and anomalous vertebrae | Communicating | Cytogenetics | Karyotyping, SNP, FISH | *De novo* | 9q22.32 (PTCH1) and 9q22.32 (FANCC) genes 9q22.3 |
| Shimkets et al., 1996 [253] | Molecular analysis of chromosome 9q deletions in two Gorlin syndrome patients | Yale University School of Medicine, | 2 Subjects, 4 Parents | African American | Case series | macrocephalus, agenesis of the corpus callosum | bilateral inguinal hernias, bilateral | Communicating | Cytogenetics, genotyping, | G-banding, restriction | AD | chromosome 9q22 deletion and 9q22-q3l |
|  |  | New Haven, CT 06520-8005, USA |  | Caucasian |  |  | conductive hearing loss, strabismus, and ectopic eruption of the upper central incisors, multiple basal cell carcinomas, medulloblastomas, ovarian fibromas |  |  | enzyme analysis |  |  |
| Uguen et al., 2015 [254] | Severe hydrocephalus caused by diffuse leptomeningeal and neurocutaneous melanocytosis of antenatal onset: a clinical, pathologic, and molecular study of 2 cases | Service d'anatomie et cytologie pathologiques, Brest, F-29220 France; Université Européenne de Bretagne, 29238 France | 2 Subjects | - | Case series | leptomeningeal pigmentation, Dandy walker malformation | melanocytic nevi | Obstructive | Cytogenetics, TGS | aCGH, FISH, pyrosequencing, NGS | - | 1p13.2 (NRAS) |

### WNT signaling

WNT signal transduction is involved in numerous pathways regulating cell function and development. **Table 13** summarizes gene mutations identified in HC patients with this pathway. Numerous studies have reported gene mutations in coiled-coil and C2 domain containing 2A (*CC2D2A*) and coiled-coil domain containing 88C (*CCDC88C*). COACH syndrome is defined as cerebellar vermis hypoplasia, oligophrenia, ataxia, colobomas, and hepatic fibrosis [89]. This gene locus has been shown to interact with the WNT signaling pathway and is associated with centrosome stability [90]. In addition, *CCDC88C* is associated with the WNT signaling pathway through interaction with the Dishevelled protein [91]. The dishevelled protein contains a binding domain which interacts with a hook related protein transcribed from the *CCDC88C* locus [92]. WNT signaling plays numerous roles in cell communication and embryonic development, suggesting potential mechanisms contributing to HC [93].

**Table 13:**
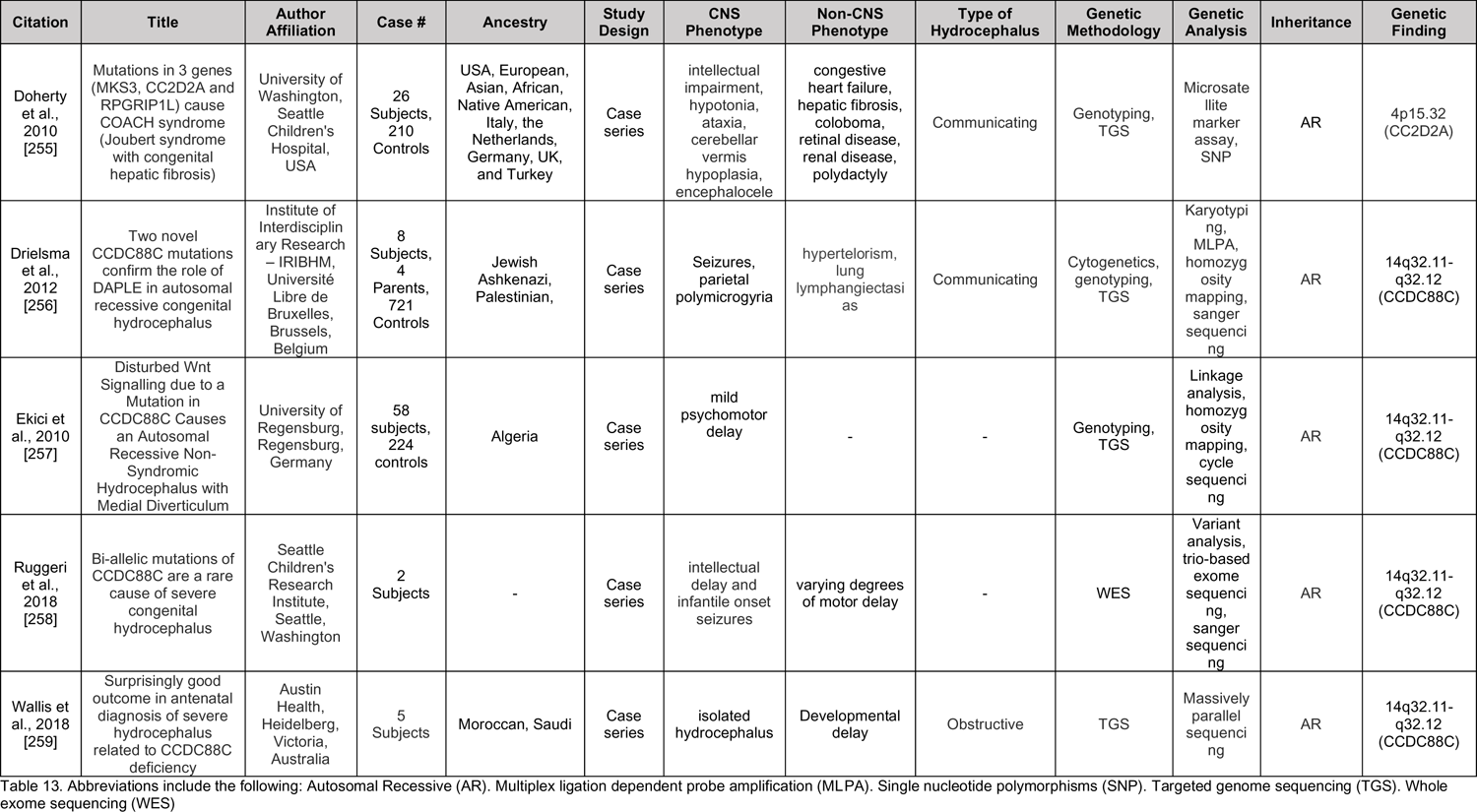
WNT signaling.

### Transcriptional, Post-Transcriptional, and Epigenetic Regulation

**Table 14** summarizes mutations in genes that regulate transcription, post-transcriptional, and epigenetic processes. Missense mutations were seen in THO complex subunit 6 (*THOC6*) and *HYLS1*, genes involved in transcriptional regulation. Patients with loss of function mutations in FA complementation group L (*FANCL*) we identified. Additional mutations observed included interferon regulatory factory 6 (*IRF6*), small nucleolar RNA, C/D box 118 (*SNORD118*), nuclear factor I A (*NFIA*), SET binding protein 1 (*SETBP1*), SWI/SNF related, matrix associated, actin dependent regulator of chromatin, subfamily b, member 1 (*SMARCB1*), maelstrom spermatogenic transposon (*MAEL*), a deletion in chromosome 5 (5q35.3), and 20q13.3 trisomy. Deletions in chromosome 1 (1q42.3-q44) resulted in the deletion of zinc finger and BTB domain containing 18 (*ZBTB18*) and heterogeneous nuclear ribonucleoprotein U (*HNRNPU*). THOC6 is a part the TREX complex responsible for mRNA export and is localized to the 5’ cap of mRNA [94]. It has been associated with Beaulieu-Boycott-Innes syndrome, which is charactered by developmental delay and organ dysgenesis [95]. Patients identified in this review with Beaulieu-Boycott-Innes syndrome and *THOC6* mutations have been shown to develop HC, suggesting a role for mRNA export regulation in association with HC phenotypes [96]. *HYLS1* is associated with Hydrolethalus syndrome, a disorder characterized by HC and craniofacial abnormalities [97]. Expression analysis of this gene suggests a role in CNS development, where a HC associated mutation gene causes nuclear localization whereas the WT form is expressed in the cytoplasm [97]. *SNORD118* is involved in regulation of ribosome biology and associated with the hydrocephalic phenotype of Labrune syndrome, characterized by leukoencephalopathy, intracranial cysts, and calcification [98]. While the function of *SETBP1* remains largely unknown, mutations in this gene are associated with Schinzel-Giedion syndrome, characterized by facial abnormalities, intellectual disability, congenital malformations, and HC [99]. *SMARCB1* is involved in chromatin remodeling to further enhance or repress transcription [100]. Finally, a transcriptome-wide association study (TWAS) and multi-omics study of HC identified maelstrom (*MAEL*), a gene that regulates transposons and epigenetic modifications, as an experiment-wide predictor of HC in the cortex [10; 101]. These studies identified transcriptional regulators and further emphasize the need to explore these mechanisms to understand the mechanistic associations with HC.

**Table 14:** Transcriptional, Post-Transcriptional, and Epigenetic Regulation.

Abbreviations include the following: Array comparative genomic hybridization (aCGH).
| Citation | Title | Author Affiliation | Case # | Ancestry | Study Design | CNS Phenotype | Non-CNS Phenotype | Type of Hydrocephalus | Genetic Methodology | Genetic Analysis | Inheritance | Genetic Finding |
| --- | --- | --- | --- | --- | --- | --- | --- | --- | --- | --- | --- | --- |
| Chen et al., 2020 [260] | Prenatal diagnosis and molecular cytogenetic characterization of a chromosome 1q42.3-q44 deletion in a fetus associated with ventriculomegaly on prenatal ultrasound | Mackay Memorial Hospital, Taipei, Taiwan | 1 Subject, 2 Parents | - | Case study | anomalies of corpus callosum, microcephaly | hypertelorism, large low-set ears, micrognathia, a broad nose, arched eyebrows, prominent forehead and flat nasal bridge | - | Cytogenetics | aCGH, FISH, polymorphic DNA marker analysis | Paternal | 1q42.3-q44 deletion (including 1q43 (RGS7), 1q43 (FH), 1q43 (CEP170), 1q43-44 (AKT3), 1q44 (ZBTB18 and 1q44 (HNRNP U)) |
| Diets et al., 2019 [261] | A recurrent de novo missense pathogenic variant in SMARCB1 causes severe intellectual disability and choroid plexus hyperplasia with resultant hydrocephalus | Radboud University Medical Center and Radboud Institute for Molecular Life Sciences, Nijmegen, The Netherlands | 4 Subjects | - | Case series | Choroid plexus hyperplasia w/ papilloma, truncal hypotonia, intellectual disability | visual impairment, myopia, sleep apnea, joint hypermobility, renal and cardiac anomalies | - | WES | Trio-based exome sequencing | De novo | 22q11.23 (SMARCB1) |
| Hale et al., 2021 [262] | Multi-omic analysis elucidates the genetic basis of hydrocephalus | Vanderbilt University School of Medicine, Medical Scientist Training Program, Nashville, TN | 287 Subjects , 18,740 Controls | European | Case series | various neurological phenotypes | - | Variable | Gene expression | PrediXcan analysis | Variable | 1q24.1 (MAEL) |
| Hishimura et al., 2016 [263] | Genetic and prenatal findings in two Japanese patients with Schinzel-Giedion syndrome | Tenshi Hospital N-12, E-3 Sapporo, Japan | 2 Subjects | Japanese | Case series | Isolated Hydrocephalus | overlapping fingers, hydronephrosis. high, prominent forehead, hypertelorism, and depressed nasal root | - | TES, cytogenetics | G-banding | AD | 18q12.3 (SETBP1) |
| Mattioli et al., 2019 [264] | Clinical and functional characterization of recurrent missense variants implicated in THOC6-related intellectual disability | Institut de Génétique et de Biologie Moléculaire et Cellulaire, 67400 Illkirch-Graffenstaden, France | 2 Subjects , controls used | European | Case series | Intellectual disability, multiple brain abnormalities | facial dysmorphism, a cleft palate, micrognathia, choanal atresia, congenital heart defect, micropenis | Communicating | TGS, cytogenetics | Karyotyping, aCGH, SNP | AR | 16p13.3 (THOC6 gene-Trp100Arg, Val234Leu, Gly275Asp) |
| Mee et al., 2005 [265] | Hydroletharus syndrome is caused by a missense mutation in a novel gene HYLS1 | David Geffen School of Medicine, University of California at | 24 subjects , 40 Controls | Finland | Case series | absent midline structures of the brain | micrognathia, polydactyly, defective lobation of the lungs, anomalies of the respiratory | Communicating | Genotyping, TGS | Microsatellite marker analysis, SNP, haplotype | AR | 11q24.2 (HYLS1 gene) |
|  |  | Los Angeles, Los Angeles, CA, USA |  |  |  |  | tract, small chin and anomalous nose |  |  | analysis, two-point linkage analysis |  |  |
| Negishi et al., 2015 [266] | Truncating mutation in NFIA causes brain malformation and urinary tract defects | Nagoya City University Graduate School of Medical Sciences, Nagoya, Japan | 1 Subject, Control database used | - | Case study | ventricular enlargement, callosal agenesis, urinary tract defects, mildly dysmorphic facial features | urinary tract defects | Communicating | WES | Variant analysis, sanger sequencing | *De novo* | 1p31.3 (de novo truncating mutation (c.1094delC; p.Pro365Hisfs*32) in the NFIA gene) |
| Nyboe et al., 2015 [267] | Familial craniosynostosis associated with a microdeletion involving the NFIA gene | Copenhagen University Hospital Rigshospitalet, Copenhagen, Denmark | 4 Subjects | - | Case series | hypoplasia of the corpus callosum, craniosynostosis, lambdoid synostosis | dysmorphic features, renal defects | Obstructive | Cytogenetics | aCGH | *De novo* | 1p31.3 (NFIA gene) |
| Shtaya et al., 2019 [268] | Leukoencephalopathy, Intracranial Calcifications, Cysts, and SNORD118 Mutation (Labrune Syndrome) with Obstructive Hydrocephalus | Neurosciences Research Centre, St. George's, University of London, London, United Kingdom; Atkinson Morley Neurosurgery Centre, St. George's University Hospital NHS Foundation Trust, London, United Kingdom | 1 Subject | - | Case study | widespread intracranial calcifications, cysts, and leukoencephalopathy | motor developmental delay | Obstructive | - | - | - | 17p13.1 (SNORD 118) |
| Verkerk et al., 2010 [269] | Unbalanced der(5)t(5;20) translocation associated with megalencephaly, perisylvian polymicrogyria, polydactyly and hydrocephalus | Erasmus Medical Center, Rotterdam, The Netherlands | 2 Subjects | Dutch | Case series | perisylvian polymicrogyria, megalencephaly | ASD, hypothalamic hypothyroidism, kyphoscoliosis, pectus carinatum and rickets, vesicoureteral reflux, high broad forehead, large fontanel, hypertelorism with epicanthic folds, short, upturned nose with hypoplastic nostrils, down turned corners of the mouth with thick vermilion of the lips, high arched palate, small, pointed chin with a vertical groove, large low-set ears, barrel shaped chest with kyphoscoliosis, postaxial polydactyly of the 5th right toe | - | Cytogenetics, WGS | Karyotyping, MLPA, FISH, SNP, CNV | - | 5q35.3 deletion and 20q13.3 trisomy |
| Vetro et al., 2015 [270] | Loss-of-Function FANCL Mutations Associate with Severe Fanconi Anemia Overlapping the VACTERL Association | Biotechnology Research Laboratories, Fondazione IRCCS Policlinico San Matteo, Pavia, Italy | 3 Subjects, 2 Parents, 3 Controls | Morocco, Dutch | Case series | aqueductal stenosis, cerebellar hypoplasia | bilateral radial and thumbs aplasia, hypoplasia of the left shoulder girdle, bilateral club feet, micrognathia, single and ectopic kidney, absent uterus, micropenis, hypoplastic lungs with abnormal lobation, Tetralogy of Fallot, ventricular septal defect and patent ductus arteriosus, esophageal atresia with tracheoesophageal fistula, anal atresia and rectovaginal fistula | - | WES | Sanger sequencing | AR | 2p16.1 (FANCL truncating mutation) |
| Zechi-Ceide et al., 2007 [271] | Hydrocephalus and moderate mental retardation in a boy with Van der Woude phenotype and IRF6 gene mutation | Hospital de Reabilitação de Anomalias Craniofaciais Department of Biological Sciences, Universidade Estadual Paulista, Bauru Human Genome Center and Department of Genetics and Evolutionary Biology, Institute of Biosciences, USP, São Paulo, SP, Brazil | 1 Subject, 2 Parents, Controls used | Finnish | Case study | callosal hypoplasia, intellectual delay | lip pits, distinct craniofacial dysmorphism with cleft lip and palate | - | TES | Segregation analysis, direct sequencing | AD w/ variable expressivity | 1q32.2 (IRF6) |

### Ion Transport and Regulation

**Table 15** summarizes gene mutations implicating ion transport. Mutations were seen in aquaporin 4 (*AQP4*) and FLVCR heme transporter 2 (*FLVCR2*). Mutations on chromosome 17 (17p13) implicated transient receptor potential cation channel subfamily V member 3 (*TRPV3*). Aquaporin 4 (*AQP4*) regulates water transport on ependymal cells and knockout of this gene in mice show disrupted gap junctions which alter the ependymal zone and CSF flow contributing to HC development [102; 103]. Mutations in the enhancer of *TMEM50b* alter expression of *TTF*, a direct transcriptional regulator of *AQP1*, have also been identified [101]. Mutations in *FLVCR2* are associated with Fowler’s syndrome, a disorder characterized by HC and hydranencephaly [104]. This gene locus encodes a transmembrane protein involved in solute transport, suggesting that defects in chemiosmotic regulation contribute to HC development [105].

**Table 15:** Ion Transport and Regulation.

Abbreviations include the following: Autosomal Recessive (AR). Single nucleotide polymorphisms (SNP). Targeted genome sequencing (TGS). Whole exome sequencing (WES).
| Citation | Title | Author Affiliation | Case # | Ancestry | Study Design | CNS Phenotype | Non-CNS Phenotype | Type of Hydrocephalus | Genetic Methodology | Genetic Analysis | Inheritance | Genetic Finding |
| --- | --- | --- | --- | --- | --- | --- | --- | --- | --- | --- | --- | --- |
| Castañeyra-Ruiz1 et al., 2013 [272] | Aquaporin-4 expression in the cerebrospinal fluid in congenital human hydrocephalus | Facultad de Medicina, Universidad de La Laguna, La Laguna, Tenerife, Canary Island, Spain | 13 Subjects, 4 Controls | - | Case series | isolated hydrocephalus | - | Communicating, Obstructive | Gene expression | Western blot, ELISA assay | - | 18q11.2 (AQP4) |
| Kvarnung et al., 2016 [273] | Mutations in FLVCR2 associated with Fowler syndrome and survival beyond infancy | Center for Molecular Medicine, Karolinska Institutet, Stockholm, Sweden | 2 Subjects | Somalian | Case series | Intellectual disability, glomerular vasculopathy in the central nervous system, hypokinesia/akinesia | arthrogryphosis | - | TES | Variant analysis, sanger sequencing | AR | 14q24.3 (FLVCR2) |
| Lalonde et al., 2010 [274] | Unexpected allelic heterogeneity and spectrum of mutations in Fowler syndrome revealed by next-generation exome sequencing | McGill University and Genome Quebec Innovation Centre, Montreal, Canada | 2 Subjects | French Canadian | Case series | CNS microcalcifications and hyperplastic microvessels forming glomeruloid structures | arthrogryposis multiplex, webbing of joints, muscular atrophy | Obstructive | WES | Variant analysis, SNP | AR | 14q24.3 (FLVCR2) |
| Martínez-Glez et al., 2010 [275] | Macrocephaly-capillary malformation: Analysis of 13 patients and review of the diagnostic criteria | Hospital Universitario La Paz, Madrid, Spain | 13 Subjects | Spain | Case series | megalencephaly, Chiari I, Sylvius aqueduct stenosis, polymicrogyria and hippocampic nodular hippocampic, septum pellucidum bifida, hemimegalencephaly, tonsillar herniation, polymicrogyria, subependymal cyst | both overgrowth/asymmetry, capillary malformations, skeletal abnormalities | Communicating | Cytogenetics, TGS, genotyping | G-banding, MLPA, SNP | - | 17p13: ABR, YWHAE, SMYD4, TRPV3 |
| Meyer et al., 2010 [276] | Mutations in FLVCR2 are associated with proliferative vasculopathy and hydranencephaly-hydrocephaly syndrome (Fowler syndrome) | Institute of Biomedical Research, University of Birmingham, Birmingham, UK | 7 Subjects, 646 Controls | Pakistan | Case series | hydranencephaly, brain stem, basal ganglia, and spinal cord diffuse clastic ischemic lesions with calcifications | glomeruloid vasculopathy of the retinal vessel, akinesia deformation sequence (FADS) with muscular neurogenic atrophy | Obstructive | Genotyping | SNP, microsatellite marker assay | AR | 14q24.3 (FLVCR2) |
| Özdemir et al., 2016 [277] | Neonatal Bartter syndrome with cholelithiasis and hydrocephalus: Rare association | Pamukkale University Faculty of Medicine, Denizli, Turkey | 1 Subject | - | Case study | isolated hydrocephalus | renal abnormalities | Communicating | Cytogenetics | Karyotyping | AR | - |
| Thomas et al., 2010 [278] | High-throughput sequencing of a 4.1 Mb linkage interval reveals FLVCR2 deletions and mutations in lethal cerebral vasculopathy | Hôpital Necker-Enfants Malades, Paris, France | 16 Subjects, 2 Controls | Turkish | Case series | brain angiogenesis, hydranencephaly | arthrogryposis/pterygia | Obstructive | TGS | Homozygosity mapping, SNP, cycle sequencing | AR | 14q24.3 (FLVCR2) |
| Visapää et al., 1999 [279] | Assignment of the locus for hydroletharus syndrome to a highly restricted region on 11q23-25 | National Public Health Institute, Helsinki, Finland | 15 Subjects, 20 Family Members, 41 Controls | Finnish | Case series | absent midline structures of the brain | micrognathia, polydactyly, anomalous eyes and nose, and a keyhole-shaped defect of the occipital bone, cleft lip or palate, anomalous or low-set ears, abnormal larynx or trachea, defective lobulation of the lungs, congenital heart defect, abnormal genitalia, and club feet | Communicating | Genotyping | Radiation-hybrid mapping, two-point and multipoint linkage analysis | AR | 11q23-25 |

### Normal Pressure Hydrocephalus

Normal pressure HC (NPH) is a form of communicating HC in which the progressive pressure of CSF is believed to result in in ventricular dilatation and further CSF accumulation. **Table 16** summarizes the genes implicated in human studies of NPH. Scm like with four mbt domains 1 (*SFMBT1*) displayed an intron 2 deletion. Cilia and flagella associated protein 43 (*CFAP43*) was found to have a nonsense mutation. The gene locus contributing to the development of ETINPH, a disorder characterized with essential tremors and idiopathic NPH, was localized to 19q12-13.31 on chromosome 19. *SFMBT1* is highly expressed in ependymal cells and epithelial cells of the brain, suggesting that a mutation in this gene locus may contribute to the dysfunctional CSF circulation [106]. Furthermore, a binding site had been identified within intron 2 of this gene locus, suggesting that the deletion of this intron, as seen in our review, will impact function [107; 108]. Deletion of cell wall biogenesis 43 (*CWH43*) in humans has also been associated with NPH [109].

**Table 16:** Normal Pressure Hydrocephalus.

Abbreviations include the following: Array comparative genomic hybridization (aCGH). Autosomal Dominant (AD). Copy number variant (CNV). Single nucleotide polymorphisms (SNP). Targeted genome sequencing (TGS). Whole exome sequencing (WES). Whole genome sequencing (WGS).
| Citation | Title | Author Affiliation | Case # | Ancestry | Study Design | CNS Phenotype | Non-CNS Phenotype | Type of Hydrocephalus | Genetic Methodology | Genetic Analysis | Inheritance | Genetic Findings |
| --- | --- | --- | --- | --- | --- | --- | --- | --- | --- | --- | --- | --- |
| Kato et al., 2011 [280] | Segmental copy number loss of SFMBT1 gene in elderly individuals with ventriculomegaly: a community-based study | Yamagata University Faculty of Medicine, Japan | 8 Subjects, 10 Controls | Japanese | Case series | isolated hydrocephalus | - | Communicating | WGS, cytogenetics | CNV, aCGH | - | 3p21.1 (12 kb deletion within intron 2 of SFMBT1) |
| Morimoto et al., 2019 [281] | Nonsense mutation in *CFAP43* causes normal-pressure hydrocephalus with ciliary abnormalities | Kagawa University, Takamatsu, Japan | 5 Subjects, Controls used | Japanese | Case study | isolated hydrocephalus | chronic sinusitis, pneumonia | Communicating | WES | Sanger sequencing | Heterozygous | 10q25.1 (c.C1058934 68T in CFAP43) |
| Sato et al., 2016 [282] | A Segmental Copy Number Loss of the SFMBT1 Gene Is a Genetic Risk for Shunt-Responsive, Idiopathic Normal Pressure Hydrocephalus (iNPH): A Case-Control Study | Yamagata University Faculty of Medicine, Yamagata, Japan | 50 Subjects, 110 Controls | Japanese | Case-Control | isolated hydrocephalus | - | Communicating | TGS | CNV | *De novo* | 3p21.1 (deletion in intron 2 of the SFMBT1) |
| Zhang et al., 2010 [283] | Genome-wide linkage scan maps ETINPH gene to chromosome 19q12-13.31 | Center for Neurosciences, Texas Tech University Health Science Center, El Paso, TX, USA | 26 Subjects | - | Case series | essential tremor | - | Communicating | Genotyping | SNP, linkage analysis | AD | ETINPH locus localized to chromosome 19q12–13.31 |

#### Metabolism

**Table 17** indicates genes involved in metabolic pathways. Mutations were seen in cytochrome c oxidase subunit 6B1 (*COX6B1*), methylenetetrahydrofolate reductase (*MTHFR),* and sulfatase modifying factor 1 (*SUMF1*). Mutations in *COX6B1* have been shown to disrupt the electron transport chain suggesting that alterations in cellular energetics can contribute to HC [110] [111]. *SUMF1* encodes formylglycine generating enzyme (FGE) involved in modifying cysteine residues in the endoplasmic reticulum [112]. *MTHFR* regulates folate metabolism, and mutations within this gene locus have been identified in congenital HC patients providing rationale to explore metabolic genes and their association with pathology [113].

**Table 17:** Metabolism.

Abbreviations include the following: Array comparative genomic hybridization (aCGH).
| Citation | Title | Author Affiliation | Case # | Ancestry | Study Design | CNS Phenotype | Non-CNS Phenotype | Type of Hydrocephalus | Genetic Methodology | Genetic Analysis | Inheritance | Genetic Finding |
| --- | --- | --- | --- | --- | --- | --- | --- | --- | --- | --- | --- | --- |
| Abdulhag et al., 2015 [284] | Mitochondrial complex IV deficiency, caused by mutated COX6B1, is associated with encephalomyopathy, hydrocephalus and cardiomyopathy | Hadassah-Hebrew University Medical Center, Jerusalem, Israel | 1 Subject, 60 Controls | Palestinian | Case study | hypotonia, cortical blindness | symmetrical left ventricular hypertrophy tricuspid regurge and pulmonary hypertension | - | WES | Variant analysis, sanger sequencing | Mitochondrial inheritance | 19q13.12 (COX6B1) |
| Cizmeci et al., 2013 [285] | Multiloculated hydrocephalus of intrauterine-onset: a case report of an unexpected MTHFR A1298C positive test result | Fatih University Medical School, Ankara, Turkey | 1 Subject | - | Case study | loculated hydrocephalus | - | Obstructive | - | - | - | 1p36.22 (MTHFR A1298C homozygosity) |
| Schaaf et al., 2016 [286] | Desmosterolosis-phenotypic and molecular characterization of a third case and review of the literature | Baylor College of Medicine, Houston, Texas, USA | 1 Subject, 1 Control, 2 Parents | - | Case study | macrocephaly, thickening of the tectum and massa intermedia, effaced gyral pattern, underopercularization, thin corpus callosum | arthrogryposis, disorder of cholesterol biosynthesis, bilateral fifth finger clinodactyly, mild cutaneous 2–4 toe syndactyly, and proximal placement of the great toes, and dysmorphic facial features | Obstructive | TGS | - | De novo | 1p32.3 (compound heterozygote for c.281G>A (p.R94H) and c.1438G>A (p.E480K) mutations in DHCR24 gene) |
| Schlotawa et al., 2011 [287] | SUMF1 mutations affecting stability and activity of formylglycine generating enzyme predict clinical outcome in multiple sulfatase deficiency | Georg August University Göttingen, Göttingen, Germany | 7 Subjects, Controls used | USA, Turkey, Switzerland, Pakistan | Case series | intellectual disability, neurodegeneration | Skeletal changes, cardiac involvement, corneal clouding, organomegaly | Communicating | TGS | - | AR | 3p26.1 (SUMF1) |

### Cell Cycle & Cytoarchitecture

**Table 18** displays genes involved in cell cycle regulation and cytoarchitecture. Mutations were seen in spindle apparatus coiled-coil protein 1 (*SPDL1*), tubulin alpha 3e (*TUBA3E*), nidogen 1 (*NID1*), tRNA splicing endonuclease subunit 15 (*TSEN15*), clathrin heavy chain linker domain containing 1 (*CLHC1*), TBC1 domain containing kinase (*TBCK*), xin actin binding repeat containing 1 (*XIRP1*), nucleoporin 107 (*NUP107*), erythrocyte membrane protein band 4.1 like 4A (*EPB41L4A*), protein phosphatase 2 regulatory subunit B delta (*PPP2R5D*), protein phosphatase 2 scaffold subunit Alpha (*PPP2R1A*), prolyl 4-hydroxylase subunit beta (*P4HB*), and crumbs cell polarity complex component 2 (*CRB2*). *SPDL1* has been shown to regulate mitotic checkpoints, and mutations arrested affected cells in metaphase [114]. *TUBA3E* maintains microtubule integrity by encoding for part of the microtubule heterodimer, alpha tubulin [115]. TSEN15 contributes to an endonuclease complex involved in tRNA splicing, and mutations affecting this gene locus can lead to defects in cell division [116]. *XIRP1* has been shown to maintain actin integrity and stability [117]. *P4HB* encodes an enzyme subunit involved in collagen formation, and mutations affecting this gene location are associated with reduced cytoarchitectural stability [118].

**Table 18:** Cell Cycle & Cytoarchitecture.

Abbreviations include the following: Array comparative genomic hybridization (aCGH).
| Citation | Title | Author Affiliation | Case # | Ancestry | Study Design | CNS Phenotype | Non-CNS Phenotype | Type of Hydrocephalus | Genetic Methodology | Genetic Analysis | Inheritance | Genetic Finding |
| --- | --- | --- | --- | --- | --- | --- | --- | --- | --- | --- | --- | --- |
| Alazami et al., 2015 [288] | Accelerating novel candidate gene discovery in neurogenetic disorders via whole-exome sequencing of prescreened multiplex consanguineous families | King Faisal Specialist Hospital and Research Center, Riyadh, Saudi Arabia | 143 Families | - | Case series | global developmental delay, autism, epilepsy, primary microcephaly, ataxia, and neurodegeneration (*among many others*) | Wide Variability | Variable | WES | Autozygosity mapping, sanger sequencing | Variable | 5q35.1 (SPDL1), 2q21.1 (TUBA3E), 15q15.1 (INO80), 1q42.3 (NID1), 1q25.3 (TSEN15), 1p33 (DMBX1), 2p16.1 (CLHC1), 12p13.32 (C12orf4), 15q26.1 (WDR93), 7q31.2 (ST7), 20q13.12 (MATN4), 4q26 (SEC24D), 5q31.3 (PCDHB4), 3p21.31 (PTPN23), 7q22.1 (TAF6), 4q24 (TBCK), 14q13.2 (FAM177A1), 4q27 (KIAA1109), 16q22.1 (MTSS1L), 3p22.2 (XIRP1), 1q41 (KCTD3), 21q22.12-q22.13 (CHAF1B), 1q42.2 (ARV1), 14q24.3 (ISCA2), 17q23.1 (PTRH2), 17p13.3 (GEMIN4), 17p12 (MYOCD), 16q22.1 (PDPR), 17p13.3 (DPH1), 12q15 (NUP107), 17q21.33 (TMEM92), 5q22.1-q22.2 (EPB41L4A), and 9q22.31 (FAM120AOS) |
| Houge et al., 2015 [289] | B56δ-related protein phosphatase 2A dysfunction identified in patients with intellectual disability | Haukeland University Hospital, Bergen, Norway | 16 Subjects, Controls used | Dutch, English, Israeli, Norwegian | Case series | Intellectual disability, seizures, callosal agenesis, hypotonia | frontal bossing, mild hypertelorism, and down slanting palpebral fissures | Communicating | Diagnostic exome sequencing, cytogenetics | Sanger sequencing, NGS, aCGH, SNP | *De novo* | 6p21.1 (c.C157T, p.P53S; c.G592A, p.E198K; c.G598A, p.E200K; c.C602G, p.P201R; c.T619A, p.W207R in PPP2R5D); 19q13.41 (c.C536T, p.P179L; c.C544T, p.R182W; c.G773A, p.R528H in PPP2R1A) |
| Ouyang et al., 2017 [290] | Cole-Carpenter syndrome-1 with a de novo heterozygous deletion in the P4HB gene in a Chinese girl: A case report | West China Second University Hospital, Sichuan University, Chengdu, Sichuan, China | 1 Subject, 2 Parents, 1 Control | Chinese | Case subject | craniosynostosis | plump anterior fontanel, growth retardation, osteopenia, and distinctive facial features, ocular proptosis, frontal bossing | - | WES | CNV, FQ-PCR | *De novo* | 17q25.3 (P4HB) |
| Rauch et al., 2015 [291] | Cole-Carpenter syndrome is caused by a heterozygous missense mutation in P4HB | Shriners Hospital for Children, Montréal, QC H3G 1A6, Canada | 2 Subjects, Controls used | - | Case series | craniosynostosis | bone fractures, ocular proptosis, and distinctive facial features | Communicating | WES | Variant analysis, sanger sequencing | *De novo*, Mosaic | 17q25.3 (P4HB) |
| Slavotinek et al., 2015 (265) [292] | CRB2 mutations produce a phenotype resembling congenital nephrosis, Finnish type, with cerebral ventriculomegaly and raised alpha-fetoprotein | University of California, San Francisco, San Francisco, CA 94143-2711, USA | 6 Subjects, 6 Parents | Ashkenazi Jewish | Case series | gray matter heterotopias | severe, congenital renal involvement; congenital nephrotic syndrome | - | WES, cytogenetics | aCGH, Karyotyping, variant analysis, sanger sequencing | AR | 9q33.3 (CRB2) |
| Zhang et al., 2020 [293] | Genetic and preimplantation diagnosis of cystic kidney disease with ventriculomegaly | Children's Hospital of Shanxi and Women Health Center of Shanxi, Taiyuan, Shanxi, 030013, PR China | 1 Subject, 2 Parents | Chinese | Case study | isolated hydrocephalus | echogenic kidneys and bowel, small fetal stomach bubble | - | WES | Variant analysis, sanger sequencing | - | 9q33.3 (CRB2) |

### Lipid Structure and Regulation

**Table 19** summarizes genes involved in lipid structure and regulation associated with HC in humans. Mutations were seen in bridge-like lipid transfer protein family member 1 (*KIAA1109*), and glucosylceramidase beta 1 (*GBA*). The *KIAA1109* ortholog in *Drosophila melanogaster* has shown to affect synaptic growth at the neuromuscular junction through modulation of phosphatidylinositol 4,5-bisphosphate (PIP^2^) [119]. *GBA* encodes for a lysosomal enzyme responsible for metabolizing glycolipids [120].

**Table 19:** Lipid Structure and Regulation.

Abbreviations include the following: Autosomal Recessive (AR). Targeted genome sequencing (TGS). Whole exome sequencing (WES).
| Citation | Title | Author Affiliation | Case # | Ancestry | Study Design | CNS Phenotype | Non-CNS Phenotype | Type of Hydrocephalus | Genetic Methodology | Genetic Analysis | Inheritance | Genetic Finding |
| --- | --- | --- | --- | --- | --- | --- | --- | --- | --- | --- | --- | --- |
| Meszarosova et al., 2020 [294] | Two novel pathogenic variants in KIAA1109 causing Alkuraya-Kučinskas syndrome in two Czech Roma brothers | Second Faculty of Medicine Charles University and University Hospital Motol, Prague | 2 Subjects | Roma | Case series | hypotonia, cerebellar malformation, lissencephaly, callosum agenesis | facial dysmorphic features, dysplastic ears, bilateral cataracts, finger contractures on both hands | - | WES | Variant analysis | AR | 4q27 (KIAA1109) |
| Shiihara et al., 2000 [295] | Communicating Hydrocephalus in a Patient with Gaucher's Disease Type 3 | Institute of Neurological Sciences, Faculty of Medicine, Tottori University, Tottori, Japan | 1 Subject, Controls used | Japanese | Case study | Isolated hydrocephalus | splenomegaly, thrombocytopenia, bilateral papilledema, motor deficits | Communicating | TGS | Restriction enzyme analysis | - | 1q22 (D409H mutation in GBA) |

### Genes of Unknown Function

**Table 20** summarizes genes that are associated with HC pathology without a clear function. Additional variants include partial 1q trisomy, tetrasomy 5p, tetraploidy of chromosome 9, trisomy 9p, and chromosome 21 trisomy. Studies that have identified mutations in chromone 6 displayed microdeletions or mosaicism of monosomy. Deletions in chromosome 8 (8q12.2-q21.2) and chromosome 16 (16q) were also identified, and microduplications in chromosome 17 (17p13.1) have been reported. The vast genetic influence on HC emphasizes importance of exploring and understanding the factors that confer genetic risk to improve diagnostic and prognostic efficiency. Autosomal and sex chromosomal location of all genetic findings included in this review is summarized in **Figure 3**.

**Table 20:** Genes of Unknown Function.

Abbreviations include the following: Array comparative genomic hybridization (aCGH).
| Citation | Title | Author Affiliation | Case # | Ancestry | Study Design | CNS Phenotype | Non-CNS Phenotype | Type of Hydrocephalus | Genetic Methodology | Genetic Analysis | Inheritance | Genetic Finding |
| --- | --- | --- | --- | --- | --- | --- | --- | --- | --- | --- | --- | --- |
| Basel-Vanagaithe et al., 2010 [296] | Familial hydrocephalus with normal cognition and distinctive radiological features | Raphael Recanati Genetics Institute, Rabin Medical Center, Beilinson Campus, Petah Tikva, Israel | 6 Subjects | - | Case series | Mega cisterna magna, midline cysts | bilateral cleft lip and palate | Obstructive | TGS, cytogenetics | X-inactivation analysis, karyotyping | - | - |
| Bernstock et al., 2020 [297] | Complex Management of Hydrocephalus Secondary to Choroid Plexus Hyperplasia | Brigham and Women's Hospital, Harvard Medical School, Boston, Massachusetts | 1 Subject | - | Case study | developmental delay, villous hyperplasia of choroid | hydrocele, abdominal distension, short stature, developmental delay, low-set ears, hypertelorism, deep-set eyes, down slanting palpebral fissure, and a bulbous nose | Communicating | Cytogenetics | aCGH | *De novo* | tetraploidy of chromosome 9 |
| Boxill et al., 2018 [298] | Choroid plexus hyperplasia and chromosome 9p gains | Viborg Regional Hospital, Viborg, Denmark | 4 Subjects | - | Case series | Choroid plexus hyperplasia | enophthalmia, hypertelorism, downslanting palpebral fissures, broad nasal bridge, bulbous nose, downturned corners of the mouth, anomalous ears, clinodactyly, single fifth finger crease, hydrocele | Communicating | Cytogenetics | Q-banding, G-banding FISH, a-CGH | *De novo* | trisomy 9p |
| Brock et al., 2012 [299] | Mosaic tetrasomy 5p resulting from an isochromosome 5p marker chromosome: case report and review of literature | Dalhousie University, Halifax, Nova Scotia, Canada | 1 Subject | Irish | Case study | mild scoliosis, refractory seizures, global delay, hypotonia | supernumerary nipples, transverse left palmar crease, square fingertips, bilateral 5th finger clinodactyly and shortened 4th and 5th metacarpals, overlapping toes bilaterally, skin pigmentary changes | Communicating | Cytogenetics | G-banding, FISH w/ WCP, aCGH | *De novo* | tetrasomy 5p |
| Brunetti-Pierri et al., 2008 [300] | Recurrent reciprocal 1q21.1 deletions and duplications associated with microcephaly or macrocephaly and developmental and behavioral abnormalities | Baylor College of Medicine, Houston, TX, USA | 36 Subjects, 50 Controls | - | Case series | attention deficit hyperactivity disorder autism, anxiety/dep ression, antisocial behavior, aggression, hallucinations | frontal bossing, deep-set eyes and bulbous nose, hypertelorism | Communicating | Cytogenetics | aCGH, FISH | AR | 1q21.2 microdeletion/microduplication |
| Cai et al., 2021 [301] | Classifying and Evaluating Fetuses with Ventriculomegaly in Genetic Etiologic Studies | Fujian Maternity and Child Health Hospital, Affiliated Hospital of Fujian Medical University, Fujian Key Laboratory for Prenatal Diagnosis and Birth Defect, Fuzhou, China | 293 Subjects | - | Case series | isolated hydrocephalus | many - large study: Cardiac, renal, facial agenesis, orthopaedic malformations, vascular malformations | - | WGS | SNP | *De-novo*, Maternal | Incidence of varying chromosomal abnormalities is higher in patients with non-isolated ventriculomegaly |
| Cambosu et al., 2013 [302] | Partial trisomy of the long arm of chromosome 1: prenatal diagnosis, clinical evaluation and cytogenetic findings. Case report and review of the literature | University of Sassari, Sassari, Italy | 1 Subject | - | Case study | macrocephaly with dolichocephaly | prominent foreheads, modest microphthalmia, flat nasal bridge, microstomia, retrognathia, small, dysmorphic ears with small lobe and short neck, and hypoplastic left kidney | - | Cytogenetics | Q-banding, FISH | - | Partial 1q trisomy |
| Capra et al., 2009 [303] | Craniosynostosis, hydrocephalus, Chiari I malformation and radioulnar synostosis: probably a new syndrome | UO Neurochirurgia, Istituto G. Gaslini, Genova, Italy | 2 Subjects | Caucasian, European | Case series | sagittal craniosynostosis, chiari I malformation, | blepharophimosis, small low-set ears, hypoplastic philtrum, radioulnar synostosis, kidney malformation, and hypogenitalism | Obstructive | Cytogenetics, TGS | Karyotyping, aCGH, MLPA | - | - |
| Castro-Gago et al., 2001 [304] | Congenital hydranencephalic-hydrocephalic syndrome with proliferative vasculopathy: a possible relation with mitochondrial dysfunction | Hospital Clínico Universitario, Santiago de Compostela, Spain | 1 Subject | - | Case study | severe encephalomalacia | muscle body inclusions | - | - | - | - | - |
| Chen et al., 2011 [305] | Prenatal diagnosis of a de novo 17p13.1 microduplication in a fetus with ventriculomegaly and lissencephaly | Mackay Memorial Hospital, Taipei, Taiwan | 1 Subject | - | Case study | mental and motor retardation, hypotonia | skeletal anomalies, clinodactyly of the fingers, hypertrichosis, congenital heart defects, craniofacial abnormalities such as microcephaly, down-slanting palpebral fissures, ptosis, hypertelorism, low-set malformed ears, smooth philtrum, micrognathia, high-arched palate, and a short neck | - | Cytogenetics | aCGH | *De novo* | 17p13.1 microduplication |
| Chen et al., 2013 [306] | VACTERL association with hydrocephalus in a fetus conceived by in vitro fertilization and embryo transfer | Mackay Memorial Hospital, Taipei, Taiwan | 1 Subject | - | Case study | isolated hydrocephalus | bilateral arthrogryposis, right radial aplasia, a right club hand, aplasia of the right thumb, hypoplasia of the left thumb, scoliosis, and an imperforate anus | - | Cytogenetics, TGS | aCGH | - | - |
| Descipio et al., 2005 [307] | Subtelomeric deletions of chromosome 6p: molecular and cytogenetic characterization of three new cases with phenotypic overlap with Ritscher-Schinzel (3C) syndrome | The Children's Hospital of Philadelphia, and The University of Pennsylvania School of Medicine, Philadelphia, Pennsylvania USA | 6 Subjects, 12 Parents, Controls used | - | Case series | Dandy–Walker malformation intellectual disability | ptosis, posterior embryotoxon, optic nerve abnormalities, mild glaucoma, atrial septal defect, patent ductus arteriosus | Communicating, Obstructive | Cytogenetics, TGS | STS mapping, FISH, direct sequencing | - | - |
| Dubé et al., 2000 [308] | A new association of congenital hydrocephalus, albinism, megalocornea, and retinal coloboma in a syndromic child: a clinical and genetic study | McGill University, Montreal Children's Hospital, Montreal, Quebec, Canada | 1 Subject | French-Canadian | Case study | global developmental delay, trigonocephaly | oculocutaneous albinism, retinal coloboma, and megalocornea, prominent metopic suture, and cryptorchidism | - | Cytogenetics | FISH, karyotyping | - | - |
| Forcelini et al., 2006 [309] | Down syndrome with congenital hydrocephalus: case report | Rua Paissandu, Passo Fundo RS, Brazil. | 1 Subject | - | Case study | isolated hydrocephalus | upslanting)palpebral fissures; flat nasal bridge; open mouth; protruding tongue; transverse palmar creases; poor Moro reflex; hyper flexibility; short stature; loose skin on nape of neck; flat facial profile; epicanthic folds; short broad hands; clinodactyly of fifth finger; gap between the first and second toes | - | Cytogenetics | - | - | Chr. 21 Trisomy |
| Garavelli et al., 2007 [310] | Megalencephaly and perisylvian polymicrogyria with postaxial polydactyly and hydrocephalus (MPPH): report of a new case | S. Maria Nuova Hospital, Reggio Emilia, Italy | 1 Subject | - | Case study | Hypotonia, dysmorphic facial features, hypoplasia of corpus callosum | Polydactyly | - | Cytogenetics | Karyotyping, FISH, SNP | *De novo* | - |
| Inui et al., 2001 [311] | A new variant neuropathic type of Gaucher's disease characterized by hydrocephalus, corneal opacities, deformed toes, and fibrous thickening of spleen and liver capsules | Osaka University, Osaka, Japan | 1 Subject | Japanese | Case study | oculomotor apraxia, rigidity, spasticity, and hyperactive deep tendon reflexes | corneal opacities, deformed toes, gastroesophageal reflux, and fibrous thickening of splenic and hepatic capsules | Communicating | WES | Cycle sequencing | *De novo* | 1342G to C (D409H) homozygous mutation |
| Kariminejad et al., 2012 [312] | Megalencephaly-polymicrogyria-polydactyly-hydrocephalus syndrome: a case report | Kariminejad-Najmabadi Pathology and Genetics Center, Tehran, Iran | 1 Subject | - | Case study | megalencephaly, polymicrogyria, Hypotonia | Polydactyly, developmental delay, bossing forehead, long philtrum, strabismus, and mild hypertelorism | - | Cytogenetics | aCGH | - | - |
| Lemire et al., 2000 [313] | Chudley-McCullough syndrome: bilateral sensorineural deafness, hydrocephalus, and other structural brain abnormalities | Royal University Hospital and University of Saskatchewan, Saskatoon, Saskatchewan, Canada | 2 Subjects | Canadian | Case series | callosal dysgenesis, gray matter heterotopia, cortical dysplasia, cerebellar dysgenesis, intellectual disability | bilateral sensorineural hearing loss, developmental delay | Obstructive | Cytogenetics, TGS | - | AR | - |
| Lowry et al., 2007 [314] | Absence of PITX2, BARX1, and FOXC1 mutations in De Hauwere syndrome (Axenfeld-Rieger anomaly, hydrocephaly, hearing loss): a 25-year follow up | Alberta Children's Hospital & University of Calgary, Calgary, Alberta, Canada | 1 Subject | - | Case study | Isolated hydrocephalus | Short stature, hyperlaxity of joints, hearing loss | Communicating | Cytogenetics, TGS | Karyotyping, FISH | AD | - |
| Matteucci et al., 2006 [315] | Sensorineural deafness, hydrocephalus and structural brain abnormalities in two sisters: the Chudley-McCullough syndrome | Department of Neurosciences, University of Pisa, Pisa, Italy | 2 Subjects | Italian | Case series | callosum agenesis, interhemispheric cyst, cerebral and cerebellar abnormalities | sensorineural hearing loss, developmental delay | Communicating | TGS, Cytogenetics | G-banding, Q-banding | AR | - |
| Naritomi et al., 1988 [316] | 16q21 is critical for 16q deletion syndrome | School of Medicine, University of the Ryukyus, Okinawa, Japan | 1 Subject | - | Case study | hypotonia | bossed forehead, epicanthal folds, hypertelorism, a flat, broad nasal bridge, a short nose with a bulbous tip, and low-set posteriorly rotated, deformed ears, high-arched<br>pallet, short<br>neck, medial toe<br>curvature | - | Cytogenetics | G-banding | *De novo* | Chr. 16q deletion |
| Østerga<br>ard et<br>al., 2004<br>[317] | Brothers with Chudley-McCullough syndrome:<br>sensorineural deafness,<br>agenesis of the corpus<br>callosum, and other<br>structural brain<br>abnormalities | The John F.<br>Kennedy<br>Institute, Gl.<br>Landevej 7, DK-2600 Glostrup,<br>Denmark | 2 Subjects | Pakistan | Case<br>series | callosal<br>agenesis,<br>colpocephaly | bilateral<br>sensorineural<br>deafness | Obstructive | TGS | - | AR | - |
| Remes<br>et al.,<br>1992<br>[318] | Fumarase deficiency:<br>two siblings with<br>enlarged cerebral<br>ventricles and<br>polyhydramnios in utero | University of<br>Oulu, Finland | 2 Subjects,<br>5 Family<br>Members,<br>Controls | - | Case<br>series | hypotonia,<br>seizures | aciduria,<br>dystonic<br>tetraplegia | Communicating | - | - | AR | - |
| Silan et<br>al., 2003<br>[319] | A new mutation of the<br>fukutin gene in a non-Japanese patient | Abant Izzet<br>Baysal<br>University<br>Duzce Medical<br>Faculty, Duzce,<br>Turkey | 1 Subject, 2<br>Parents, 1<br>Brother | Turkish | Case<br>study | hypotonia,<br>polymicrogyria in<br>several<br>cortical<br>segments<br>and severe<br>cortical<br>disorganization | congenital<br>muscular<br>dystrophy,<br>bilateral<br>buphthalmus,<br>proptosis, and<br>cataracts | Communicating | TGS | Restriction<br>enzyme<br>analysis,<br>direct<br>sequencing | AR | 9q31.2<br>(1bp<br>insertion<br>mutation in<br>exon 5 of<br>the fukutin<br>gene) |
| Tohyama<br>et al.,<br>2007<br>[320] | Megalencephaly and<br>polymicrogyria with<br>polydactyly syndrome | Kariminejad-Najmabadi<br>Pathology and<br>Genetics<br>Center, Tehran,<br>Iran | 1 Subject | Japanese | Case<br>study | megalencephaly,<br>hypotonia,<br>polymicrogyria, and<br>thin corpus<br>callosum | bilateral<br>postaxial<br>polydactyly of<br>upper and lower<br>limbs,<br>developmental<br>delay | Communicating | Cytogenetics | FISH | - | - |
| Toren et<br>al., 2020<br>[321] | Chromosomal<br>Microarray Evaluation of<br>Fetal Ventriculomegaly | Sheba Medical<br>Center, Tel<br>Hashomer,<br>Israel | 164<br>Subjects,<br>209 Controls | - | Case<br>series | Severity<br>and<br>anatomical<br>variability in<br>ventriculomegaly | - | - | Cytogenetics | Karyotyping,<br>chromosomal<br>microarray | *De Novo*,<br>Paternal | Incidence<br>of varying<br>chromosomal<br>abnormalities<br>is<br>higher in<br>patients<br>with non-isolated<br>ventriculomegaly<br>and<br>bilateral<br>ventriculomegaly |
| Vincent<br>et al.,<br>1994<br>[322] | A proposed new<br>contiguous gene<br>syndrome on 8q consists<br>of Branchio-Oto-Renal<br>(BOR) syndrome, Duane<br>syndrome, a dominant<br>form of hydrocephalus<br>and trapeze aplasia;<br>implications for the<br>mapping of the BOR<br>gene | Unité de<br>Génétique<br>Moléculaire<br>Humaine<br>(CNRS URA<br>1445), Institut<br>Pasteur, Paris | 1 Subject | - | Case<br>study | isolated<br>hydrocephalus | trapeze aplasia | - | Cytogenetics | Linkage<br>analysis | *De novo* | 8q12.2-q21.2<br>deletion |
| Wadt et<br>al., 2012<br>[323] | Fetal ventriculomegaly<br>due to familial<br>submicroscopic terminal<br>6q deletions | University<br>Hospital of<br>Copenhagen, | 2 Subjects,<br>16 Family<br>Members | - | Case<br>series | hypotonia | oromotor<br>difficulties,<br>hypermobility,<br>high, flat | Obstructive | Cytogenetics | Karyotyping,<br>aCGH,<br>MLPA | Variable<br>expressivity | terminal<br>deletion of<br>chromosome 6q |
|  |  | Rigshospitalet, Denmark |  |  |  |  | forehead, bilateral ptoses, |  |  |  |  |  |
| Walker et al., 1996 [324] | Prenatal diagnosis of ring chromosome 6 in a fetus with hydrocephalus | Children's Hospital Research Foundation, Cincinnati, Ohio, USA | 1 Subject | - | Case study | microcephaly, seizures | severe bilateral hearing loss, and global development delay | Obstructive | Cytogenetics | - | *De novo* | chromosome 6/monosomy 6 mosaicism |
| Wang et al., 2020 [325] | Prenatal diagnosis of chromosomal aberrations by chromosomal microarray analysis in foetuses with ventriculomegaly | West China Second University Hospital, Sichuan University, No. 20, Section 3, Renminnan Road, Chengdu, 610041, Sichuan, China | 548 Subjects | - | Case series | agenesis/hypoplasia of the corpus callosum, Dandy–Walker malformation, migration abnormality, and holoprosencephaly | renal, cardiac, and skeletal anomalies | - | WGS, cytogenetics | SNP, Karyotyping, CNV | *De-novo*, maternal, balanced translocation | Incidence of varying chromosomal abnormalities (13 types) is higher in patients with severe ventriculomegaly |
| Welch et al., 2003 [326] | Chudley-McCullough syndrome: expanded phenotype and review of the literature | Gallaudet University, Washington DC, USA | 3 Subjects, 13 Family Members | Western European | Case series | obstruction of the foramen of Monro, arachnoid cyst, partial agenesis of the corpus callosum, and abnormalities in the migration of cerebellar cells | deafness | Obstructive | TES | Linkage analysis | AR | - |
| Yoshioka et al., 1994 [327] | Clinical spectrum and genetic studies of Fukuyama congenital muscular dystrophy | Kobe General Hospital, Japan | 48 Subjects | Japanese | Case series | lissencephaly | weakness, joint contractures, delayed motor development | Communicating | - | - | AR | - |

### Global Burden of Genetic Hydrocephalus

We next aimed to quantify the country of origin for patients included in this review (**Figure 5**). Given the wide range of HC disease burden across the world [4], we aimed to determine if genetic studies of HC were similarly representative. What is clear is that for regions of the world where HC prevalence is highest (Africa, East Asia, etc.), there is an obvious lack of genetic studies of HC of any kind. For example, there is not a single study performed by authors in Sub-Saharan Africa (SSA) or including people from SSA. Given that African genomes are the most diverse and complex with generations of environmental pressures (including emerging pathogens) shaping the genome, understanding genetic risk factors in these populations is essential. While epidemiological estimates of the contribution of genetically linked forms of HC is not feasible at present, these data begin to highlight disparities in representation of genetic studies and the need for large-scale genetic studies of HC in diverse populations. These data also provide a reasonable estimate of the potential burden, although likely underestimated, contribution of genetic factors contributing to HC. It is our hope that this review highlights the diverse mechanisms underlying HC, the complex molecular pathways that may contribute to HC pathogenesis, and the need to greatly expand the representation of diverse peoples in HC genetics research.

**Figure 5.**
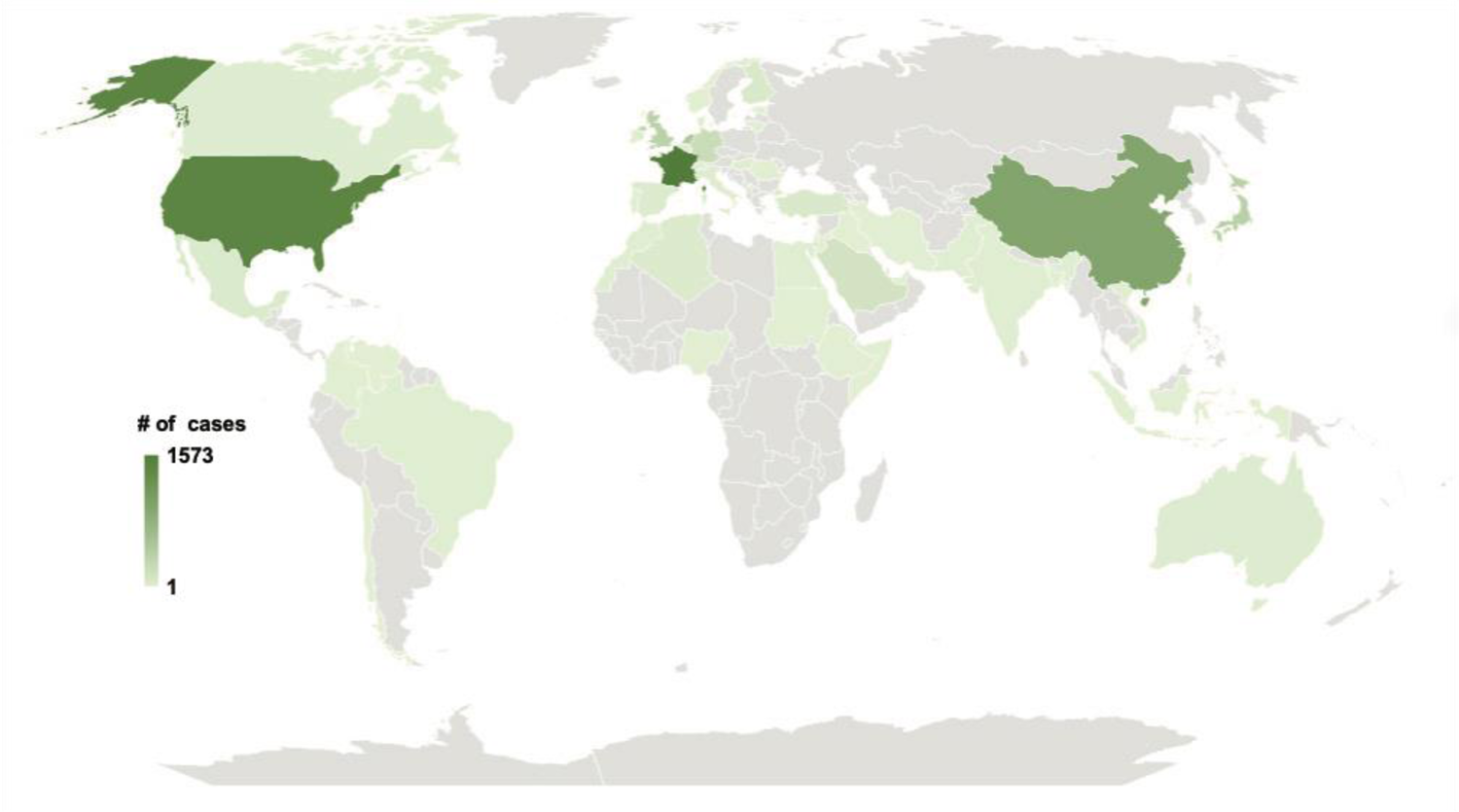
Heatmap of the globe demonstrating the country of origin for patients with genetic contributions to hydrocephalus. Figure created with OpenStreetMap.

## Discussion

HC is a complex, heterogenous condition that can be a component of a wide range of genetic conditions and can be caused by a variety of preceding environmental factors. Because HC is a component of many syndromes with a wide range of concomitant phenotypes, understanding the genetic pleiotropism of contributing genes is important for delineating the pathophysiologic basis of the disease. This review provides a broad overview of the associations between genetic mechanisms underlying HC. The variability in phenotypes observed, methodology used to uncover genetic information, and wide range of validation of genetic findings highlights the major challenges in the field. While many studies are descriptive, a wide range of hypotheses are generated based on implicated genes and potential mechanisms. Specifically, many studies implicate alterations in neurogenesis and primary brain development, as opposed to direct alterations in CSF regulation, as potential pathophysiologic mechanisms. Overall, as genomic technologies become more ubiquitous in clinical practice and more patients undergo unbiased genomic sequencing, our understanding of HC will improve. However, there are several limitations and points to consider as this field evolves.

An ongoing challenge in human genetics is proving causality of implicated genetic findings. Classical validation technique requires reproducing the implicated mutation (if evolutionarily conserved) in a model organism such as a mouse or rat. However, the physiologic regulation of CSF and mechanisms underlying brain development are markedly different in these model organisms and often do not recapitulate human disease. Many genes underlying HC are associated with other phenotypes, and it may not be possible to identify a secondary causative genetic factor that unmasks the phenotype. Since this approach relies on the gene product being evolutionarily conserved, identification of human-specific disease mechanisms is impaired. Alternative approaches to determining the relationship of a gene variant to a HC phenotype include structural biologic modeling of presumed deleterious mutations; however, this approach does not consider physiological and phenotypic heterogeneity. Similarly, these approaches often rely on protein expression in prokaryotic systems, limiting interpretation of post-translational modifications and other physiologic contributors to protein function.

Based on the significant co-occurrence of traits affecting other organ systems, it is likely that genes associated with HC display significant pleiotropy. A simplistic model of monogenic contributions to HC is unlikely to capture the genetic etiology of most cases. Even among monogenic contributions to HC, there is significant phenotypic and genetic variability (i.e., L1CAM). As quantitative genetic methodology improves to identify polygenic contributors to disease, we suspect that a much larger proportion of cases will have polygenic contributions. Because HC is a heterogenous disease, accrual of large numbers of ‘homogenous’ cases are needed to accurately quantify reproductible genetic associations.

The variability in genomic technology used to determine potential genetic contributions to HC is significant. Agnostic methods such as genome wide association studies (GWAS), transcriptome-wide association studies (TWAS), whole-exome sequencing (WES), and whole-genome sequencing (WGS) have been used, but are limited by cost, sample size, and technical expertise involved in analysis. In contrast, targeted sequencing approaches rely on hypothesis-driven identification of implicated genetic loci introducing significant experimental bias.

Our review highlights that most genetic studies of HC are performed in countries where disease burden, paradoxically, is amongst the lowest in the world. This reflects disproportionately low resources for genetic studies in low- and middle-income countries. For example, Sub Saharan Africa the most genetically diverse and complex region in the world, where the burden of HC is also the highest, yet there are no genetic studies of HC of any kind in these populations. Although the burden of HC is largely the result of infections, the genetic contributors to infection susceptibility are largely uncharacterized in these populations. Evolutionary selection pressures have been differentially shaped by exposure to infectious pathogens, geographic shifts of ancestral peoples, and population isolation. Therefore, understanding genetic factors specific to these populations is paramount to improve secondary prevention and moving towards non-surgical treatment options.

Advances in genetic technology and interpretation coupled with decreased costs will garner a new era of precision medicine that can be applied in the clinic. The extent to which genetic information may guide treatment in HC has not been fully realized. As more patients are rigorously studied using complementary and convergent genomic approaches coupled with long-term clinical outcomes, we may be able to incorporate genetic information into clinical care. Owing to the genetic architecture of HC highlighted here and across many studies, we anticipate that creation of polygenic risk scores (PRS) may be the most clinically meaningful and practical for disease prognostication and understanding comorbid disease risks.

## Conclusions

HC is a phenotypically and genetically complex disease. While the literature describing the genetic causes of HC is vast, this comprehensive review highlights opportunities for further mechanistic study and disparities in ancestral representation. The varying rigor with which genetic studies are conducted highlights the challenge of determining causality of implicated genomic alterations, inadequacies of current model systems, and the need for human-specific molecular validation studies. What is clear is that our genetic understanding of HC is incomplete and our understanding of pleiotropy of implicated HC genes requires further maturation. This study represents the first large-scale systematic literature review of the genetic basis of HC in humans and highlights many areas ripe for future investigation.

## Data Availability

All data produced in the present work are contained in the manuscript.

